# Context and health: a systematic review of natural experiments among migrant populations

**DOI:** 10.1101/2023.01.18.23284665

**Authors:** Louise Biddle, Maren Hintermeier, Diogo Costa, Zahia Wasko, Kayvan Bozorgmehr

## Abstract

**Background:** Studies on contextual effects on health often suffer from compositional bias and selective migration into contexts. Natural experiments among migrants may allow for the causal effect of contexts in generating health inequalities to be examined. We synthesised the evidence on and health from natural experiments among migrant populations.

**Methods:** Systematic literature review searching the databases PubMed/MEDLINE, The Cochrane Library, Web of Science, CINAHL and Google Scholar for literature published until October 2022. 5870 studies were screened independently in duplicate using pre-defined criteria for inclusion: quantitative natural experiment methodology, migrant study population, context factor as treatment variable and health or healthcare outcome variable. Synthesis without meta-analysis was performed following data extraction and quality appraisal.

**Findings:** The 46 included natural experiment studies provide causal evidence for the negative effects of neighbourhood disadvantage on physical health and mortality, while finding mixed effects on mental health. Studies comparing migrants with those that stayed behind demonstrate the detrimental effects of migration and adverse post-migratory contexts on physical health and mortality, while demonstrating favourable effects for mental health and child health. Natural experiments of policy contexts indicate the negative impacts of restrictive migration and social policies on healthcare utilization, mental health and mortality as well as the positive health effects when restrictions are lifted.

**Interpretation:** Natural experiments can serve as powerful tools in reducing bias through self-selection. With careful consideration of causal pathways, results from migration contexts can serve as a magnifying glass for the effects of context for other population groups. Studies demonstrate the negative impacts for health which lie at the nexus of context and health. At the same time, they uncover the potential of health and welfare programs to counteract the disadvantages created by othering processes and promote healthy (post-migratory) contexts.

**Funding:** German Science Foundation (FOR: 2928/ GZ: BO5233/1-1).

**Panel 1: research in context:** *Evidence before this study:* We searched PubMed/MEDLINE to identify pre-existing reviews on contextual effects on health with the following search terms: ((review[Title/Abstract]) AND (((context[Title]) OR (neighbourhood[Title])) OR (small-area[Title]))) AND (health[Title]). Eight reviews existed and pointed to consistent, but small effects of neighbourhood disadvantage on physical and mental health outcomes, as well as on child and adolescent health. However, these reviews also point to the methodological shortcomings of most studies, which are unable to disentangle compositional from contextual effects. In order to improve causal inference, natural experiments are needed. Natural experiments have previously delivered crucial evidence on the causal effects of public health interventions including suicide prevention, air pollution control, public smoking bans and alcohol taxation.

*Added value of this study:* This review uses natural experiments among migrants to contribute to the existing evidence base by synthesising insights on the causal mechanism of contextual effects. It uses migration as an example to assess how contextual factors, ranging from policy environments to neighbourhood characteristics, generate or exacerbate inequalities among societies. We thereby circumvent and avoid limitations of other reviews on these topics, by exploiting five main sources of variation of contextual exposures: residential dispersal, arbitrary eligibility cut-offs, on-/off-timing of events, regional variation, and place of birth. Based on these, we identify three main types of natural experiments among migrant populations: 1) Studies “using” migration as an example to analyse contextual health effects or neighbourhoods in the post-migration phase; 2) Studies examining interactions between changes in environmental factors following migration processes as compared to those staying behind; and 3) Studies using natural experiments to study policy effects. The synthesised evidence confirms and provides causal evidence for the negative effects of neighbourhood disadvantage on physical health and mortality, while effects on mental health are mixed. The body of literature demonstrates that migration processes can unfold detrimental effects on physical health and mortality through adverse post-migratory contexts, while also demonstrating favourable effects for mental health and child health depending on the respective context. Our synthesis further provides causal evidence for the negative impacts of restrictive migration and social policies on healthcare utilization, mental health and mortality as well as the positive health effects when restrictions are lifted.

*Implications of all available evidence:* The evidence presented here demonstrates the health disadvantages faced by migrants in the immediate post-settlement phase, which are exacerbated by restrictive health, social and visa policies. More broadly, however, the evidence points to neighbourhood disadvantage as a crucial and causal mechanism underlying health inequities at a societal level. At the same time, studies uncover the potential of health and welfare programs to counteract the disadvantages created by othering processes and instead promote healthy contexts. Such evidence is valid beyond migrant populations and allows inference of the positive effects of inclusive health and welfare programs for other marginalized groups and the population as a whole.

## INTRODUCTION

A long research tradition in public health and social epidemiology has shown important effects of contextual factors of the place of residence on diverse health outcomes, including mortality, chronic diseases and mental health ^1^. Such research has examined the influence and impact on health of socio-demographic (e.g. area-level poverty, inequality or deprivation) ^2–4^, physical (e.g. green space, noise pollution or housing quality) ^5,6^, or social attributes of neighbourhoods (e.g. social capital, safety and community participation) ^7–9^. Furthermore, health and social policies also shape living contexts and thereby impact people’s health status ^10,11^. Adverse contextual conditions have been found to disproportionately affect vulnerable subgroups ^12–14^. However, studies on contextual effects are often limited in their ability to disentangle contextual from compositional effects. Because of self-selection of individuals into neighbourhoods, many studies suffer from compositional bias and are thus unable to draw causal inferences. Experiments allocating individuals to places of residence exist ^15^ but are rare for ethical and practical reasons.

Natural experiments allow for the causal effect of contexts to be examined ^1^. They can be broadly defined as events where the allocation into exposed and unexposed groups are not under the control of the researcher and subject to a random (or quasi-random) process ^16,17^. The allocation may be the result of truly “natural” forces, such as extreme weather or geographical events, but also phenomena under human control, such as quasi-random policy introduction or administrative processes. Natural experiments minimise selection bias in the allocation process. This makes them particularly valuable for contextual health research, including the causal effects of public health policies ^16,18^ or social and economic policies ^17^.

Natural experiments are scarce, but when they intersect with human migration they offer powerful opportunities to study the effects of changing environments and contexts, including the consequences of migration-specific, but also broader health and social policies. Human migration is a large-scale, global phenomenon involving highly diverse groups of individuals and circumstances ^19,20^. It takes place in a contested political space in which substantial variation exists with respect to exclusionary or inclusionary forces ^21^, which produce health inequalities through mechanisms linked to economic and social policies ^22^. Migration thus serves a lens function to learn lessons for public health about the generation and (re)production of inequalities in the population. We hence aimed to systematically review and synthesise the evidence on contextual effects on health and healthcare outcomes from research using natural experiments among migrant populations.

## METHODOLOGY

We conducted a systematic literature review in line with the Preferred Reporting for Systematic Reviews and Meta-Analyses (PRISMA) guidelines. A protocol of the review strategy was registered in the International Prospective Register of Systematic Reviews (PROSPERO) on 28 April 2020 (CRD42020169236).

We searched the scientific databases PubMed/MEDLINE, The Cochrane Library, Web of Science/Knowledge (WoS) and CINAHL using English search terms within the search blocks migrants, natural experiments or dispersal programs, health outcome, and contextual effects (see Appendix Table S1) without limitations to the start or end date of publication. Additionally, we used the search engine Google Scholar to find further academic literature published since 2000 that might not be available in other databases ^23^. Results in all databases were restricted to English and German language articles. Searches were conducted on 4 February 2020, and updated (in PubMed/MEDLINE, WoS and CINAHL) on 13 October 2022 to capture the most recent publications. A total of 7150 items were identified in the search, with 5870 remaining for screening after deduplication (Figure 1).

**Figure 1:**
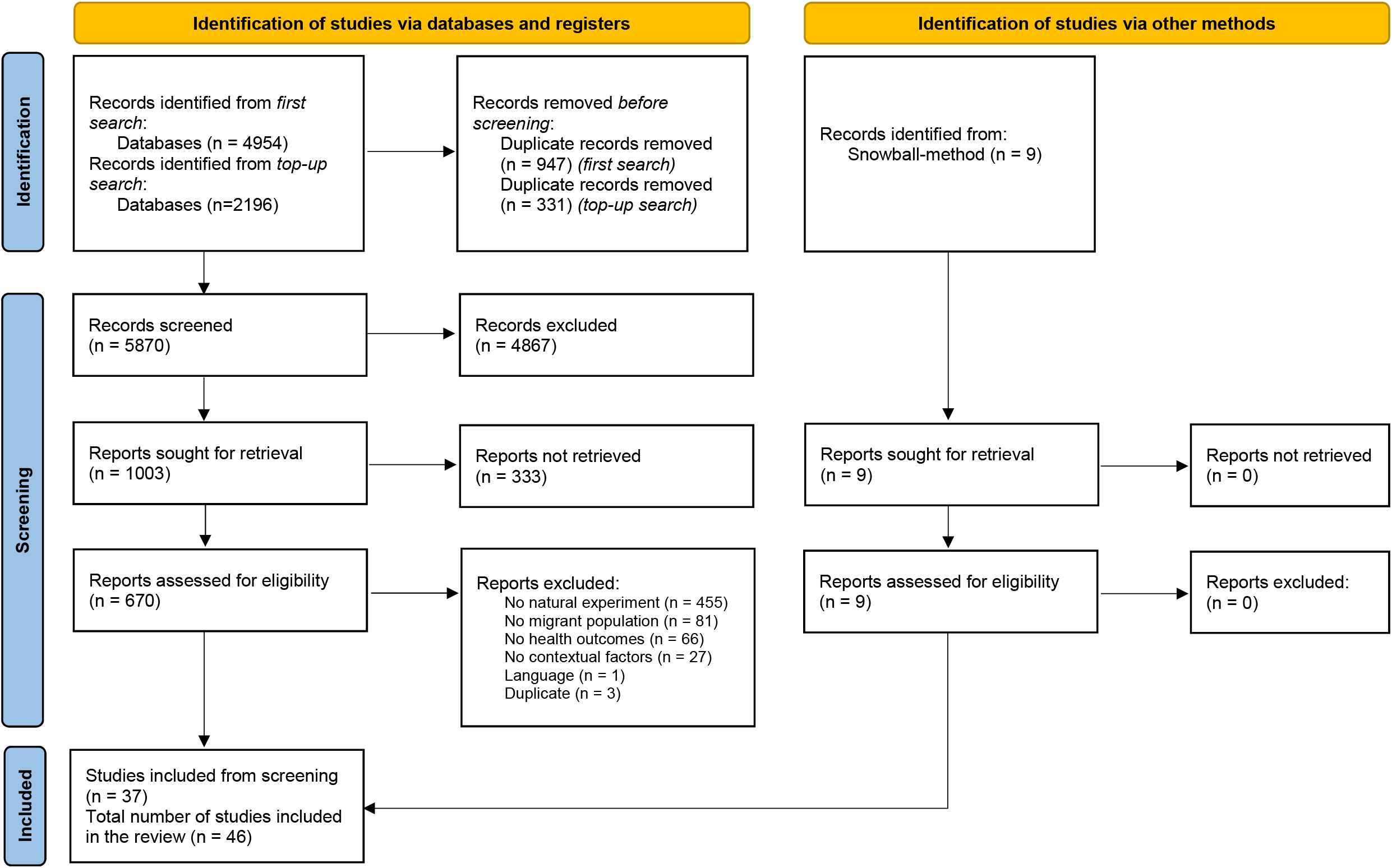
PRISMA flow-chart. *From:* Page MJ, McKenzie JE, Bossuyt PM, Boutron I, Hoffmann TC, Mulrow CD, et al. The PRISMA 2020 statement: an updated guideline for reporting systematic reviews. BMJ 2021;372:n71. doi: 10.1136/bmj.n71.

We conducted the screening process in two steps. In a first step, two members of the review team independently screened the title and abstract of each item, applying pre-defined inclusion/exclusion criteria (see Appendix Table S2). Studies were excluded if they did not cover a migrant population, did not use quantitative research methods, or did not report health outcomes and/or assess contextual factors. In a second step, we reviewed full-text articles using the same exclusion criteria and additionally excluded studies which did not exploit a natural experiment. We considered studies as natural experiments if they fulfilled the following criteria: allocation was not under the control of the research team, the source of the variation in exposure was (quasi-)random, participants were unable to self-select the exposure. Articles reporting a natural experiment design but not fulfilling these criteria were excluded (Appendix Table S3). Articles using other terminology (quasi-experiment, natural quasi-experiment, quasi natural-experiment) were included if fulfilling the above criteria. Each item was screened independently by two members of the review team, with discrepancies resolved through joint discussion. In cases where consent could not be reached, articles were discussed by all members of the research team. References of the included studies were additionally screened for further relevant articles using a snowballing methodology.

Data on the following were extracted for all studies: bibliographic information, study characteristics (incl. research objectives, context and period of study, theoretical foundation and sampling), research design, analysis strategy, study population, contextual exposures, and health or healthcare outcome variables used, confounders, as well as findings and limitations as reported. Data for all items were extracted in duplicate and consensus reached through joint discussion. Quality Appraisal of the included studies was carried out independently by pairs of two reviewers using an adapted version of the “Quality Assessment Tool for Quantitative Studies” developed by the Effective Public Health Practice Project (EPHPP) ^24^. We adapted the following items to suit the natural experiment methodology of included articles (Appendix Table S4). Discrepancies in scoring were discussed in the team to reach consensus.

Results were analysed using a synthesis without meta-analysis (SWiM) methodology ^25^. In a first synthesis step, two researchers inductively created several groupings of the included articles based on type of contextual exposure, health outcome and the source of variation in exposure underlying the natural experiment (Panel 2). These groupings were further refined in joint discussions of the review team. Studies were assigned to one of the following four groups based on the context factor assessed: contextual factors of the place of residence in receiving countries, migration-context interactions, policy contexts and cultural factors as context. Health outcomes were grouped into physical health (including non-communicable diseases, mortality and self-rated health), mental health (including symptoms of mental illness, psychiatric hospital admissions and substance abuse), child health (including self-reported mental and physical health, anthropometric measures and birth outcomes) and healthcare utilisation (including healthcare visit and cost data). Using the groups as a guide, study methodologies and results were discussed in six repeated rounds of synthesis meetings. The main synthesis was based on the groupings by health or healthcare outcomes. Due to the high heterogeneity of the included studies meta-analysis was not possible.

### Panel 2: Categorisation of the source of (random) variation in exposure underlying the natural experiments

Besides considering the events which give rise to the possibility of a natural experiment design, it proved helpful to consider the source of the variation in exposure in order to further understand the success of the (quasi-)random allocation. We divided the natural experiments into five groups. These differ with respect to the unit of allocation (individual/community), the temporality and the underlying assumptions made. We briefly describe each group below:

1. Residential dispersal Studies using residential dispersal as the source of variation rely on some policy which disperses individuals into different geographical units based on, for example, administrative quota or a ballot. Here, allocation may occur on the individual or household level. The extent to which the process can be regarded as ‘random’ depends on the characteristics considered in the dispersal process. For example, Gibson et al. (2010) describe a migration ballot which is determined purely by chance, while some dispersal systems for refugees may be affected by family composition, health status and nationality and are thus only quasi-random (Wenner et al. 2020). These designs are strengthened by coexistent residential mobility restrictions, which reduce the possibility of selective secondary migration, e.g. in the case of many asylum processes where individuals are obliged to reside in the municipality or region where they first made their asylum claim for a given period of time. Residential dispersal allows for the use of contemporaneous comparison groups.
2. Arbitrary eligibility cut-offs These studies make use of a policy or programme with eligibility criteria that have some cut-off which can be considered to be set arbitrarily. Thus, individuals at either side of the cut-off can be said to be comparable in all aspects except for their treatment status. Our review includes studies which use eligibility for welfare benefits after an arbitrary length of time (e.g. Bozorgmehr & Razum 2015) or an arbitrary age cut-off (e.g. Hainmueller et al. 2017) as well as a study which uses the arbitrary water-level rise following dam construction (e.g. Hwang et al. 2011). Arbitrary eligibility cut-offs allocate the exposure at an individual or household level, and allow for contemporaneous comparison groups.
3. Timing of policy introduction/ event Several studies make use of the timing of a policy introduction or an event as the source of variation in the exposure. They make before/ after comparisons and thus do not allow for the use of contemporaneous comparison groups. The rationale behind classifying these studies as natural experiments is that the timing can be considered to be to some extent arbitrary or unpredictable and the policy change usually applies to an entire country, thus minimising bias through self-selection into exposed and unexposed groups. The exposure is allocated at a community level. Examples of this variation in exposure include the introduction of a paternal leave policy (Honkaniemi et al. 2021), the restriction of healthcare access (Juanmarti Mestres et al. 2020) and the introduction of a repatriation policy (Fu & VanLandingham 2010). One study (Lopez et al. 2016) makes use of the unpredictable timing of an immigration raid as the source of variation in the exposure.
4. Regional variation The studies making use of regional variation in the exposure utilise the uneven implementation of policies (e.g. Kaushal 2007, Ye & Rodriguez 2021) or the uneven effects of a natural disaster (Hori et al. 2021) as their source of exposure variation. Allocation necessarily occurs at the community level. Again, these are classified as natural experiments because of the arguably arbitrary pattern of policy introduction which can be considered to be unrelated to other characteristics of the region or its inhabitants or the outcome studies. These regional comparisons allow for the use of contemporaneous comparison groups, but bias through self-selection can often not be ruled out when utilising regional variation on its own.
5. Place of birth Finally, some studies make use of the randomness of the place of birth as the source of variation in their exposure. The argument presented in these studies is that the comparison groups can be considered to be equal in all aspects except the context of birth, the effects of which they subsequently examine. This approach allows for contemporaneous comparison groups, with allocation at both individual and community levels possible. Black et al. (2015) analyse the proximity of the place of birth to a railroad in the Southern United States as an instrument for migration and examine its effect on mortality. Hajdu & Hajdu (2015) examine the effect of the overall subjective well-being in the country of birth on individual well-being after migration.

## RESULTS

A total of 1003 items remained after abstract screening, with 37 items remaining after full-text screening (Figure 1). Nine items were identified by the snowballing method, resulting in a total of 46 studies included for analysis. Almost all were published after 2007, with only one included study from 1963. While studies assessing migrations/context interactions dominated in the 2010s, when assessing the so-called “healthy migrant effect” was a particular concern, studies looking at contextual factors of the place of residence and policy contexts have risen to prominence in more recent years (Figure 2a). 33 studies were conducted in countries receiving migrants (n=18), refugees (n=13) and resettlers (n=1). These were based in Europe (n=21) ^26–46^ and the United States of America (USA) (n=12) ^47–58^ (Table 1). Studies in China (n=3) ^59–61^ and the USA (n=2) ^62,63^ focused on internal migrants. Eight studies conducted research in both the country of destination and the country of origin, covering a natural experiment in Vietnam and the USA (n=3) ^64–66^ and one in Tonga and New Zealand (n=5) ^67–71^. 22 studies report only data on migrants ^28–32,34–36,38,42–48,50,51,56,57,60,61^, while the others include migrants as a subsample of a broader population, with migrants making up between 5-83% of the total sample (Appendix Table S6). Median of the total sample size is 10 347 (254 – 5 009 832), excluding two ecological studies which reported sample size in person-years ^28^ and person-quarters ^45^. A large majority of studies use a cohort analytic design (n=31), with others being cohort studies (n=1), cross-sectional studies (n=7), repeated cross-sectional studies (n=2), ecological studies (n=3) or combining both cross-sectional and cohort-analytic study designs (n=2) (Table 1).

**Table 1:**
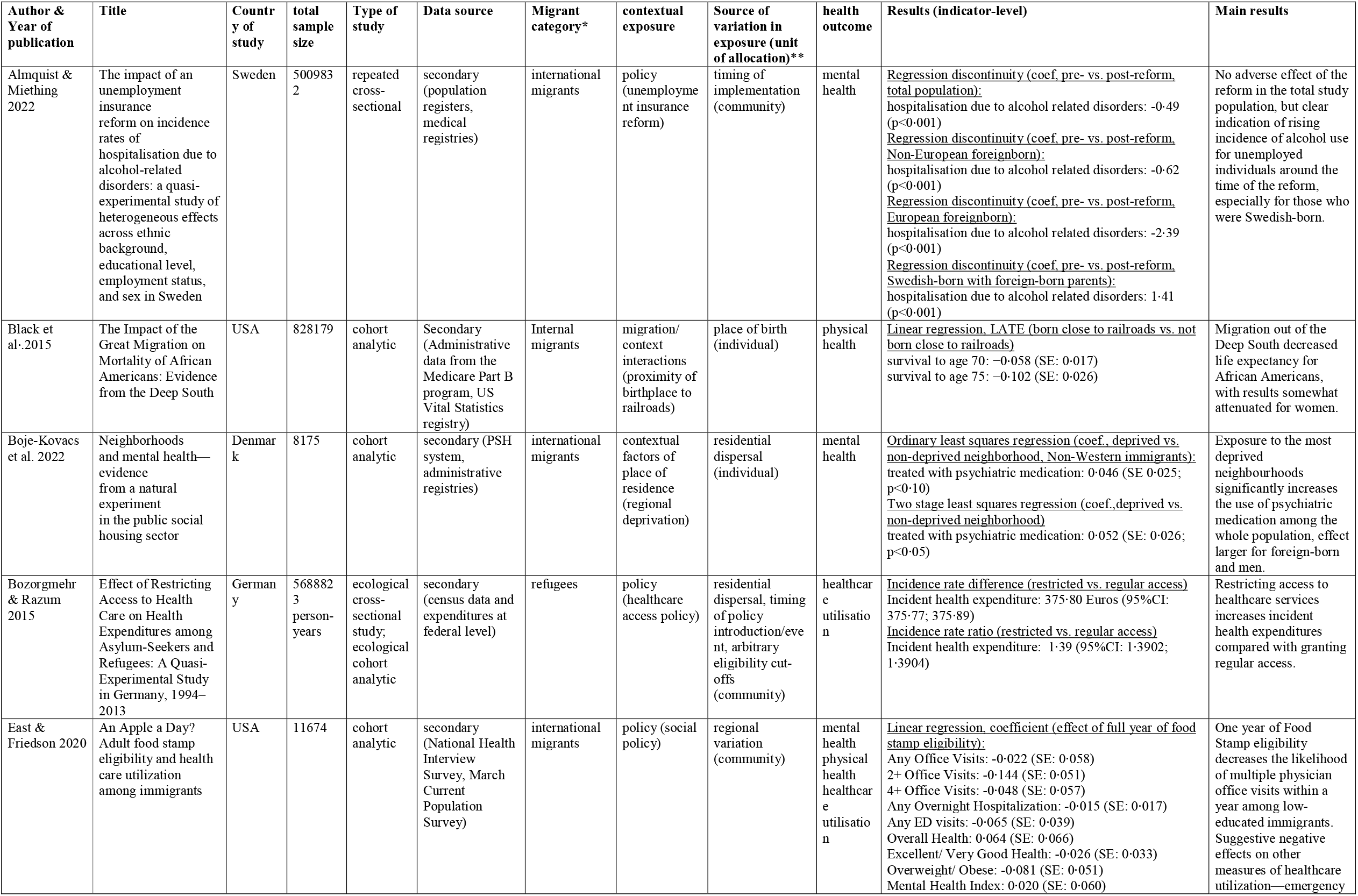
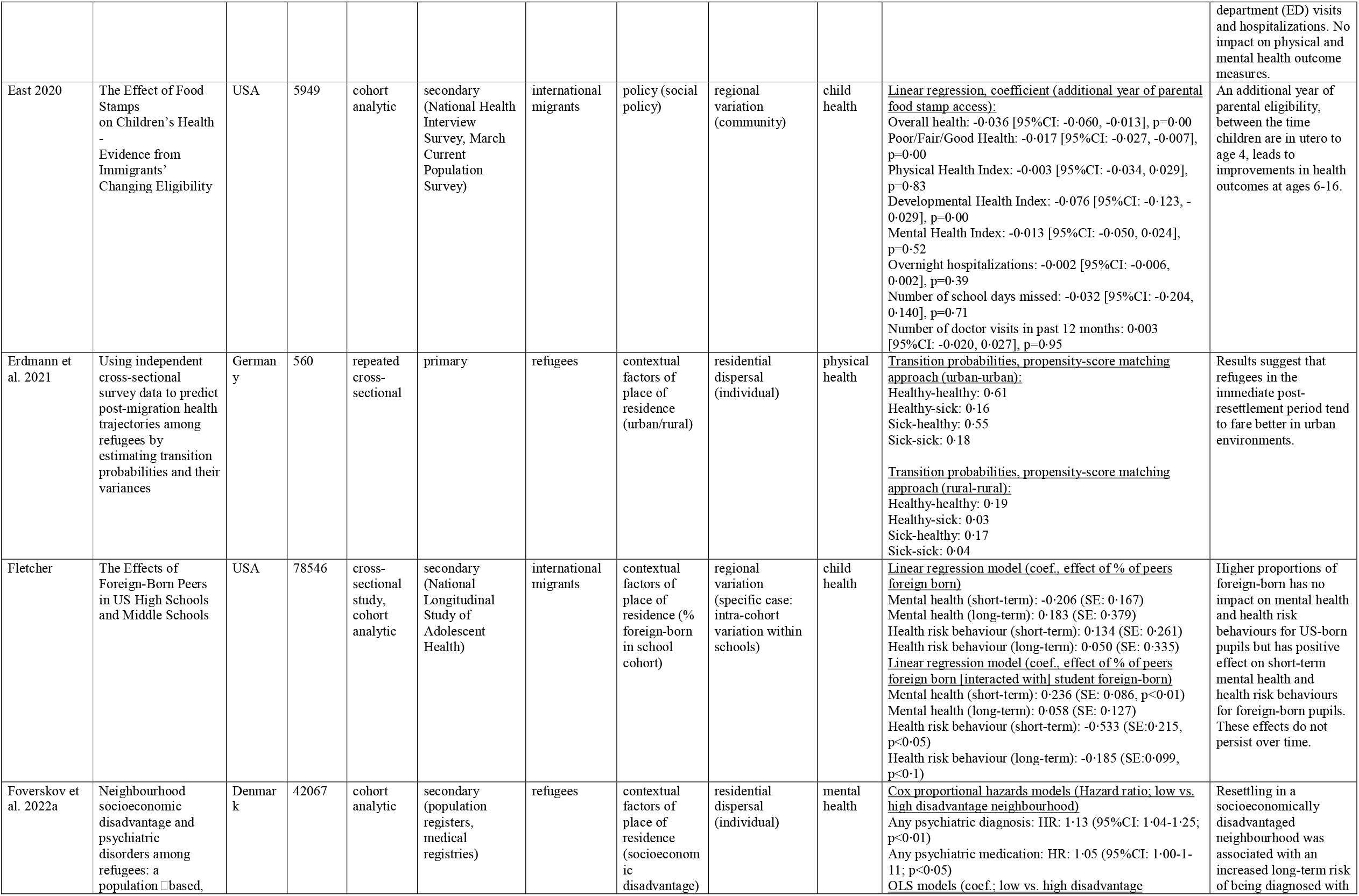
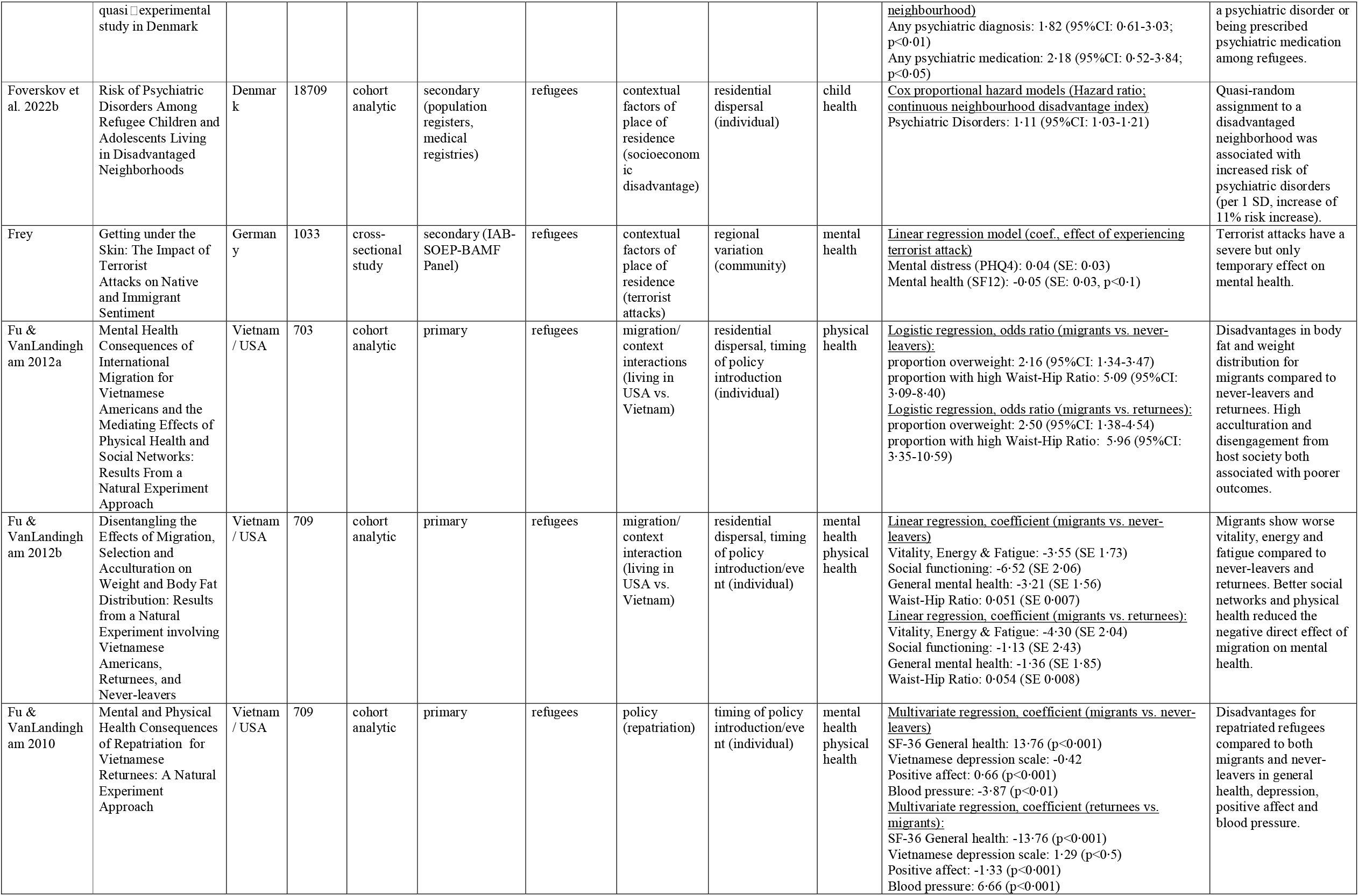
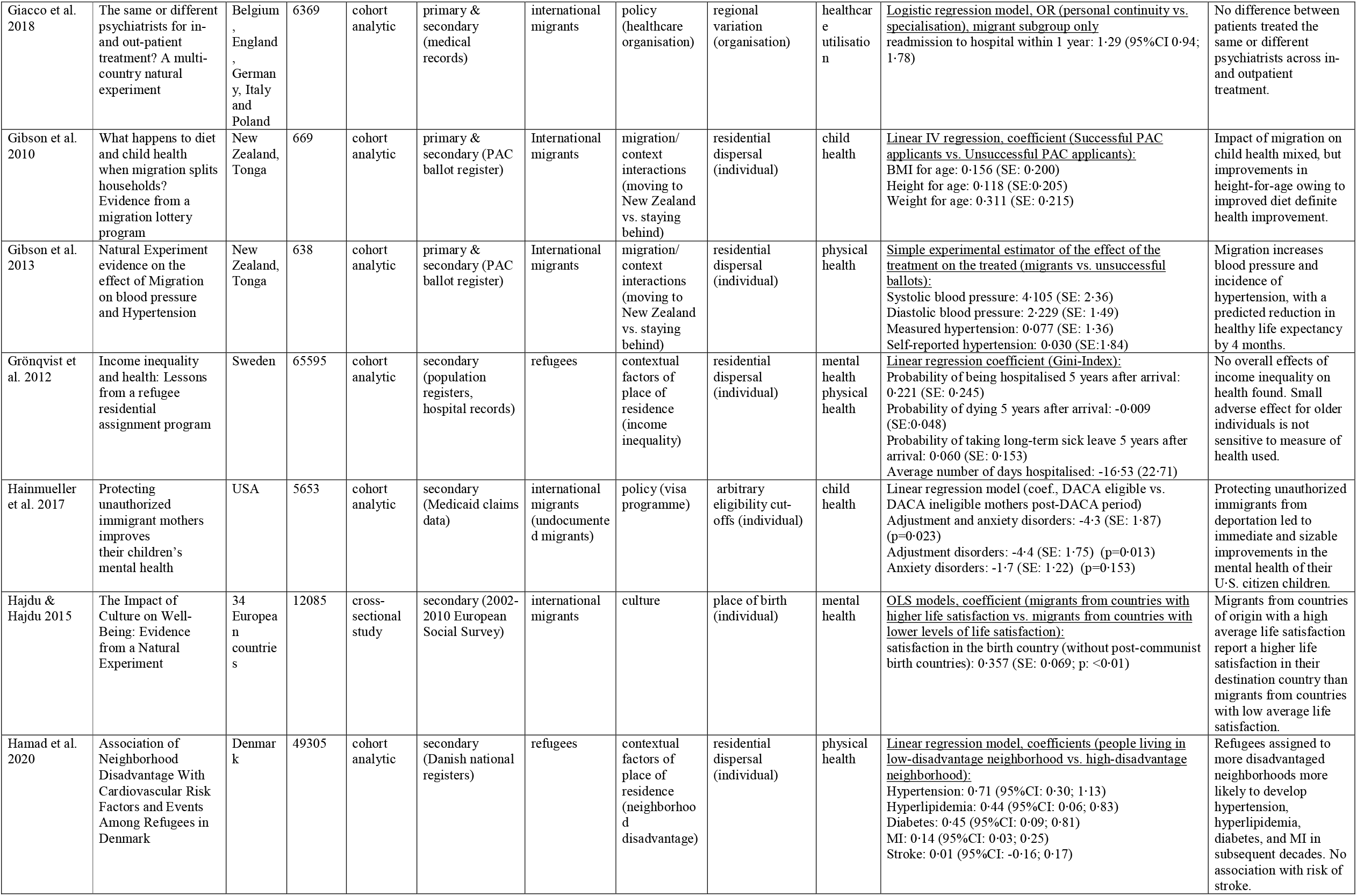
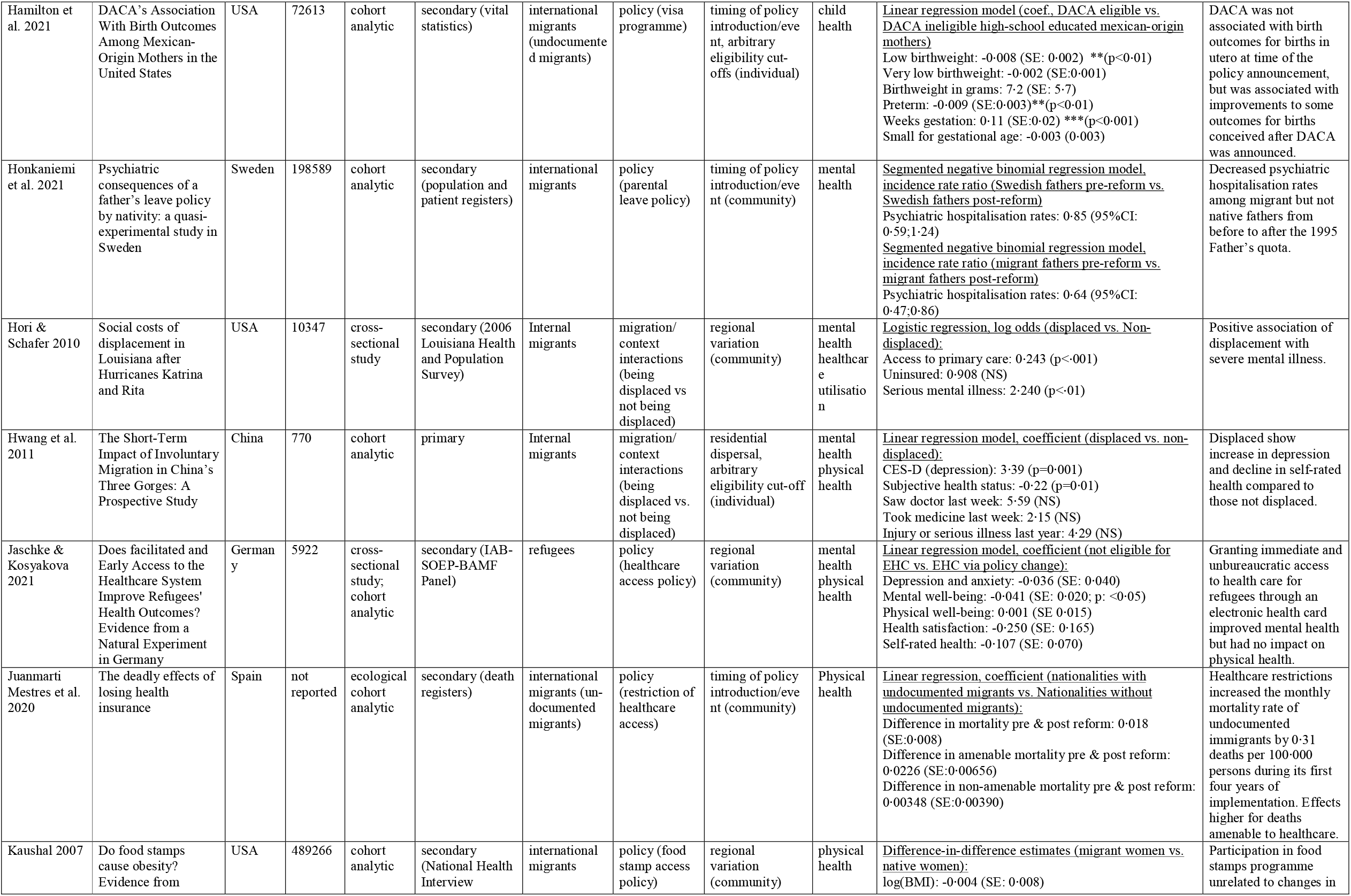
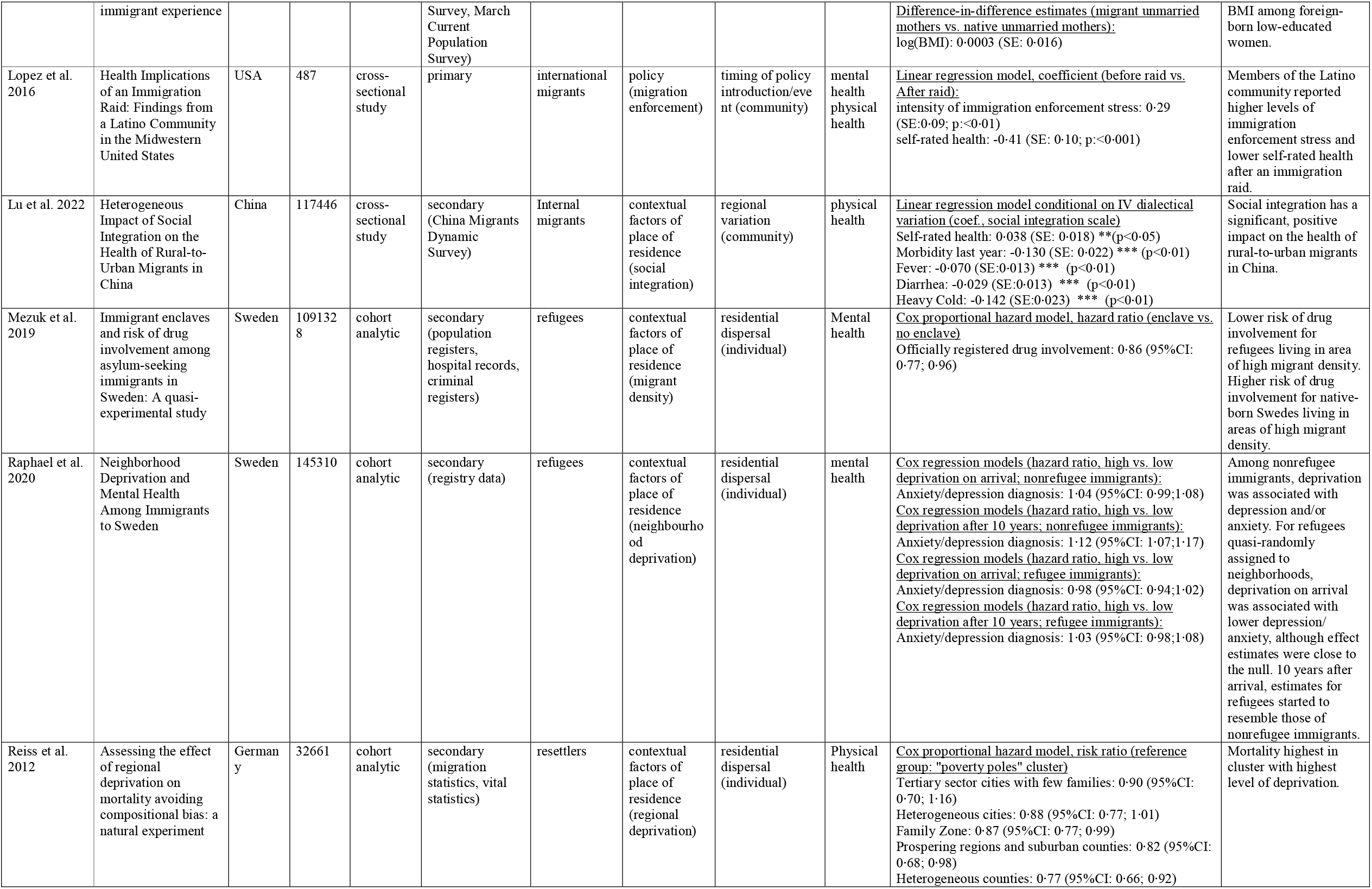
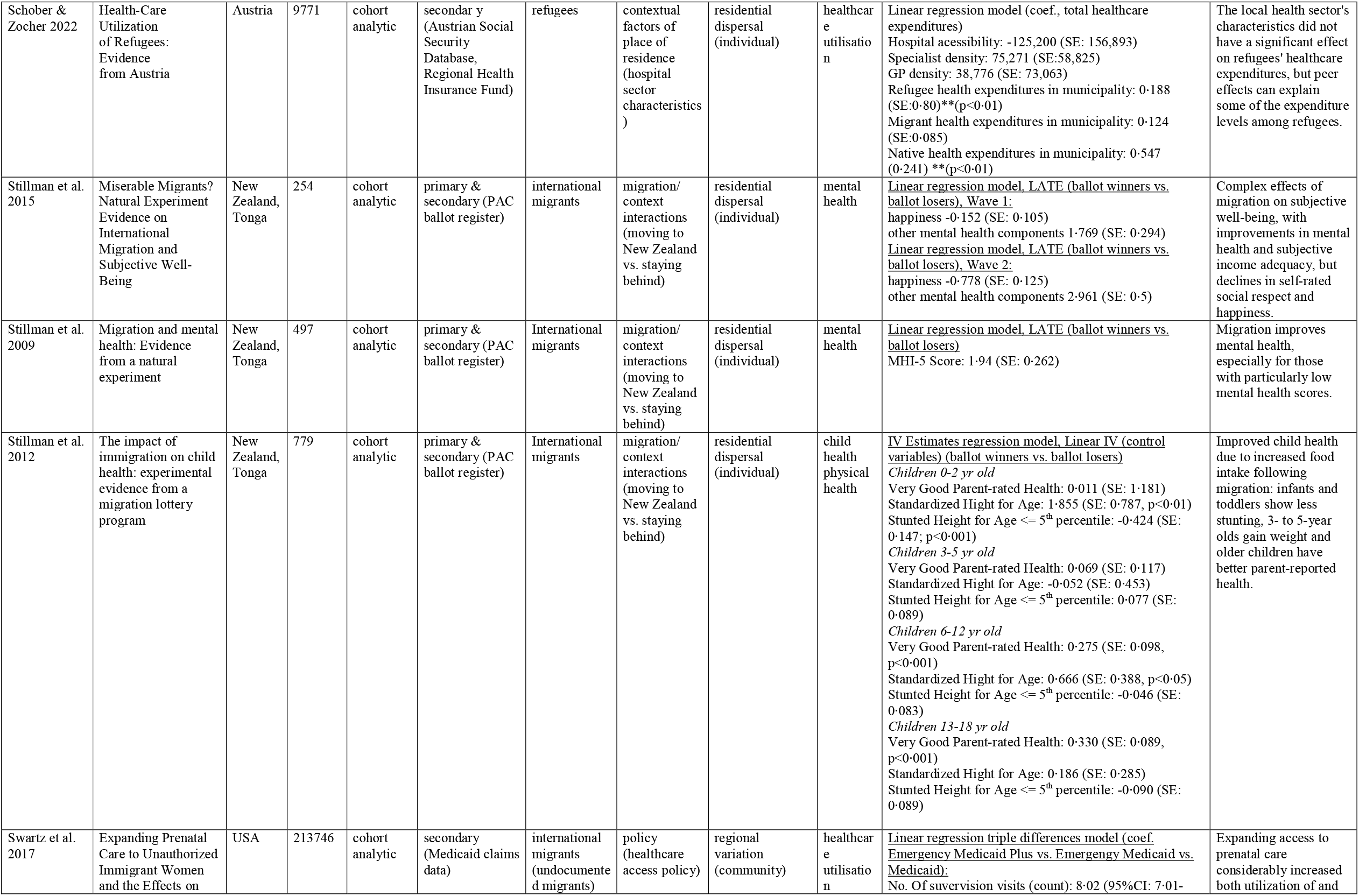
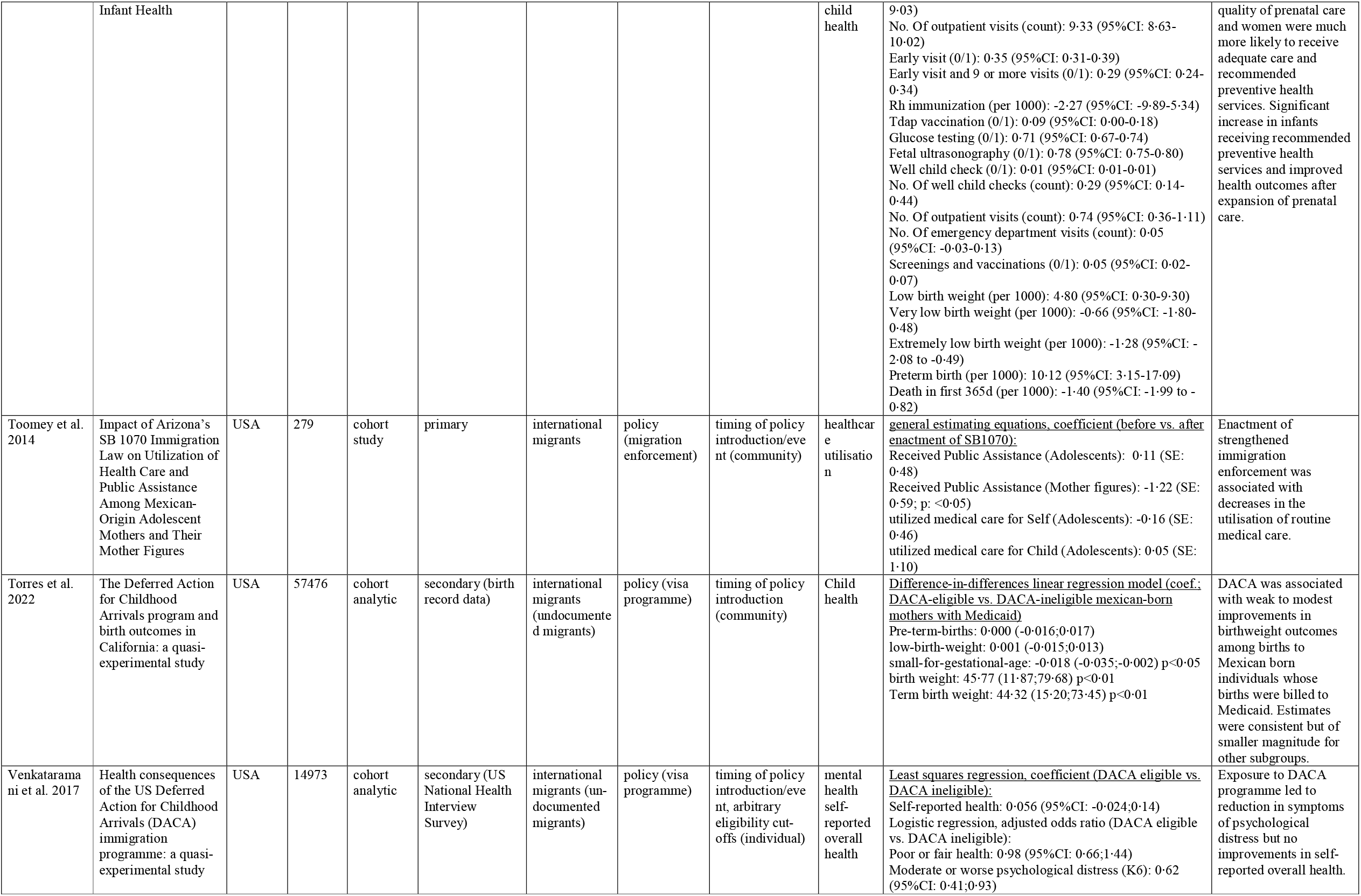
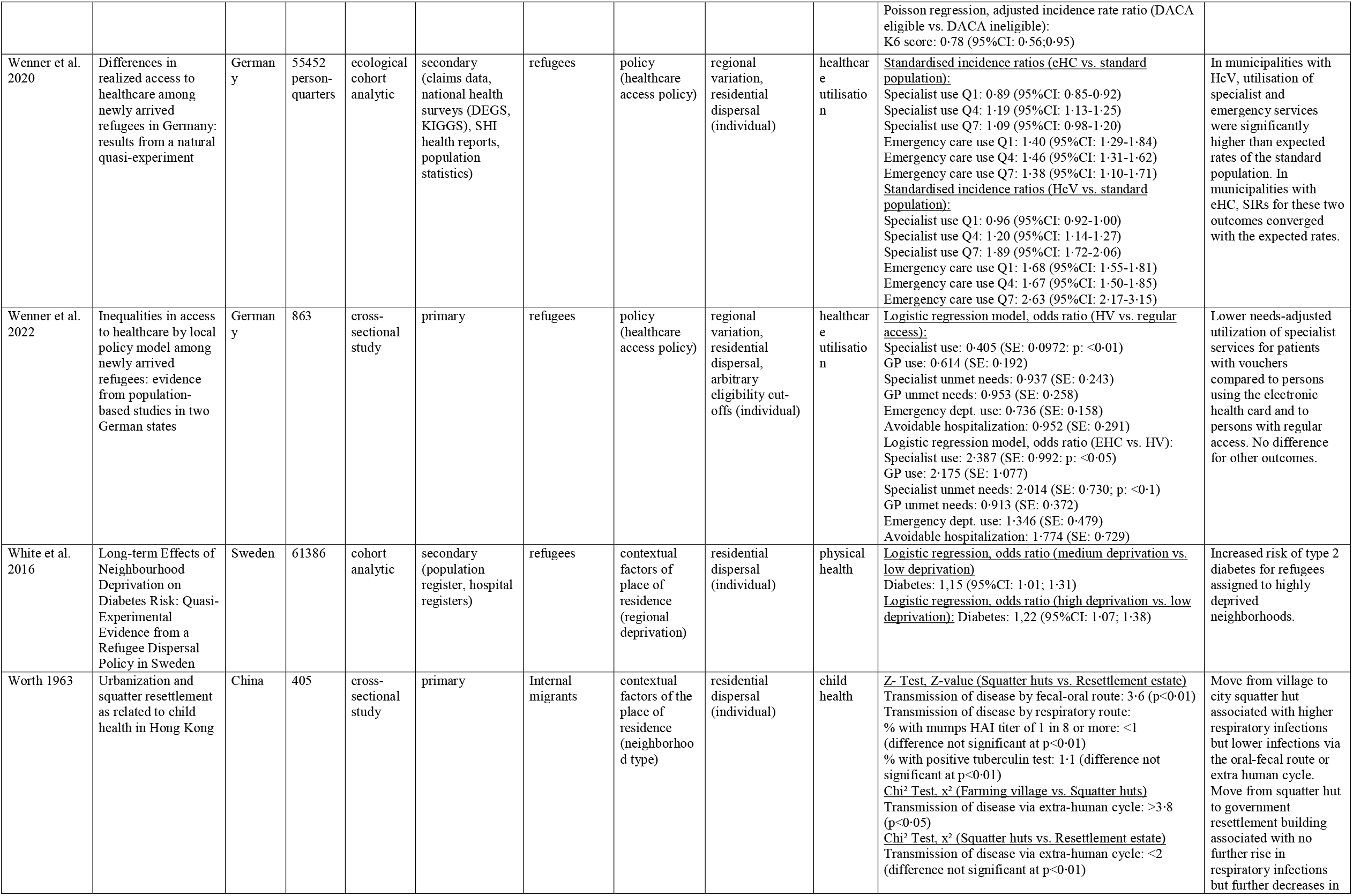
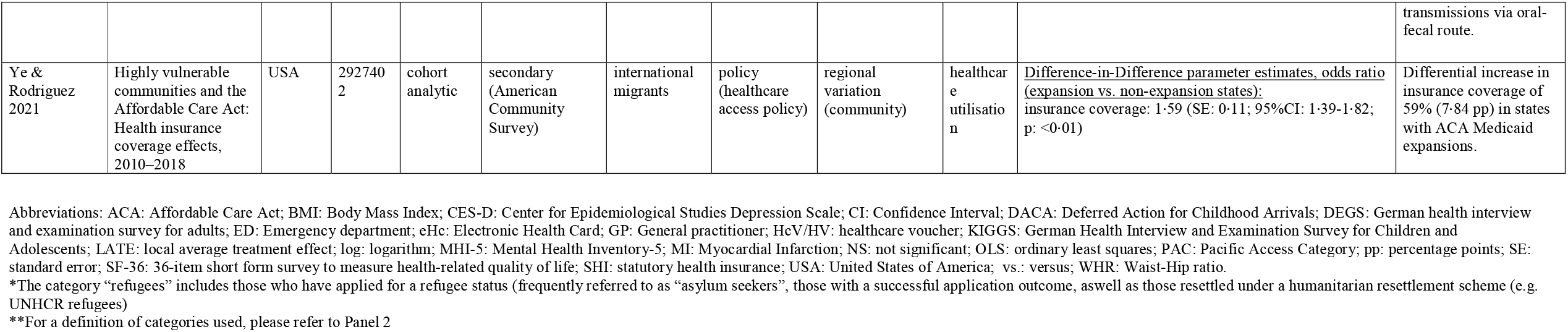
List of included studies and main study characteristics.

**Figure 2:**
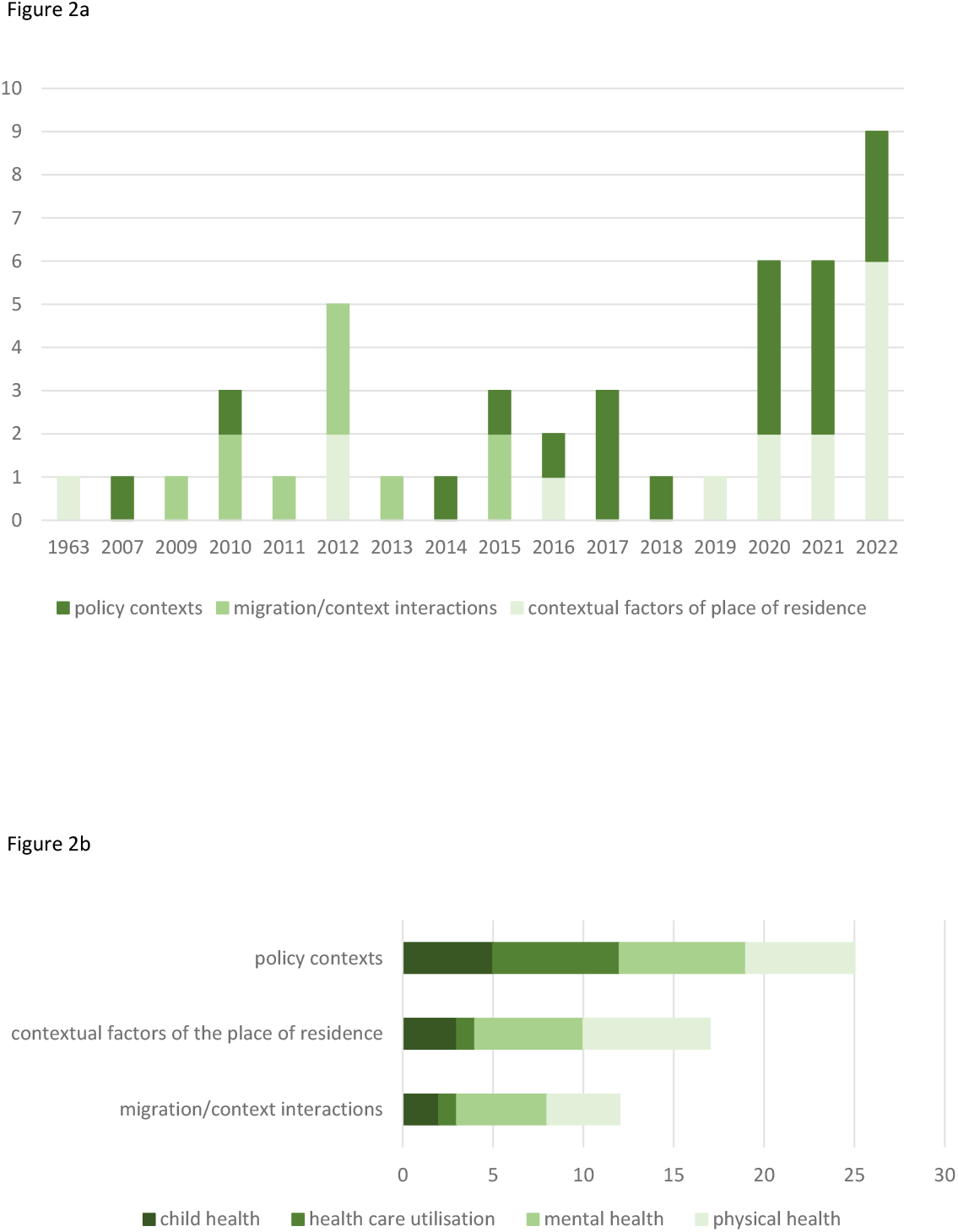
Overview of included studies by year of publication, main context factor groups and health outcome assessed. **2a: Included studies by year of publication and main context factor group** **2b: Included studies by main context factor group and health outcome assessed (totals surpass the total number of included studies as some studies assess multiple health outcomes)**

With respect to the source of variation in exposure underlying the natural experiments (see Panel 2), 17 studies make use of residential dispersal ^27,29–31,34,36,40–43,46,61,67–71^, 10 use regional variation in a policy implementation or natural event ^32,33,47–49,52,54,58,60,63^, seven studies make use of the timing of a policy or event ^26,37,39,53,55,56,64^ and two studies use the place of birth ^35,62^ (Table1). Nine studies combine two or more of these mechanisms in order to further strengthen the natural experiment design. For example, some studies ^38,44,45^ use a regional variation in exposure in combination with residential allocation mechanisms (e.g. in the context of a dispersed refugee population) in order to avoid bias through self-selection. Other studies ^28,51,57^ utilise quasi-random timing of policy implementation and an arbitrary eligibility cut-off to generate both contemporaneous and before/after comparison groups.

We found four types of contextual effects which were examined across studies: contextual factors of the place of residence in receiving countries (n=15) ^27,29–32,34,36,40–43,46,49,60,61^, migration-context interactions (n=10) ^59,62,63,65–71^, policy contexts (n=20) ^26,28,33,37–39,44,45,47,48,50–58,64^, as well as cultural factors as context (n=1) ^35^, the latter being an outlier in this set. Studies related to “contextual factors of the place of residence” examined specific regional characteristics such as regional deprivation, income inequality, neighbourhood disadvantage or migrant density among migrants in the resettlement phase. Studies focusing on “migration-context interactions” set out to examine the effect of migration on different health outcomes, but not by considering the process of migration itself, but rather by comparing outcomes among those who moved in comparison with a comparable group which did not migrate thus examining contextual characteristics of the new vs. old place of living. Finally, studies on policy contexts utilise a quasi-random uneven implementation of a policy, across time and/or space, to assess the effect of a given policy on health outcomes.

Just under half of studies (n=22) ^28,30–36,38,40–44,46,49,53,55,63–66^ use regression adjustment as their statistical analysis approach (Appendix Table S6). Other popular approaches include difference-in-difference analyses (n=10) ^39,47,48,51,52,54,56–59^, instrumental variables (n=8) ^27,60,62,67–71^ and regression discontinuity (n=3) ^26,37,50^. 17 studies ^27,28,30,31,33,37–39,45,47,48,50–52,54,57,62^ use the natural experiment as “intention to treat”, e.g. by program eligibility, rather than measuring actual treatment assignment. Given the potential of natural experiments to elucidate causal effects, all studies offer potential theoretical mechanisms through which this effect could occur. Less than half of studies (n=17) empirically assess such potential mediating pathways (Appendix Table S6).

Natural experiments on *physical health* provide strong evidence of a negative effect of contextual factors such as regional deprivation and neighbourhood disadvantage on diabetes^36,46^, heart disease^36^ and mortality^42^. An exception is the study by Grönqvist et al. (2012), which finds no effect of income inequality on mortality, days of sick leave or hospitalisation^34^. Living in an urban environment^29^ and increased social integration^60^ have positive effects on self-reported health (weak evidence). Studies examining policy contexts showed no effects or mixed effects on adult physical health, including studies on healthcare entitlements^38^ and visa access^57^ on self-rated health, as well as social benefits programmes on self-rated health and obesity^48,52^. One notable exception is the study by Juanmarti Mestres et al. (2020), which provides strong evidence for increased mortality due to restricting healthcare access policy^39^. Several natural experiments considering the effect of inclusionary policies on birth outcomes show consistent and strong positive effects; these include the effect of increased healthcare entitlements^54^ and increased visa access^51,56^. Natural experiments comparing migrants to non-migrants provide moderate evidence of the physical health disadvantages faced in post-migratory contexts, including effects on self-rated health, heart disease, obesity and mortality, considering a broad range of population groups: Tongan migrants to New Zealand^68^ Vietnamese refugees in the US^65^, internal migrants in the US^62^ and displaced persons in China^59^. Studies comparing Tongan children migrating to New Zealand compared to those that stayed behind show improvements in height-for age^67^ and stunting^69^.

With respect to *mental health*, the evidence on the effects of contextual factors of the place of residence is mixed (Figure 2). While some studies provide strong evidence of negative effects of regional deprivation^27^ and socioeconomic disadvantage^31^ on mental health, other high-quality studies find no such relationship^34,41^. Further studies find moderate evidence of the effect of the benefits of high migrant density^40^ and weak evidence of the negative effects of terrorist attacks^32^ on the mental health of migrants. Studies considering contextual factors of the place of residence on child health are also mixed, but one strong study demonstrates the negative effect of neighbourhood socioeconomic disadvantage on child mental health^30^. Another strong natural experiment looking at the composition of school classes shows a positive effect of increased proportions of foreign-born co-students on migrant mental health and health-risk behaviour, although this effect does not last into adulthood^49^. Studies assessing the effect of policy contexts on adult and child mental health find moderate but consistent evidence of the positive impacts of inclusionary migration, health and social policies^37,38,47,50,57^. The evidence on the mental health impacts of restrictive policies such as strengthened migration enforcement^53^, repatriation^64^ or reduced unemployment benefits^26^ is mixed. Natural experiments comparing migrants with individuals in the country of origin who did not migrate also provide mixed results with respect to mental health. While evidence from Tongan migrants in New Zealand^70,71^ and Vietnamese refugees in the US^66^ show positive or mixed results, natural experiments looking at the effects of displacement due to infrastructure projects^59^ or natural disasters^63^ show negative effects on mental health. One weak quality study found a positive effect of the life satisfaction in the country of origin on migrants life satisfaction after migration^35^.

Turning to *healthcare utilisation*, there is weak to moderate evidence for improvements when healthcare entitlements^44,45,54,58^ and access to social benefits are increased^48^. At the same time, there is evidence for decreases in timely utilisation when healthcare entitlements are restricted^28^ and migration enforcement is strengthened^55^. One moderate quality study found no effect of health sector characteristics on refugee healthcare expenditures^43^, while another weak quality study had mixed results regarding the healthcare accessibility of persons displaced by Hurricanes Kathrina and Rita^63^.

*Migrant groups can act as a lens* for the effects of context and health. All studies presented here include migrant groups as part of their analysis, but not all are migrant-specific. Based on their analytical perspective of migrants as a study population, we identified three groups of studies: 1) those that address a migrant-specific research question, 2) those who use natural experiments among migrants to address broader research questions on health and social environments, and 3) those who use migrants as a subgroup of a broader study population to test effect moderation of a given exposure among a marginalised group. Studies addressing migrant-specific research questions (n=29) ^28,30,31,38,39,43–45,49–51,53–57,59–71^ can be found among the migration/context interaction and policy contexts papers (Appendix Table S6). They address issues which are unique to the migration experience, such as the health effect of being removed from a known social and economic context into an unknown one (migration/ context interactions), or evaluate the health implications of migration and healthcare entitlement policies (policy contexts). As some of the studies point out, however, results are salient for other populations which are subject to similar restrictions on entitlements or benefits. 12 studies, nine of which consider contextual factors of the place of residence ^29,32,34–36,40–42,46^ and three of which consider social policies ^47,48,52^, use the (quasi-)random dispersal of migrants as an opportunity to answer broader questions of societal relevance on context and health, thereby extrapolating results beyond the migrant population. Finally, five studies ^26,27,33,37,58^ use migrant subgroups to examine the moderation of context effects among a well-defined, marginalised population group.

**Figure 3:**
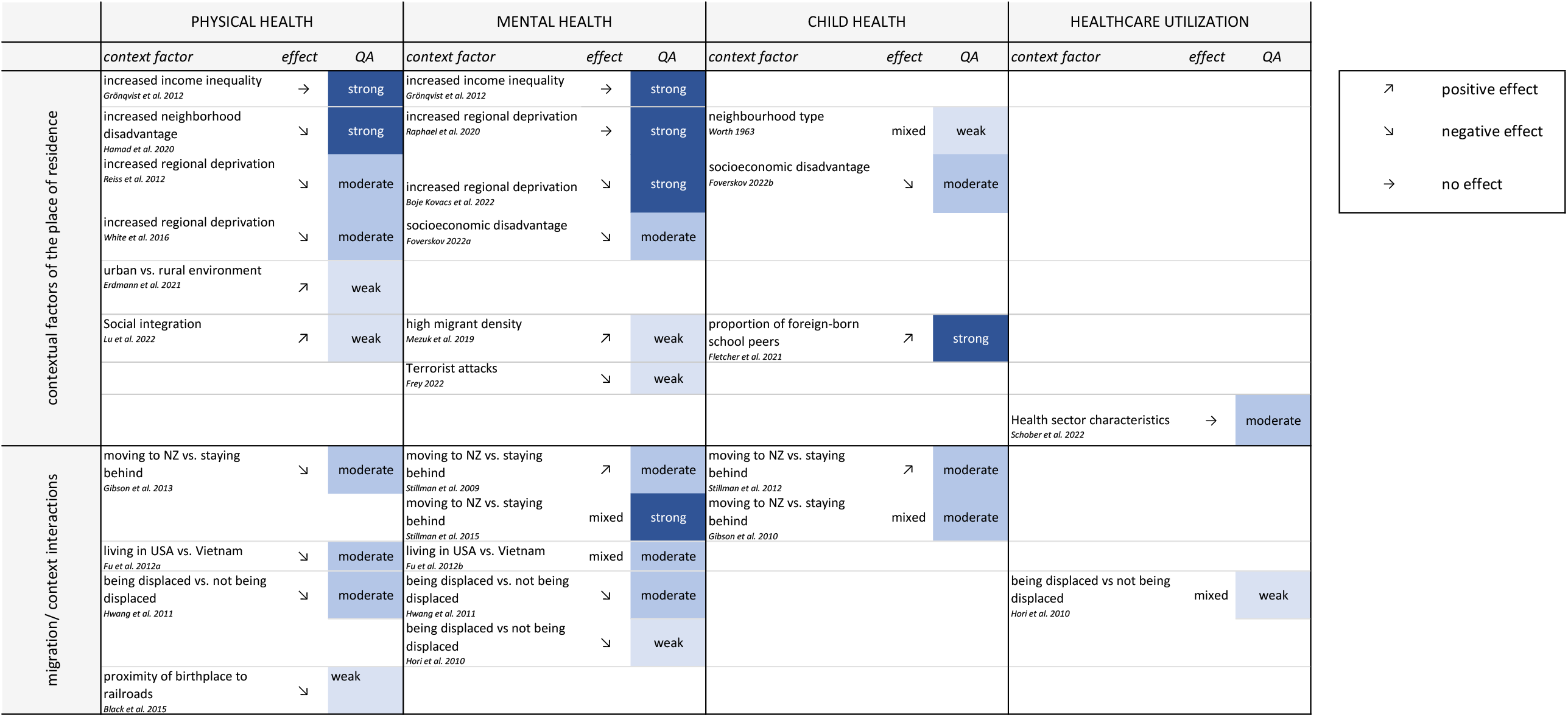
Evidence map illustrating results and quality of studies in the “contextual factors of the place of residence” and “migration/context interactions” groups. Abbreviations: NZ: New Zealand, QA: Quality Appraisal, USA: Unites States of America, vs.: versus

**Figure 4:**
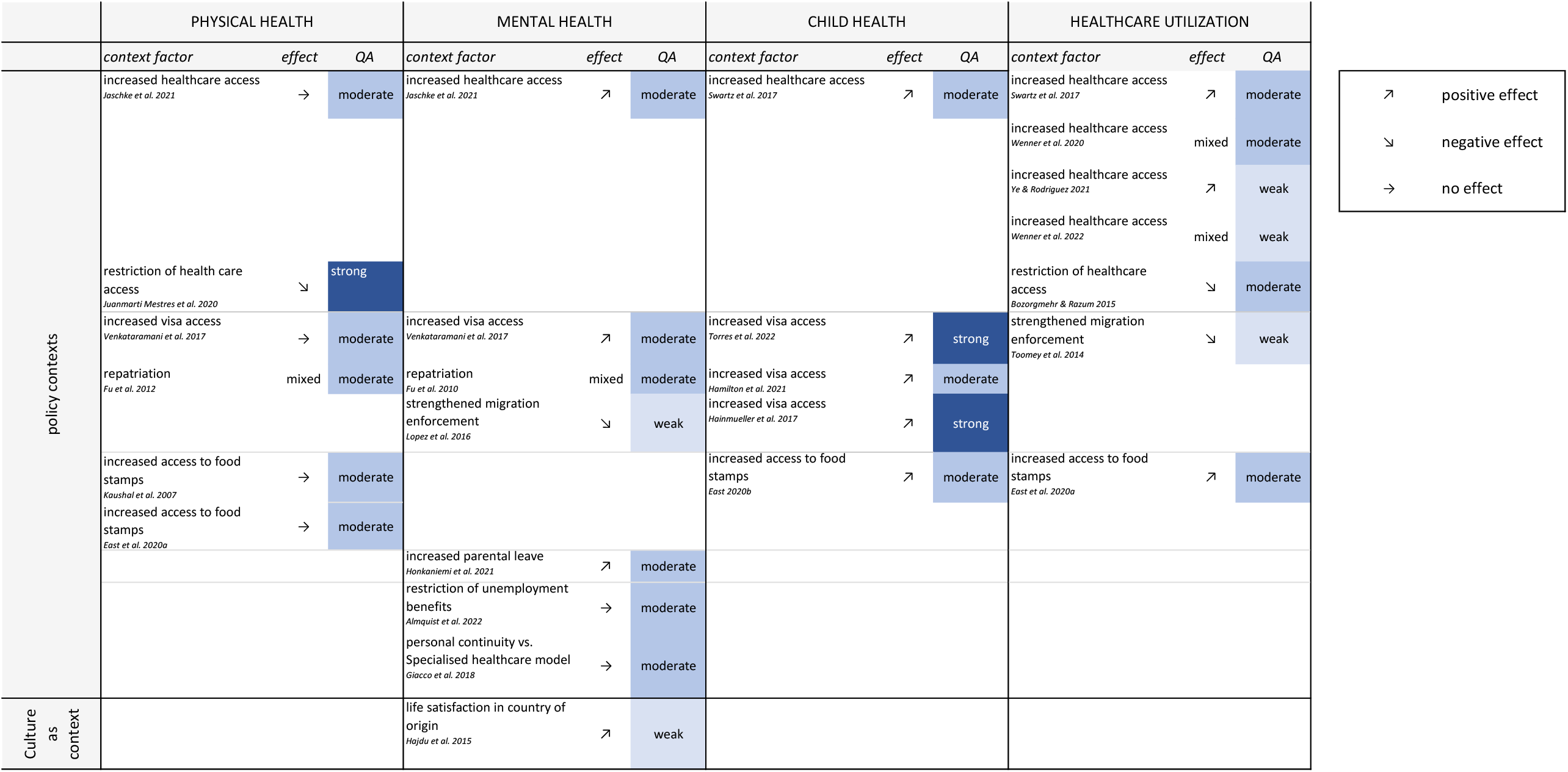
Evidence map illustrating results and quality of studies in the “policy contexts” and “culture as context” groups. Abbreviations: NZ: New Zealand, QA: Quality Appraisal, USA: Unites States of America, vs.: versus

## DISCUSSION

The evidence from the natural experiments included in this review confirms the negative effects of contextual factors such as socio-economic deprivation and neighbourhood disadvantage on physical health and mortality ^1^, while finding mixed effects of these neighbourhood attributes on mental health outcomes, suggesting that associations observed in the existing literature may be due to selection effects. Natural experiments comparing migrants with those that stayed behind are able to circumvent selective “healthy migrant effects”, and demonstrate the often detrimental effects of migration and post-migratory contexts on physical health and mortality. However, these studies also show that migrating to new social and economic contexts can be favourable for mental health and child health. Analyses of policy contexts indicate the negative impacts of restrictive migration and social policies on healthcare utilization, mental health and mortality as well as the positive effects when restrictions are lifted. These add to the emerging evidence base surrounding the effects of non-health targeted policies on migrant health ^10^ and the conflicting goals of migration and health policies ^21^.

Due to the nature of our research question and our search process, many of the included studies had migrant issues at the centre of their research objectives. They offered important insights into the detrimental effects of migration policies currently being broadly discussed and implemented, such the restriction of benefits for migrants and refugees ^28,38,39^, the restriction of visa programmes ^50,57^ and the increased enforcement of migration policies ^53,55,64^. Such policies emerge against the backdrop of wider social, political and economic developments which may be broadly described as “othering” processes. “Othering” refers to a discursive practice, in which an out-group is created which is marked as being different to reinforce and normalise the privileges of the antithetical “normal” in-group, disempowering and marginalising those considered to be on the “outside” in the process ^72^. Othering affects not just migrants, but also other marginalised populations, and may be conceptualised as the root of many pervasive health inequities. The benefit of specifically studying the effects of policies on migration is that they often make these underlying processes visible. However, the detrimental effects of benefit restrictions, tightened police control and accessibility hurdles described by the studies examined here can be applied to other marginalised populations for which these processes are less visible.

While the research questions of these studies and the policies examined are migration-specific, policy developments such as benefit restrictions and immigration raids emerge against a particular social and political backdrop which is not exclusive to migrants. Some studies also specifically use natural experiments among migrants to answer questions about the effect of contexts on the population more broadly. Given the diversity of refugee populations, and, increasingly, of so-called “host” populations in an era of super-diversity ^20^, there is no good reason to believe that the effects of context would operate any differently on migrant and non-migrant groups give careful consideration of causal models and mechanisms. We may, therefore, be able to use these studies as a lens and state that the identified negative effects of neighbourhood disadvantage and deprivation on physical health, mortality and child health ^27,30,31,36,41,42,46^ as well as the absence of effects of income inequality ^34^ apply to other human beings exposed to these contextual effects. This applies equally to other studies using natural experiments among migrants to study the effects of policy changes such as food stamp restrictions ^47,48,52^.

As described above, natural experiments among migrants can be powerful tools to yield insights on questions of contextual effects on health. The studies included in this review make use of a widely varying set of natural experiments, but this list is by no means exhaustive. Future research should continue to utilise the opportunities presented by natural experiments among migrants. This is sensible not just from a methodological standpoint, but also keeping the ethical use of research resources in mind. However, the included studies were highly heterogeneous in terms of not only the context factors and health outcomes considered, but also the methodological quality and detail of reporting. Given the “real life” complexity involved in the allocation to exposure, a highly detailed account of the nature of the natural experiments and the potential sources of bias is required ^18^, but was only provided by some studies. This also requires a more in-depth consideration of the sources of variation in exposure (see Panel 2) and to what extent these can be considered “successful” in terms of achieving random or quasi-random allocation. Of the mechanisms utilised in the studies included in this review, residential dispersal can be considered a particularly strong natural experiment because mechanisms of the dispersal are usually explicit and the true “randomness” of the allocation can be easily assessed. For all other sources of variation in exposure, the random nature of the allocation may be more implicit and requires the examination of a number of assumptions of the event or policy giving rise to the natural experiment. This requires in-depth knowledge of the political and institutional contexts in which the study takes place. Sources of variation in exposure which utilise allocation at a community level are further limited by the fact that these are often “intention to treat” studies. While the policy may apply at a community level, it is frequently not known whether individuals utilise/ are affected by the policy in question. Future research utilising natural experiment approaches should be more explicit in the mechanisms underlying the experiment, and may benefit from using our classification of the sources of variation in exposure (Panel 2).

Another methodological challenge is the consistent use of causal models to link exposure and outcome, as well as the deliberate use of mediation analyses to explore potential causal pathways. Very few studies provided details on potential causal mechanisms underlying the observed effects, so that these remain a black box. While many studies included covariates both at the individual and group/contextual level, the use of covariates was rarely justified and there was little recognition for the potential problems of introducing individual and group-level covariates in the same model. The use of directed acyclic graphs ^73^ and multi-level modelling approaches ^74^ should be considered to strengthen the quality of natural experiment approaches. Overall, methodological guidelines for natural experiments exist ^16^, but their application is not yet common practice. As research using natural experiments becomes increasingly popular, the use of such guidelines should be adopted by researchers and encouraged by funders and publishers. The scope and applicability of these guidelines may need to be revised for a more diverse set of research objectives, including research on contextual effects.

This study benefitted from a systematic review process, a search without a time limit in multiple databases, an additional snowballing search and an extensive data extraction process. The criterion of the natural experiment was applied at the full-text stage, meaning that we were able to select studies based on the full description of the methodology rather than relying on abstracts. The review was further enriched by the application of a rigorous quality appraisal tool which was adapted for use with natural experiments. A limitation was the use of “natural/quasi experiment” as an explicit search term, which may have excluded studies which used a natural experiment methodology but did not describe these as such. Finally, the review included a number of studies which applied econometric methodological approaches, but we did not have a researcher with an economic background as part of the research team.

Natural experiments can serve as powerful tools in disentangling the effect of context on health and reduce bias through self-selection. Results demonstrate the negative impacts for health brought about by neighbourhood disadvantage and restrictive policies. At the same time, studies uncover the potential of health and welfare programs to counteract the disadvantages created by othering processes and instead promote healthy (post-migratory) contexts. With careful consideration of causal pathways, results from migration contexts can serve as a useful magnifying glass for the effects of context for other population groups.

## Data Availability

A full list of studies identified in the search as well as the full data extracted from included studies are available for academic research projects by request to the corresponding author

## Contributors

LB: Conceptualization, Methodology, Investigation, Formal analysis, Writing – Original Draft, Visualization, Project administration

MH: Methodology, Investigation, Formal analysis, Data curation, Writing – Review & Editing, Visualization, Project administration

DC: Investigation, Formal analysis ZW: Investigation, Formal analysis

KB: Conceived the study, Conceptualization, Methodology, Formal analysis, Writing – Review & Editing, Supervision, Funding acquisition

All authors had full access to all data in the study and had final responsibility for the decision to submit for publication.

## Acknowledgments

This study was funded the German Science Foundation (DFG) in the scope of the NEXUS project, a part of the PH-LENS research consortium (FOR 2928 / GZ: BO 5233/1-1). We thank Judith Wenner for her support in the early stages of the screening process.

