## supplementary_material for "Context and health: a systematic review of natural experiments among migrant populations"

### Table S1: Full search terms for one database

#### Search conducted on the 4 Feb 2020

| Search | Search strategy PubMed | No. of records |
|---|---|---|
| 1 | ((((((((((((((Refugees[MeSH Terms]) OR Human Migration[MeSH Terms]) OR (Emigration and Immigration[MeSH Terms])) OR Asylum seek*) OR Migrant*) OR migrat*) OR Foreign*) OR immigra*) OR emigra*) OR internal migra*) OR IDP*)) AND (((((((Health Services[MeSH Terms]) OR Health Services Accessibility[MeSH Terms]) OR Quality of Health care[MeSH Terms]) OR Social Determinants of Health[MeSH Terms]) OR Health status[MeSH Terms]) OR Health*) OR healthcare)) AND (((((((((((dispers*) OR resettle*) OR transfer*) OR reloca*) OR distribut*)))) OR ((("natural experiment*") OR "quasi experiment*"))))) AND ((((((((((((Labour*[Other Term]) OR Work*[Other Term]) OR Occup*[Other Term]) OR Depriv*[Other Term]) OR Educa*[Other Term]) OR Income*[Other Term])) OR Hous*[Other Term]) OR Polic*[Other Term]) OR Context*[Other Term]) OR neighborhood[Other Term]) OR small-area[Other Term]))) AND (english[Filter] OR german[Filter])) | 1040 |
| 2 | ((((((((((((((Refugees[MeSH Terms]) OR (Human Migration[MeSH Terms])) OR (Emigration and immigration[MeSH Terms])) OR (Asylum seek*[Other Term])) OR (Migrant*[Other Term])) OR (Foreign*[Other Term])) OR (Migrat*[Other Term])) OR (non-citizen)) OR (immigra*[Other Term])) OR (emigra*[Other Term])) OR (internal migra*[Other Term])) OR (IDP*[Other Term]) AND (english[Filter] OR german[Filter])) AND (((((((("natural experiment*"[Other Term]) OR ("quasi experiment*"[Other Term])) OR (dispersal program*[Other Term])) OR (dispers*[Other Term])) OR (relocation[Other Term])) OR (tansfer*[Other Term])) OR (distribut*[Other Term]) AND (english[Filter] OR german[Filter]))) AND ((((((health services[MeSH Terms]) OR (health service accessability[MeSH Terms])) OR (quality of health care[MeSH Terms])) OR (health status[MeSH Terms])) OR (social determinants of health[MeSH Terms])) OR (health*[Other Term]) AND (english[Filter] OR german[Filter]))) AND (((((((((((Labour*[Other Term]) OR (Work*[Other Term])) OR (Occup*[Other Term])) OR (Income*[Other Term])) OR (Depriv*[Other Term])) OR (Educa*[Other Term])) OR (Hous*[Other Term])) OR (Polic*[Other Term])) OR (Context*[Other Term])) OR (neighbourhood[Other Term])) OR (small-area[Other Term]) AND (english[Filter] OR german[Filter])) | 344 |

#### Update Search conducted on October 13, 2022

| Search | Search strategy PubMed | No. of records |
|---|---|---|
| 1 | (("refugees"[MeSH Terms] OR "human migration"[MeSH Terms] OR "emigration and immigration"[MeSH Terms] OR (("asylum"[All Fields] OR "asylum s"[All Fields] OR "asylums"[All Fields]) AND "seek*"[All Fields]) OR "migrant*"[All Fields] OR "migrat*"[All Fields] OR "foreign*"[All Fields] OR "immigra*"[All Fields] OR "emigra*"[All Fields] OR (("internal"[All Fields] OR "internally"[All Fields] OR "internals"[All Fields]) AND "migra*"[All Fields]) OR "idp"[All Fields] OR "IDPs"[All Fields]* OR "displaced*"[All Fields]) AND ("health services"[MeSH Terms] OR "health services accessibility"[MeSH Terms] OR "quality of health care"[MeSH Terms] OR "social determinants of health"[MeSH Terms] OR "health status"[MeSH Terms] OR "health*"[All Fields] OR "delivery of health care"[MeSH Terms] OR ("delivery"[All Fields] AND "health"[All Fields] AND "care"[All Fields]) OR "delivery of health care"[All Fields] OR "healthcare"[All Fields] OR "healthcare*"[All Fields] OR "healthcares"[All Fields]) AND ("dispers*"[All Fields] OR "resettle*"[All Fields] OR "transfer*"[All Fields] OR "reloca*"[All Fields] OR "distribut*"[All Fields] OR "natural experiment*"[All Fields] OR "quasi experiment*"[All Fields]) AND ("labour*"[Other Term] OR "work*"[Other Term] OR "occup*"[Other Term] OR ("depriv*"[Other Term] OR "educa*"[Other Term] OR "income*"[Other Term] OR "hous*"[Other Term]) OR ("polic*"[Other Term] OR "context*"[Other Term] OR "neighborhood"[Other Term] OR "small-area"[Other Term]))) AND ((2020/2/4:2022/10/13:[pdat]) AND (english[Filter] OR german[Filter])) | 190 |
| 2 | (("refugees"[MeSH Terms] OR "human migration"[MeSH Terms] OR "emigration and immigration"[MeSH Terms] OR "asylum seek*"[Other Term] OR "migrant*"[Other Term] OR "foreign*"[Other Term] OR "migrat*"[Other Term] OR "non-citizen"[All Fields] OR "immigra*"[Other Term] OR "emigra*"[Other Term] OR "internal migra*"[Other Term] OR "IDP"[Other Term] OR "IDPs"[All Fields] OR "displace*"[All Fields]) AND ("natural experiment*"[Other Term] OR "quasi experiment*"[Other Term] OR (("dispersability"[All Fields] OR "dispersable"[All Fields] OR "dispersal"[All Fields] OR "dispersals"[All Fields] OR "dispersant"[All Fields] OR "dispersants"[All Fields] OR "disperse"[All Fields] OR "dispersed"[All Fields] OR "disperser"[All Fields] OR "dispersers"[All Fields] OR "disperses"[All Fields] OR "dispersibilities"[All Fields] OR "dispersibility"[All Fields] OR "dispersible"[All Fields] OR "dispersing"[All Fields] OR "dispersion"[All Fields] OR "dispersions"[All Fields] OR "dispersities"[All Fields] OR "dispersity"[All Fields] OR "dispersive"[All Fields] OR "dispersively"[All Fields] OR "dispersivities"[All Fields] OR "dispersivity"[All Fields]) AND "program*"[Other Term]) OR "dispers*"[Other Term] OR "relocation"[Other Term] OR "tansfer*"[Other Term] OR "distribut*"[Other Term]) AND ("health services"[MeSH Terms] OR "health services accessibility"[MeSH Terms] OR "quality of health care"[MeSH Terms] OR "health status"[MeSH Terms] OR "social determinants of health"[MeSH Terms] OR "health*"[Other Term]) AND ("labour*"[Other Term] OR "work*"[Other Term] OR "occup*"[Other Term] OR "income*"[Other Term] OR "depriv*"[Other Term] OR "educa*"[Other Term] OR "hous*"[Other Term] OR "polic*"[Other Term] OR "context*"[Other Term] OR "neighbourhood"[Other Term] OR "small-area"[Other Term])) AND (2020/2/4:2022/10/13:[pdat]) | 1 |

* "idp"[All Fields] OR "IDPs"[All Fields] added in update search

#### Table S2: Inclusion and Exclusion criteria.

| | Inclusion | Exclusion |
|---|---|---|
| **Population** | • Refers to migrants according to the definition of IOM (1)<br>"An umbrella term, not defined under international law, reflecting the common lay understanding of a person who moves away from his or her place of usual residence, whether within a country or across an international border, temporarily or permanently, and for a variety of reasons. The term includes a number of well-defined legal categories of people, such as migrant workers; persons whose particular types of movements are legally defined, such as smuggled migrants; as well as those whose status or means of movement are not specifically defined under international law, such as international students." | *Title- and Abstract-Screening:*<br>• Unclear population or no specific mention of migrant populations<br>*Additionally, in Full-text-Screening:*<br>• Results not reported separately for migrant population |
| **Phenomen of interest / Exposure / Outcome** | *Title- and Abstract-Screening:*<br>• Health outcomes reported<br>• Exposure of a contextual effect<br>*Additionally, in Full-text-Screening:*<br>• Research reporting evidence on contextual effects on health/ healthcare among migrant populations derived from a natural experiment. This includes studies reporting information on one or more of the following issues or aspects:<br>○ information about research design<br>○ analysis strategies<br>○ considered variables<br>○ Found effects | *Title- and Abstract-Screening:*<br>• No health outcomes<br>• No exposure of a contextual effect<br>• No quantitative research methods used<br>*Additionally, in Full-text-Screening:*<br>• Study did not employ natural experiment |
| **Setting** | • Any | • none |
| **Other** | • Studies in English or German | • Studies in other languages than English or German |

Note: Where necessary, divided into title- and abstract-screening and additional criteria for full-text-screening.

#### Table S3: List of excluded reports

| Authors | Year (published) | Title | Reason for exclusion | Notes |
|---|---|---|---|---|
| Honkaniemi H,Katikireddi SV,Rostila M,Juárez SP | 2022 | Psychiatric consequences of a father's leave policy by nativity: a quasi-experimental study in Sweden | Duplicate | |
| Gibson, John; McKenzie, David; Stillman, Steven | 2011 | Miserable Migrants? Natural Experiment Evidence on International Migration and Subjective Well-Being | Duplicate | |
| Ramachandran, Rajesh | 2012 | Language use in education and primary schooling attainment: Evidence from a natural experiment in Ethiopia.(Job Market Paper) | Duplicate | |
| Kammogne, Christiane Liliane; Marchand, Alain | 2019 | Les traits d'identité culturelle en lien avec le statut d'immigrant et l'ethnicité : quel lien avec les symptômes de détresse psychologique et les symptômes dépressifs dans la main-d'œuvre canadienne? Résultats des neuf cycles de l'ENSP | Language | Language other than German or English (French) |
| Javanbakht, Arash; Stenson, Anais; Nugent, Nicole; Smith, Alicia; Rosenberg, David; Jovanovic, Tanja | 2021 | Biological and Environmental Factors Affecting Risk and Resilience among Syrian Refugee Children | No contextual factors | |
| Denkinger, Jana Katharina; Rometsch, Caroline; Engelhardt, Martha; Windthorst, Petra; Graf, Johanna; Pham, Phuong; Gibbons, Niamh; Zipfel, Stephan; Junne, Florian | 2021 | Longitudinal Changes in Posttraumatic Stress Disorder After Resettlement Among Yazidi Female Refugees Exposed to Violence | No contextual factors | |
| Hahnefeld, Andrea; Sukale, Thorsten; Weigand, Elena; Muench, Katharina; Aberl, Sigrid; Eckler, Lea V.; Schmidt, Davin; Friedmann, Anna; Plener, Paul L.; Fegert, Joerg M.; Mall, Volker | 2021 | Survival states as indicators of learning performance and biological stress in refugee children: a cross-sectional study with a comparison group | No contextual factors | |
| Allem, J. P.; Ayers, J. W.; Irvin, V. L.; Hofstetter, C. R.; Hovell, M. F. | 2012 | South Korean Military Service Promotes Smoking: A Quasi-Experimental Design | No contextual factors | |
| Ljunge, Martin | 2018 | Trust promotes health: addressing reverse causality by studying children of immigrants | No contextual factors | |
| Cao, X. Y.; Chen, L.; Tian, L.; Jiang, X. L. | 2015 | Psychological Distress and Health-related Quality of Life in Relocated and Nonrelocated Older Survivors after the 2008 Sichuan Earthquake | No contextual factors | |
| Stillman, Steven; McKenzie, David; Gibson, John | 2007 | Migration and mental health: Evidence from a natural experiment | No contextual factors | |
| Haukka, Jari; Suvisaari, Jaana; Sarvimäki, Matti; Martikainen, Pekka | 2017 | The impact of forced migration on mortality | No contextual factors | |
| Wong, Y. Joel; Wang, Kenneth T.; Maffini, Cara S. | 2014 | Asian International Students' Mental Health-Related Outcomes: A Person x Context Cultural Framework | No contextual factors | |
| Steel, Zachary; Silove, Derrick; Chey, Tien; Bauman, Adrian; Phan, Tuong; Phan, T | 2005 | Mental disorders, disability and health service use amongst Vietnamese refugees and the host Australian population | No contextual factors | |
| Poole, Danielle N.; Hedt-Gauthier, Bethany; Liao, Shirley; Raymond, Nathaniel A.; Bärnighausen, Till | 2018 | Major depressive disorder prevalence and risk factors among Syrian asylum seekers in Greece | No contextual factors | |
| Kitabayashi, H.; Chiang, C.; Al-Shoaibi, A. A. A.; Hirakawa, Y.; Aoyama, A. | 2017 | Association Between Maternal and Child Health Handbook and Quality of Antenatal Care Services in Palestine | No contextual factors | |
| Lam, Kelvin KF; Johnston, Janice M | 2015 | Depression and health-seeking behaviour among migrant workers in Shenzhen | No contextual factors | |
| Feyera, F.; Mihretie, G.; Bedaso, A.; Gedle, D.; Kumera, G. | 2015 | Prevalence of depression and associated factors among Somali refugee at melkadida camp, southeast Ethiopia: a cross-sectional study | No contextual factors | |

| | | | |
|---|---|---|---|
| Abbas, Mohsin; Kashif, Muhammad; Balkhyour, Mansour; Ahmad, Ijaz; Asam, Zaki-Ul-Zaman; Saeed, Rashid | 2018 | Trends in occupational injuries and diseases among Saudi and non-Saudi insured workers | No contextual factors |
| Anderson, L. M.; Wood, D. L.; Sherbourne, C. D. | 1997 | Maternal acculturation and childhood immunization levels among children in Latino families in Los Angeles | No contextual factors |
| Baidoobonso, S.; Bauer, G. R.; Speechley, K. N.; Lawson, E.; Blacch Study Team | 2013 | HIV risk perception and distribution of HIV risk among African, Caribbean and other Black people in a Canadian city: mixed methods results from the BLACCH study | No contextual factors |
| Bates, L. M.; Acevedo-Garcia, D.; Alegria, M.; Krieger, N. | 2008 | Immigration and generational trends in body mass index and obesity in the United States: Results of the National Latino and Asian American survey, 2002-2003 | No contextual factors |
| Liang, Y.; Li, S. Q. | 2014 | Landless female peasants living in resettlement residential areas in China have poorer quality of life than males: results from a household study in the Yangtze River Delta region | No contextual factors |
| Horng, L.; Kakoly, N. S.; Abedin, J.; Luby, S. P. | 2019 | Effect of household relocation on child vaccination and health service utilisation in Dhaka, Bangladesh: a cross-sectional community survey | No contextual factors |
| Sadarangani, Tina R. | 2017 | Older Immigrants' Cardiovascular Risk Profiles: The Impact of Health Insurance | No contextual factors |
| Dayanand, K. K.; Punnath, K.; Chandrashekar, V.; Achur, R. N.; Kakkilaya, S. B.; Ghosh, S. K.; Kumari, S.; Gowda, D. C. | 2017 | Malaria prevalence in Mangaluru city area in the southwestern coastal region of India | No contextual factors |
| Habib, Rima R; Hojeij, Safa; Elzein, Kareem; Chaaban, Jad; Seyfert, Karin | 2014 | Associations between life conditions and multi-morbidity in marginalized populations: the case of Palestinian refugees | No contextual factors |
| Swartz, Jonas J.; Hainmueller, Jens; Lawrence, Duncan; Rodriguez, Maria I. | 2017 | Expanding Prenatal Care to Unauthorized Immigrant Women and the Effects on Infant Health | No contextual factors |
| Stillman, Steven; Gibson, John; McKenzie, David | 2012 | The impact of immigration on child health: experimental evidence from a migration lottery program | No contextual factors |
| Bauer, T. K.; Giesecke, M.; Janisch, L. M. | 2019 | The Impact of Forced Migration on Mortality: Evidence From German Pension Insurance Records | No contextual factors |
| Liu WN,He MZ,Dambach P,Schwertz R,Chen SM,Yu FY,Marx M | 2022 | Trends of overweight and obesity among preschool children from 2013 to 2018: a cross-sectional study in Rhine-Neckar County and the City of Heidelberg, Germany | No contextual factors |
| Petreski, Marjan | 2021 | Return migration and health outcomes in North Macedonia | No fulltext |
| Gulson, B. L.; Jameson, C. W.; Mahaffey, K. R.; Mizon, K. J.; Korsch, M. J.; Vimpani, G. | 1997 | Pregnancy increases mobilization of lead from maternal skeleton | No fulltext |
| Brittain, A. W. | 1991 | Anticipated child loss to migration and sustained high fertility in an east Caribbean population | No fulltext |
| Zhang, Shengling; Li, Yu; Zhang, Yipeng; Lu, Zhi-Nan; Hao, Yu | 2019 | Does sanitation infrastructure in rural areas affect migrant workers' health? Empirical evidence from China | No fulltext |
| Ali, Fatma Romeh M; Gurmu, Shiferaw | 2018 | The impact of female education on fertility: A natural experiment from Egypt | No fulltext |
| Gil-Salmerón, Alejandro; Valía-Cotanda, Elisa; Garcés-Ferrer, Jorge | 2018 | The effect of perceived discrimination on the health status of immigrant population in Spain Valencia | No fulltext |
| Eastwood, John | 2018 | Social Capital and Migrant Maternal Depression. A Multilevel Bayesian Latent Variable Spatial Logistic Regression in South Western Sydney, Australia | No fulltext |
| Abernethy, V. | 1994 | Population and women's health | No fulltext |

| | | | |
|---|---|---|---|
| Aragona, M.; Castaldo, M.; Cristina Tumiati, M.; Schilliro, C.; Dal Secco, A.; Agro, F.; Forese, A.; Tosi, M.; Baglio, G.; Mirisola, C. | 2019 | Influence of post-migration living difficulties on post-traumatic symptoms in Chinese asylum seekers resettled in Italy | No fulltext |
| Barbala, I. M.; Haug, H.; Grewal, N. K.; Eriksen, A.; Terragni, L. | 2018 | "Healthy Start" at asylum reception centres in Norway: nutrition education resources to be used with newly resettled asylum seekers | No fulltext |
| Gottvall, M.; Sjolund, S.; Arwidson, C.; Saboonchi, F. | 2019 | Health-related quality of life among Syrian refugees resettled in Sweden | No fulltext |
| Ferron, C.; HaourKnipe, M.; Tschumper, A.; Narring, F.; Michaud, P. A. | 1997 | Health behaviours and psychosocial adjustment of migrant adolescents in Switzerland | No fulltext |
| Geymen, A. | 2010 | USE OF GEOGRAPHICAL INFORMATION SYSTEMS IN EPIDEMIOLOGY: A CASE STUDY OF DILOVASI DISTRICT | No fulltext |
| Gusic, S.; Cardena, E.; Bengtsson, H.; Sondergaard, H. P. | 2017 | Dissociative Experiences and Trauma Exposure Among Newly Arrived and Settled Young War Refugees | No fulltext |
| Lai, D. W. L. | 2009 | Older Chinese' Attitudes Toward Aging and the Relationship to Mental Health: An International Comparison | No fulltext |
| Smith, P. M.; Mustard, C. A. | 2010 | The unequal distribution of occupational health and safety risks among immigrants to Canada compared to Canadian-born labour market participants: 1993-2005 | No fulltext |
| Nickerson, A.; Steel, Z.; Bryant, R.; Brooks, R.; Shove, D. | 2011 | Change in visa status amongst Mandaean refugees: Relationship to psychological symptoms and living difficulties | No fulltext |
| Rousseau, C.; Drapeau, A. | 2004 | Premigration exposure to political violence among independent immigrants and its association with emotional distress | No fulltext |
| Vaz, E.; Lee, K.; Moonilal, V.; Pereira, K. | 2018 | Potential of Geographic Information Systems for Refugee Crisis: Syrian Refugee Relocation in Urban Habitats | No fulltext |
| Wang, L. | 2014 | Immigrant health, socioeconomic factors and residential neighbourhood characteristics: A comparison of multiple ethnic groups in Canada | No fulltext |
| Wiseman, C. L. S.; Parnia, A.; Chakravartty, D.; Archbold, J.; Zawar, N.; Copes, R.; Cole, D. C. | 2017 | Blood cadmium concentrations and environmental exposure sources in newcomer South and East Asian women in the Greater Toronto Area, Canada | No fulltext |
| | 1982 | A comparative review of governments' views on objectives and policy instruments in the field of population and development | No fulltext |
| | 1982 | Assessment of ASEAN population programme | No fulltext |
| | 1983 | Meeting of the four Expert Groups convened in preparation for the International Conference on Population | No fulltext |
| | 1983 | Expert group meetings in preparation for the 1984 International Conference on Population | No fulltext |
| | 1985 | Algeria. United Nations. Department of International Economic and Social Affairs. Population Division. United Nations Fund for Population Activities UNFPA | No fulltext |
| | 1985 | Nepal (country/area statements) | No fulltext |
| | 1985 | Indonesia (country/area statements) | No fulltext |
| | 1985 | Hong Kong (country/area statements) | No fulltext |
| | 1985 | Malaysia (country/area statements) | No fulltext |
| | 1991 | Main figures from 10% sampling tabulation on China's 1990 Population Census | No fulltext |

| | | | |
|---|---|---|---|
| | 1993 | Introduction. Review of the six expert group meetings | No fulltext |
| | 1994 | Population dynamics in Asia and the Pacific: implications for development | No fulltext |
| | 1996 | All India Women's Conference Seminar on Habitat II and Human Settlement | No fulltext |
| | 1999 | HIV and refugees | No fulltext |
| Adewuyi, A. A. | 1986 | Interrelations between duration of residence and fertility in a Nigerian primate city | No fulltext |
| Akkerman, A. | 1977 | The household composition matrix and its application to migration analysis and population projection | No fulltext |
| Al-dakkak, M. S. | 1987 | The interaction between the legislative policy and the population problem in Egypt | No fulltext |
| Algeria, | 1989 | Presidential Decree No. 89-18 of 28 February 1989 relating to the publication in the Journal Officiel de la Republique Algerienne Democratique et Populaire of the constitutional revision adopted by referendum on 23 February 1989 | No fulltext |
| Allafi, S. | 1999 | First results of the microcensus 1998 | No fulltext |
| Arnauld, J. | 1983 | Urban nutrition: motor or brake for rural development? The Latin American case | No fulltext |
| Arp, W.; Baver, S. L. | 1994 | Implementation of Congressional intent: a study of amnesty policy and the Immigration and Naturalization Service | No fulltext |
| Barnabas, A. P. | 1974 | Population growth and social change: a note on rural society | No fulltext |
| Beaujot, R. P. | 1978 | Canada's population: growth and dualism | No fulltext |
| Bell, M. | 1982 | Botswana's way with self-help housing | No fulltext |
| Blanco Fdez De Valderrama, C. | 1993 | The new hosts: the case of Spain | No fulltext |
| Breznik, D. | 1989 | The population of Kosovo | No fulltext |
| Brockerhoff, M.; Yang, X. | 1994 | Impact of migration on fertility in sub-Saharan Africa | No fulltext |
| Canak, W. L. | 1985 | Republic of Colombia. Country Profile | No fulltext |
| | 1979 | Family planning for refugees | No fulltext |
| | 1979 | Governments urged to adopt population policies in line with national aims | No fulltext |
| | 1980 | Population growth and development planning in Africa | No fulltext |
| | 1981 | Italy: illegal construction hampers basic services | No fulltext |
| | 1981 | Bangladesh: planning intermediate growth centers | No fulltext |
| | 1983 | Brief on the Expert Group Meeting on Population Distribution, Migration and Development, Hammamet, Tunisia, 21-25 March 1983 | No fulltext |
| | 1984 | Bangladesh: population control versus health | No fulltext |
| | 1985 | Australia (country/area statements) | No fulltext |

| | | | |
|---|---|---|---|
| | 1987 | Attempts to achieve 1 per cent growth rate. Republic of Korea | No fulltext |
| | 1987 | Population policy | No fulltext |
| | 1989 | Family planning costs and benefits | No fulltext |
| | 1992 | Ethiopia: hard work for successful AIDS prevention | No fulltext |
| | 1993 | Florida teenagers learn about AIDS, teach others | No fulltext |
| | 1993 | Integrating AIDS components into the region's family planning programs | No fulltext |
| | 1993 | "PrepCom" ends with U.S. vow to expand pop programs; House panels act on UNFPA | No fulltext |
| Hefnawi, F. | 1983 | The role of research and training in family planning programmes | No fulltext |
| Hefnawi, F. I.; Ahmed, W. | 1982 | Population and human health | No fulltext |
| Alexandraki, C. | 1981 | Services rendered by ICM to migrant and refugee women | No fulltext |
| American, Demographics | 1987 | Latin America's supercity--the metropolitan area of Mexico City | No fulltext |
| American, Demographics | 1987 | The Calcutta metropolitan district | No fulltext |
| Anderson, J. M. | 1982 | An economic-demographic model of the United States labor market | No fulltext |
| Arab Population, Conference | 1994 | Arab Population Conference. Second Amman Declaration on Population and Development in the Arab World | No fulltext |
| Arcega, M. B. | 1977 | Survival of mankind: the population viewpoint | No fulltext |
| Arnold, F. S.; Shah, N. M. | 1984 | Asian labor migration to the Middle East | No fulltext |
| Globe International General Assembly | 1994 | Action agenda on population | No fulltext |
| Bautista, E. B. | 1986 | Reattraction of needed skills to developing countries of origin | No fulltext |
| Berentsen, W. H. | 1982 | Changing settlement patterns in the German Democratic Republic: 1945-1976 | No fulltext |
| Bhatterai, M. | 1994 | Using local resources to fight HIV / AIDS in Nepal | No fulltext |
| Bohning, W. R. | 1988 | The protection of migrant workers and international labour standards | No fulltext |
| Borthwick, P. | 1997 | The race to beat AIDS on the Lao-Thai border | No fulltext |
| Bulgaria, | 1989 | Regulation No. 44 of 11 September 1989 on the development of areas affected by migration | No fulltext |
| Burke, F. | 1991 | Shanghai housing reform. Program a burden on foreign investors | No fulltext |
| Carew, J. G. | 1981 | A note on women and agricultural technology in the Third World | No fulltext |
| Chakraborty, J.; Purohit, A.; Shah, S.; Kalla, S.; Purohit, S. | 1996 | A comparative study of the awareness and attitude of HIV / AIDS among students living in India and migrants to the United States | No fulltext |
| Chambers, R. | 1986 | Hidden losers? The impact of rural refugees and refugee programs on poorer hosts | No fulltext |

| | | | |
|---|---|---|---|
| Chang, M. C. | 1980 | Migration and fertility in Taiwan | No fulltext |
| Chant, S. | 1991 | Gender, migration and urban development in Costa Rica: the case of Guanacaste | No fulltext |
| Chasek, P.; Goree, L. J. th | 1993 | International Conference on Population and Development: year-end update | No fulltext |
| Chasteland, J. C. | 1987 | State of population research and research needs as expressed at the international conference on population and its preparatory meetings | No fulltext |
| Chen, C. | 1989 | Migration and transformation of industrial and occupational structures in Taiwan | No fulltext |
| Chow, C. S. | 1985 | American immigration policy, Chinese immigration, and Chinese concentration in New York City | No fulltext |
| Connell, J. | 1984 | Status or subjugation? women, migration and development in the South Pacific | No fulltext |
| Costa, Rica | 1987 | National Population Programme, Ministry of National Planning and Economic Policy, October 1987 | No fulltext |
| Crosswell, M. | 1981 | Growth, poverty alleviation and foreign assistance | No fulltext |
| D'Agnes, T.; D'Agnes, L. | 1982 | Community-based approach to refugee relief: experiences from Thailand | No fulltext |
| Debavalya, N. | 1981 | Population control, distribution, and manpower problems of Thailand | No fulltext |
| Dekker, R. | 1995 | Forced homecoming: Ghanaians' resettlement in their rural hometown. A case study | No fulltext |
| Diaz-briquets, S.; Perez, L. | 1981 | Cuba: the demography of revolution | No fulltext |
| Domros, M. | 1984 | India: population explosion and demographic transformation | No fulltext |
| Drakakis-smith, D. W. | 1984 | The changing economic role of women in the urbanization process: a preliminary report from Zimbabwe | No fulltext |
| Ecker, N. | 1998 | Where there is no village: teaching about sexuality in crisis situations | No fulltext |
| Eddings, J. | 1990 | As AIDS makes its way to S. Africa, citizens reluctant to heed warnings | No fulltext |
| Egypt, | 1987 | Second Five Year Plan for Socio-Economic Development, 1987/1988-1991/1992 | No fulltext |
| Ethiopia, | 1987 | Constitution, February 1987 | No fulltext |
| Fa'our, A. | 1981 | Migration from South Lebanon with a field study of forced mass migration | No fulltext |
| Farley, R. | 1986 | Assessing black progress: employment, occupation, earnings, income, poverty | No fulltext |
| Fuchs, R. J. | 1981 | Conflicts between explicit and implicit population distribution policies in Asian development plans | No fulltext |
| Naggan, L.; Forman, M. R.; Sarov, B.; Lewando-Hundt, G.; Zangwill, L.; Chang, D.; Berendes, H. W. | 1991 | The Bedouin Infant Feeding Study: study design and factors influencing the duration of breast feeding | No fulltext |
| Herrin, A. N.; Pardoko, H.; Lim, L. L.; Hongladorom, C. | 1981 | Demographic development in ASEAN: a comparative overview | No fulltext |
| Heywood, M. | 1996 | Mining industry enters a new era of AIDS prevention. Eye witness: South Africa | No fulltext |

| | | | |
|---|---|---|---|
| Holzer, J. Z. | 1984 | The final report from the head of the Keynote Problem No. 11.5. "Optimization of the demographic structures and processes in the Polish People's Republic" | No fulltext |
| Hoorweg, J.; Foeken, D.; Klaver, W.; Okello, W.; Veerman, W. | 1996 | Nutrition in agricultural development: land settlement in Coast Province, Kenya | No fulltext |
| Houstoun, M. F.; Kramer, R. G.; Barrett, J. M. | 1984 | Female predominance of immigration to the United States since 1930: a first look | No fulltext |
| Hu, S. Y.; Zhao, M. | 1997 | A study of the management mode of a rural migrant community | No fulltext |
| Hughes, H. | 1990 | Haiti. Beauty parlours and health promoters | No fulltext |
| Huntoon, L. | 1998 | Immigration to Spain: implications for a unified European Union immigration policy | No fulltext |
| Huq-hussain, S. | 1996 | Female migrants in an urban setting -- the dimensions of spatial / physical adaptation. The case of Dhaka | No fulltext |
| India, | 1989 | Extension of stay by foreigners, 1989 | No fulltext |
| Inserra, P. | 1985 | Ile de France: the metro area of Paris | No fulltext |
| Japan. Office of Statistical, Standards | 1982 | On our population movement: based on the result for 1% tabulation of 1980 population census | No fulltext |
| Jian, X. | 1996 | Increasing pressure: impacts of migration on cities | No fulltext |
| Johnnie, P. B. | 1988 | Rural-urban migration in Nigeria: consequences on housing, health-care and employment | No fulltext |
| Johnston, B. | 1982 | Somalia follow-up | No fulltext |
| Jones, G. W. | 1989 | Sub-national population policy: the case of North Sulawesi | No fulltext |
| Kamaci, M.; Mermut, S. | 1985 | Turkey: pressures on employment, housing, education and health care | No fulltext |
| Kandil, M.; Metwally, M. | 1992 | Determinants of the Egyptian labour migration | No fulltext |
| Kant, S. | 1993 | Urban development policy in India with special reference to Himachal Pradesh | No fulltext |
| Kawaguchi, P. T.; Nishi, S.; Schmitt, R. C. | 1981 | Population characteristics of Hawaii, 1980 | No fulltext |
| Kazakhstan, | 1993 | Constitution of the Republic of Kazakhstan [28 January 1993] | No fulltext |
| Khawaja, M. A.; Hockey, R. L. | 1979 | Sub-national differentials in New Zealand fertility, 1971-76 | No fulltext |
| Khor Geok, Lin | 1989 | Infant feeding practices in an urban squatter community | No fulltext |
| Khorev, B. | 1984 | Current scientific and practical problems in restricting the growth of large cities in the USSR | No fulltext |
| Kiani, M. F.; Siyal, H. B. | 1991 | Dimensions of urban growth in Pakistan | No fulltext |
| Klinger, A. | 1992 | The Germans in Hungary, 1941-1980 | No fulltext |
| Klugman, B. | 1991 | Population policy in South Africa: a critical perspective | No fulltext |
| Koll, R.; Vogler-ludwig, K. | 1993 | Effects of immigration to Bavaria on population structure, the labor and housing market, infrastructure, and land use | No fulltext |

| | | | |
|---|---|---|---|
| Kondrashin, A. V. | 1992 | Malaria in the WHO Southeast Asia region | No fulltext |
| Kornev, I. N. | 1983 | The demogeographic region as an object for planning and management | No fulltext |
| Kossoudji, S. A.; Ranney, S. I. | 1984 | The labor market experience of female migrants: the case of temporary Mexican migration to the U.S | No fulltext |
| Kotliar, A. | 1971 | Territorial problems relating to the structure of employment in terms of sex | No fulltext |
| Kvasha, A. | 1984 | Theoretical problems of demographic policy in the USSR | No fulltext |
| Latin, America; Caribbean, Population; Development, Conference | 1994 | Latin American and Caribbean regional conference on population and development. Latin American and Caribbean Consensus on Population and Development | No fulltext |
| Lee, R. | 1983 | Economic consequences of population size, structure and growth | No fulltext |
| Lepore, S. | 1986 | Problems confronting migrants and members of their families when they return to their country of origin | No fulltext |
| Li, B.; Zhang, W. | 1998 | Trend of population aging in China and the strategy | No fulltext |
| Li, H.; Wu, M.; Zhu, J.; Wu, G. | 1990 | A research on the moderate transference of China's agricultural labor | No fulltext |
| Mahmood, R. A. | 1995 | Emigration dynamics in Bangladesh | No fulltext |
| McCutcheon, L. | 1978 | Occupation and housing adjustment of migrants to Surabaya, Indonesia: the case of a second city | No fulltext |
| McDonald, P. | 1989 | Ethnic family structure | No fulltext |
| McKenzie, N. F. | 1990 | AIDS at the beginning of the second decade | No fulltext |
| McMurray, C. | 1992 | Issues in population planning: the case of Papua New Guinea | No fulltext |
| Mehta, S. | 1996 | Geography and migration policies: the Indian experience | No fulltext |
| Merrick, T. W. | 1991 | Population pressures in Latin America. [Updated reprint] | No fulltext |
| Mexico, | 1989 | National Population Programme 1989-1994. [Summary] | No fulltext |
| Mia, A. | 1973 | A population programme is more than family planning | No fulltext |
| Minkin, S. | 1979 | Bangladesh: where there's a pill there's a way | No fulltext |
| Mlay, Wfi | 1984 | Agricultural development and population change in the context of settlement patterns in Tanzania | No fulltext |
| Mohammad, R. | 1998 | FORWARD against female genital mutilation | No fulltext |
| Moodie, R.; Aboagye-Kwarteng, T. | 1993 | Confronting the HIV epidemic in Asia and the Pacific: developing successful strategies to minimize the spread of HIV infection | No fulltext |
| Morgan, J. | 1994 | Sudanese refugees in Koboko: environmental health interventions | No fulltext |
| Mosse, J. C. | 1994 | From family planning and maternal and child health to reproductive health | No fulltext |
| Tabbarah, R. | 1988 | Challenges in Arab demography | No fulltext |
| Tabbarah, R.; Mamish, M. A.; Gemayel, Y. | 1978 | Population research and research gaps in the Arab countries | No fulltext |

| | | | |
|---|---|---|---|
| Tao, L. | 1986 | Developing undertakings for the aged of China | No fulltext |
| Tarver, J. | 1985 | Republic of Botswana. Country profile | No fulltext |
| Teitelbaum, M. S. | 1992 | The population threat | No fulltext |
| Teller, C.; Sibrian, R.; Talavera, C.; Bent, V.; Del Canto, J.; Saenz, L. | 1979 | Population and nutrition; implications of sociodemographic trends and differentials for food and nutrition policy in Central America and Panama | No fulltext |
| Telleria, T. | 1998 | Peer education in Portugal. Adolescent health / sex education | No fulltext |
| Teo Cheok Chin, P. | 1989 | The relationship between poverty and fertility in Peninsular Malaysia: a district analysis | No fulltext |
| Thorndike, T. | 1988 | Netherlands Antilles: country profile | No fulltext |
| Torki, F. G. | 1984 | Occupational mobility of primary male migrants to urban areas in Egypt | No fulltext |
| Tuntawiroon, N.; Samootsakorn, P. | 1984 | Bangkok--a city ready to burst | No fulltext |
| Olivar, G. B. | 1978 | A look at Philippine population in the year 2000 | No fulltext |
| Rowe, L. | 1994 | Training tribal leaders in Thailand | No fulltext |
| Sayed, H. A. | 1984 | Community and family planning: a statistical analysis of Egyptian data | No fulltext |
| Scholz, U. | 1992 | Transmigration--a disaster? Problems and prospects of the Indonesian resettlement program | No fulltext |
| Schubert, J. | 1999 | Educating the community | No fulltext |
| Serra-vega, J. | 1990 | Andean settlers rush for Amazonia | No fulltext |
| Share, M. A. | 1987 | Wage differentials in Jordan: effects on integrated labour market | No fulltext |
| Simons, J. | 1973 | The development of population policy in Britain | No fulltext |
| Singh, S. N.; Sharma, H. I. | 1984 | A study of pattern of out-migration: a regression analysis | No fulltext |
| Singh, S. N.; Singh, V. K.; Burman, D. | 1985 | A continuous time model for first birth in rural environment of India | No fulltext |
| Soehartadji, | 1980 | Urbanization in Central Java and Yogyakarta | No fulltext |
| United Nations. Department of International, Economic; Social, Affairs; United Nations. Fund for Population Activities, Unfpa | 1981 | Chile | No fulltext |
| United Nations. Department of International, Economic; Social Affairs Population, Division; United Nations. Fund for Population Activities, Unfpa | 1982 | Egypt | No fulltext |
| United Nations. Department of International, Economic; Social Affairs. Population, Division | 1984 | Population distribution, migration and development: main issues for the 1980s | No fulltext |
| United Nations. Department of International, Economic; Social Affairs. Population, Division; United Nations Fund for Population Activities, Unfpa | 1980 | Ecuador | No fulltext |
| United Nations. Department of International, Economic; Social Affairs. Population, Division; United Nations Fund for Population Activities, Unfpa | 1981 | Colombia | No fulltext |

| | | | |
|---|---|---|---|
| United Nations. Department of International, Economic; Social Affairs. Population, Division; United Nations Fund for Population Activities, Unfpa | 1982 | Jamaica | No fulltext |
| United Nations. Department of International, Economic; Social Affairs. Population, Division; United Nations. Fund for Population Activities, Unfpa | 1981 | China | No fulltext |
| United Nations. Department of International, Economic; Social Affairs. Population, Division; United Nations. Fund for Populaton Activiteis, Unfpa | 1983 | Brazil | No fulltext |
| United States. Bureau of the, Census | 1980 | Population Profile of the United States: 1979 | No fulltext |
| United States. Bureau of the, Census | 1987 | America's centenarians: data from the 1980 census | No fulltext |
| United States. Department of State. Bureau of Public, Affairs | 1985 | Portugal | No fulltext |
| United States. Department of State. Bureau of Public, Affairs | 1985 | Mozambique | No fulltext |
| United States. Department of State. Bureau of Public, Affairs | 1987 | Barbados | No fulltext |
| United States. Department of State. Bureau of Public, Affairs | 1988 | Algeria | No fulltext |
| United States. Department of State. Bureau of Public, Affairs | 1989 | Australia | No fulltext |
| United States. Department of State. Bureau of Public Affairs. Office of Public, Communication | 1992 | Pakistan | No fulltext |
| United States. Department of State. Bureau of Public Affairs. Office of Public, Communication | 1992 | Chile | No fulltext |
| United States. Department of State. Bureau of Public Affairs. Office of Public, Communication | 1992 | Tanzania | No fulltext |
| Utomo, B.; Alimoeso, S.; Park, C. B. | 1983 | Factors affecting the use and non use of contraception | No fulltext |
| Valtonen, K. | 1996 | Bread and tea: a study of the integration of low-income immigrants from other Caribbean territories into Trinidad | No fulltext |
| Vanderkamp, J. | 1986 | On testing the human capital model of migration | No fulltext |
| Visaria, P.; Visaria, L. | 1981 | India's population: second and growing | No fulltext |
| Vithal, C. P. | 1992 | Socio-economic transformation of a primitive tribal group: a study of Chenchus in Andhra Pradesh | No fulltext |
| Von Bethlenfalvy, P. | 1987 | The problem of unemployment amongst refugees in Europe | No fulltext |
| Voss, P. R.; Rank, M. | 1980 | Characteristics of recent migrants to nonmetropolitan Northern Wisconsin | No fulltext |
| Wagner, M. | 1987 | Education and migration | No fulltext |
| Wang, Y. | 1990 | An analysis of changes in Chinese migrants' income | No fulltext |
| Wei, J. | 1990 | Trends in population growth in China's towns during the eighties, and town population in-migration and its decisive factors: a historic convergence of two types of demographic change | No fulltext |
| Wei, Y. | 1994 | Urban policy, economic policy, and the growth of large cities in China | No fulltext |

| | | | |
|---|---|---|---|
| Wery, R.; Rodgers, G. B.; Hopkins, M. J. | 1976 | On the evaluation of population and employment policy | No fulltext |
| Wheeler, J. | 1992 | People, poverty and the Earth Summit | No fulltext |
| Williams, B.; Campbell, C. | 1999 | Community mobilization as an HIV prevention strategy: challenges and obstacles (South Africa) | No fulltext |
| Wilson, J. D. | 1982 | Optimal income taxation and migration: a world welfare point of view | No fulltext |
| Wurzberger, P.; Wedel, E. | 1988 | First results of the population census, 1987 | No fulltext |
| Xie, J.; Yu, J. | 1993 | A comparative study on the population urbanized under or outside state plan: different status of mobile urban population and permanent new urban residents in the urbanization of the Chinese population according to the Fourth Census in China | No fulltext |
| Xie, Shenghua | 2017 | The Migration, Mental Stress, and Tobacco use of Internal Migrants in China: The Moderating Effect of the Social Context of the Host Society | No fulltext |
| Yadava, Kns; Singh, Srj | 1983 | A model for the number of rural out-migrants at household level | No fulltext |
| Yadava, K. N. | 1987 | Determinants of rural-urban migration in India: a micro approach | No fulltext |
| Yadava, K. N.; Singh, S. K.; Kumar, U. | 1989 | A probability model for the number of migrants | No fulltext |
| Yaser, Y. | 1990 | Family planning for seasonal migrant workers in Adana Province, Turkey | No fulltext |
| Yusof, K.; Zulkifli, S. N. | 1985 | Demographic and fertility characteristics of 4 squatter settlements | No fulltext |
| Zhang, K. | 1985 | Population growth in Shanghai and its characteristics | No fulltext |
| Zhu, J. | 1996 | An analysis of the social space structure of population in the Shanghai municipality | No fulltext |
| Zhu, W. | 1992 | Pilot project on the resettlement of out-migrant agricultural population in Yangtze Gorges Reservoir Area | No fulltext |
| Adams Jr, Richard H | 2011 | Evaluating the economic impact of international remittances on developing countries using household surveys: A literature review | No fulltext |
| Agesa, Richard U; Kim, Sunwoong | 2001 | Rural to urban migration as a household decision: Evidence from Kenya | No fulltext |
| Ben-Shlomo, Yoav; Mamluk, Loubaba; Redwood, Sabi | 2019 | A life-course perspective on migrant health | No fulltext |
| Boyle, Paul; Norman, Paul | 2009 | Migration and health | No fulltext |
| Brown, Philip | 2019 | Migration and asylum | No fulltext |
| Budnik, Katarzyna B | 2009 | 6. Polish emigration to the UK after EU enlargement in 2004: a 'natural experiment' for testing the rationality of migration choice1 | No fulltext |
| Kenny, Peter; Ressi, G; Wagner, AM Miörner; Kenny, PA | 2010 | Track 4: Workshop: Cooperation between different actors regarding promotion of return to work of sickness absentees | No fulltext |
| Liebig, Thomas; Puhani, Patrick A; Sousa-• Poza, Alfonso | 2007 | Taxation and Internal Migration - Evidence from the Swiss Census Using Community-• Level Variation in Income Tax Rates | No fulltext |
| Lindert, Jutta; Schinina, Guglielmo | 2011 | Mental health of refugees and asylum-seekers | No fulltext |
| Little, Michael A; Garruto, Ralph M | 2007 | Global impacts of anthropogenic climate change on human health and adaptability | No fulltext |

| | | | |
|---|---|---|---|
| Liu, Elaine M | 2012 | Does "in utero"• Exposure to Illness Matter? The 1918 Influenza Epidemic in Taiwan as a Natural Experiment Ming-Jen Lin National Taiwan University | No fulltext |
| Chen, Shuai; Oliva, Paulina; Zhang, Peng | 2018 | Air pollution and mental health: evidence from China | No fulltext |
| Chen, Yi; Jiang, Sheng; Zhou, Li-An | 2019 | Estimating returns to education in urban China: Evidence from a natural experiment in schooling reform | No fulltext |
| Clemens, Michael A | 2005 | A first look at the consequences of African health professional emigration | No fulltext |
| De Silva, Dakshina G; McComb, Robert P; Moh, Young-Kyu; Schiller, Anita R; Vargas, Andres J | 2009 | Migration and Wages: A Natural Experiment from Hurricane Katrina | No fulltext |
| Gammage, Sarah; Stevanovic, Natacha | 2019 | Gender, migration and care deficits: what role for the sustainable development goals? | No fulltext |
| Gaynor, Marty; Propper, Carol | 2012 | Healthcare competition saves lives | No fulltext |
| Giacco, Domenico; Bird, VJ; Ahmad, T; Bauer, M; Lasalvia, A; Lorant, V; Miglietta, E; Moskalewicz, J; Nicaise, P; Pfennig, A | 2019 | The same or different psychiatrists for in-and out-patient treatment? A multi-country natural experiment-ADDENDUM | No fulltext |
| Gibson, John; McKenzie, David; Rohorua, Halahingano; Stillman, Steven | 2015 | Beliefs, Preferences and Migration: Evidence from Combining Lab-in-Field and Natural Experiments | No fulltext |
| Gibson, John; McKenzie, David; Rohorua, Halahingano; Stillman, Steven | 2018 | The Long-term impacts of international migration: Evidence from a lottery | No fulltext |
| Hall, Matthew; Graefe, Deborah Roempke; De Jong, Gordon F | 2010 | Economic self-sufficiency among immigrant TANF-leavers: Welfare eligibility as a natural experiment | No fulltext |
| Hyll, Walter; Schneider, Lutz | 2018 | Income comparisons and attitudes towards foreigners-Evidence from a natural experiment | No fulltext |
| Jakiela, Pamela; Ozier, Owen | 2019 | The impact of violence on individual risk preferences: evidence from a natural experiment | No fulltext |
| Jordan, Bill; Brown, Philip | 2007 | Migration and work in the United Kingdom: Mobility and the social order | No fulltext |
| Lozano, Fernando A | 2007 | Public Policy and Immigrant Settlement | No fulltext |
| Nauman, Elizabeth; VanLandingham, Mark; Anglewicz, Philip | 2016 | Migration, urbanization and health | No fulltext |
| Oh, Yoon Ah | 2014 | Life satisfaction of the families of migrants in the Philippines | No fulltext |
| Panda, Shilpi Smita; Mishra, Nihar Ranjan | 2018 | Factors affecting temporary labour migration for seasonal work: a review | No fulltext |
| Panos, Patrick T; Panos, Angelea Journot | 2000 | A model for a culture-sensitive assessment of patients in health care settings | No fulltext |
| Polachek, Solomon W; Tatsiramos, Konstantinos | 2019 | Health and Labor Markets | No fulltext |
| Roach, Kent | 2006 | The post-9/11 migration of Britain's Terrorism Act 2000 | No fulltext |
| Schofield, Peter | 2019 | Why urban environments matter for refugee mental health | No fulltext |
| Snodgrass, Jeffrey G; Upadhyay, Chakrapani; Debnath, Debashis; Lacy, Michael G | 2016 | World Development Perspectives | No fulltext |
| Sotomayor, Orlando | 2013 | Fetal and infant origins of diabetes and ill health: Evidence from Puerto Rico's 1928 and 1932 hurricanes | No fulltext |
| | 1993 | Expert Group Meeting on Population Policies and Programmes | No fulltext |
| | 1994 | Aid agencies urged -- more family planning | No fulltext |

| | | | |
|---|---|---|---|
| | 1995 | Commission on Population and Development advises on follow-up to ICPD | No fulltext |
| | 1996 | Breastfeeding and food security | No fulltext |
| | 1996 | Population issues surface at human settlements conference | No fulltext |
| | 1998 | NGO action on refugees and displaced people | No fulltext |
| | 1998 | Condom use soars after intervention, China | No fulltext |
| Acharya, S. | 1998 | Rethinking about HIV infection and AIDS | No fulltext |
| Addo, N. O. | 1972 | Employment and labor supply on Ghana's cocoa farms in the pre- and post-aliens compliance order era | No fulltext |
| Gondowarsito, R. | 1990 | Transmigrasi Bedol Desa: inter-island village resettlement from Wonogiri to Bengkulu | No fulltext |
| Greenspan, A. | 1992 | Fertility decline in Bangladesh: an emerging family planning success story | No fulltext |
| Hakkert, R. | 1986 | Metropolitan Lima: area profile | No fulltext |
| Hakkert, R. | 1986 | Republic of Italy (country profile) | No fulltext |
| Hamilton, B.; Whalley, J. | 1984 | Efficiency and distributional implications of global restrictions on labour mobility: calculations and policy implications | No fulltext |
| Hansen, K. A. | 1987 | Geographical mobility: 1985 | No fulltext |
| Hansen, K. A. | 1988 | Geographical mobility: March 1985 to March 1986 | No fulltext |
| Hansen, K. A.; Boertlein, C. G. | 1984 | Geographical mobility: March 1981 to March 1982 | No fulltext |
| Hardjono, J. | 1986 | Transmigration: looking to the future | No fulltext |
| Harkavy, O. | 1973 | The rationale for international assistance to population programs in the developing world | No fulltext |
| Harvey, P. D. | 1996 | Let's not get carried away with "reproductive health" | No fulltext |
| Hashmi, S. S. | 1990 | Trends in the growth of population and labour force in Pakistan | No fulltext |
| Hata, K. | 1994 | Having its own strength | No fulltext |
| Hattingh, P. S. | 1989 | A model of adaptive population migration in South Africa | No fulltext |
| Haub, C. | 1994 | Population change in the former Soviet Republics | No fulltext |
| Heer, D. M. | 1984 | Policy and legislation affecting settlement of the Pacific region: an historical comparison of the United States and the USSR | No fulltext |
| Spasojevic, Jasmina | 2010 | Chapter 9 effects of education on adult health in Sweden: results from a natural experiment | No fulltext |
| Spiegel, Jerry M; Bonet, Mariano; Yassi, Annalee; Molina, Enrique; Concepcion, Miriam; Mast, Pedro | 2001 | Developing ecosystem health indicators in Centro Habana: a community- • based approach | No fulltext |
| Gulson, B. L.; Mahaffey, K. R.; Mizon, K. J.; Korsch, M. J.; Cameron, M. A.; Vimpani, G. | 1995 | Contribution of tissue lead to blood lead in adult female subjects based on stable lead isotope methods | No fulltext |
| Haiyan, Xing; Wei, Y. U.; Sanmei, Chen; Dengke, Zhang; Rongmei, T. A. N. | 2013 | Influence of Social Support on Health-Related Quality of Life in New-Generation Migrant Workers in Eastern China | No fulltext |

| | | | |
|---|---|---|---|
| Hsieh, Wu-Shiun; Hsieh, Chia-Jung; Jeng, Suh-Fang; Liao, Hua-Fang; Su, Yi-Ning; Lin, Shio-Jean; Chang, Pei-Jen; Chen, Pau-Chung | 2011 | Favorable Neonatal Outcomes Among Immigrants in Taiwan: Evidence of Healthy Immigrant Mother Effect | No fulltext |
| James, C. H.; Scantlebury, C. | 2007 | After the storm: when Houston becomes home | No fulltext |
| Kenny, C. | 2000 | Asylum seekers are 'new underclass' | No fulltext |
| McEwen, M. M. | 2007 | A culturally tailored diabetes social support intervention for Mexican women | No fulltext |
| Alshadood, Maytham; Harpin, Scott Butler; Puma, Jini | 2018 | Burmese and Bhutanese refugee utilization of healthcare services in Colorado | No fulltext |
| Chang, Miya | 2019 | Cross-cultural comparative study of psychological distress between older Korean immigrants in the United States and older Koreans in South Korea | No fulltext |
| DeLia, D.; DeLia, Derek | 2003 | Distributional issues in the analysis of preventable hospitalizations | No fulltext |
| Fox, P. G.; Cowell, J. M.; Montgomery, A. C.; Willgerodt, M. A. | 1998 | Southeast Asian refugee women and depression: a nursing intervention | No fulltext |
| Souza, J. B. | 1980 | Half the world in cities | No fulltext |
| Spain, D. | 1984 | Switzerland: country profile | No fulltext |
| Spain, D. | 1984 | The Kingdom of Sri Lanka: high literacy is a good sign | No fulltext |
| Spain. Ministerio de Asuntos, Sociales | 1989 | Order of 13 January 1989 regulating Reception Centres for Refugees and Asylees | No fulltext |
| Spurgeon, D. | 1992 | What do young black South Africans think about AIDS? | No fulltext |
| Srinivasan, K. | 1990 | Exodus to cities and quality of life | No fulltext |
| Staiger, B. | 1990 | First results of the fourth population census of the People's Republic of China, July 1, 1990 | No fulltext |
| Starr, P. D.; Roberts, A. E. | 1982 | Community structure and Vietnamese refugee adaptation: the significance of context | No fulltext |
| Stein, E. | 1993 | Migration patterns in Central America seen in the context of economic integration and the need for sustainable development | No fulltext |
| Steinmann, G. | 1988 | Population trends in the Federal Republic of Germany and their effects on the economy and society | No fulltext |
| Struck, D. | 1994 | Refugees, immigrants aggravate population controls | No fulltext |
| Sufian, A. J. | 1994 | Socioeconomic determinants of crowding inside home in the Eastern Province of Saudi Arabia: a comparative analysis | No fulltext |
| Sun, C. S. | 1982 | The structuring of intra-urban residential movement | No fulltext |
| United Nations. Department of International, Economic; Social, Affairs; United Nations Fund for Population Activities, Unfpa | 1980 | Guinea | No fulltext |
| United Nations. Department of International, Economic; Social, Affairs; United Nations. Fund for Population Activities, Unfpa | 1980 | Tunisia | No fulltext |
| United Nations. Department of International, Economic; Social, Affairs; United Nations. Fund for Population Activities, Unfpa | 1980 | Senegal | No fulltext |
| Stöver, Britta | 2019 | The impact of a shortened schooling time on the transition from school to studies - empirical evidence from a natural experiment | No fulltext |

| | | | |
|---|---|---|---|
| Trovato, Frank | 2017 | Reflections toward an organizing framework for the study of immigrant mortality | No fulltext |
| Tumen, Semih | n.a. | Syrian Refugees: Economic Challenges and Opportunities | No fulltext |
| Snodgrass, Jeffrey G., Upadhyay, Chakrapani; Debnath, Debashis; Lacy, Michael G. | n.a. | The Mental Health Costs of Human Displacement: A Natural Experiment Involving Indigenous Indian Conservation Refugees | No fulltext |
| Vang, Zoua M; Sigouin, Jennifer; Flenon, Astrid; Gagnon, Alain | 2017 | Are immigrants healthier than native-born Canadians? A systematic review of the healthy immigrant effect in Canada | No fulltext |
| VanLandingham, Mark; Zhang, Mengxi | 2007 | Migration and Health | No fulltext |
| Venables, Katherine | 2013 | Current topics in occupational epidemiology | No fulltext |
| | 2013 | Suicide and suicidal ideation among Bhutanese refugees-United States, 2009-2012 | No fulltext |
| Eckert, Fabian; Hejlesen, Mads; Walsh, Conor | 2019 | The return to big city experience: Evidence from Danish refugees | No fulltext |
| Vasquez, A.; Cabieses, B.; Tudesca, K. | 2018 | SPATIAL DISTRIBUTION OF SOCIOECONOMICALLY DEPRIVED IMMIGRANTS AND THEIR ACCESS TO HEALTHCARE SERVICES IN A NORTHERN CITY IN CHILE | No fulltext |
| Hinman, S. E. | 2017 | Comparing spatial distributions of infant mortality over time: Investigating the urban environment of Baltimore, Maryland in 1880 and 1920 | No fulltext |
| Becklake, M. R. | 1995 | International union against tuberculosis and lung disease (IUATLD): Initiatives in non-tuberculous lung disease | No fulltext |
| Beiser, M.; Wickrama, K. A. S. | 2004 | Trauma, time and mental health: a study of temporal reintegration and depressive disorder among Southeast Asian refugees | No fulltext |
| Giacco, D.; Laxhman, N.; Priebe, S. | 2018 | Prevalence of and risk factors for mental disorders in refugees | No fulltext |
| Ling, D. C.; Holroyd, E. A.; Wong, W. C. W.; Gray, A. | 2004 | Handling emerging health needs among a migrant population -- factors associated with suicide attempts and suicide ideation among female street sex workers in Hong Kong | No fulltext |
| Kim, I. | 2018 | Behavioral Health Symptoms Among Refugees From Burma: Examination of Sociodemographic and Migration-Related Factors | No fulltext |
| Okamoto, E. | 2008 | Mortality in East Asian countries in the pre-war period: a quasi-experimental study on healthy immigrant effects | No fulltext |
| Takeda, J. | 2000 | Psychological and economic adaptation of Iraqi adult male refugees: Implications for social work practice | No fulltext |
| Santas, Gulcan; Eryurt, Mehmet Ali | 2020 | Distribution of child health indicators according to internal migration and various social variables in Turkey | No fulltext |
| Lu, S. F.; Chen, S. X.; Wang, P. G. | 2019 | Language barriers and health status of elderly migrants: Micro-evidence from China | No fulltext |
| Qin, Lijian; Chen, Chien-Ping; Wang, Wei; Chen, Hong | 2021 | How migrants get integrated in urban China - The impact of health insurance | No health outcomes |
| Puma, Jini E.; Brewer, Sarah E.; Stein, Paul | 2020 | Pathways to Refugee Integration: Predictions from Longitudinal Data in Colorado | No health outcomes |
| Hedegaard, Troels Fage; Bekhuis, Hidde | 2018 | A migration effect? Comparing the acculturation of Russian migrant populations in Western Europe to Russians in three former soviet countries on attitudes towards government responsibility | No health outcomes |
| Gibson, Mhairi A; Gurmu, Eshetu | 2012 | Rural to urban migration is an unforeseen impact of development intervention in Ethiopia | No health outcomes |

| | | | |
|---|---|---|---|
| Akgüc, Mehtap; Liu, Xingfei; Tani, Massimiliano | 2014 | Expropriation with hukou change: Evidence from a quasi-natural experiment | No health outcomes |
| Ghimire, Keshar M | 2017 | Impact of Children's Health Insurance Bene fit on Labor Supply: Evidence from Newly Arrived Immigrants in the United States | No health outcomes |
| Cockx, L.; Colen, L.; De Weerdt, J. | 2018 | From corn to popcorn? Urbanization and dietary change: Evidence from rural-urban migrants in Tanzania | No health outcomes |
| Lucero, Jessica L.; Santiago, Anna Maria; Galster, George C. | 2018 | How Neighborhood Effects Vary: Childbearing and Fathering among Latino and African American Adolescents | No health outcomes |
| Schultz-Nielsen, M. L.; Tekin, E.; Greve, J. | 2016 | Labor market effects of intrauterine exposure to nutritional deficiency: Evidence from administrative data on Muslim immigrants in Denmark | No health outcomes |
| Mata, F.; Pendakur, R. | 1999 | Immigration, labor force integration and the pursuit of self-employment | No health outcomes |
| Matin, K. A. | 1986 | Residence background and fertility in Chittagong, Bangladesh | No health outcomes |
| McCutcheon, L. | 1984 | Measuring migrant change: a look at housing in Bogota, Seoul, and Surabaya | No health outcomes |
| McDevitt, T. M.; Hawley, A. H.; Udry, J. R.; Gadalla, S.; Leoprapai, B.; Cardona, R. | 1986 | Migration plans of the rural populations of the Third World countries: a probit analysis of micro-level data from Asia, Africa, and Latin America | No health outcomes |
| Tien, H. Y. | 1981 | Demography in China: from zero to now | No health outcomes |
| Shams ur, rahman; Clarke, H. R. | 1991 | A nutrition model for developing nations with special reference to Bangladesh | No health outcomes |
| Vasileva, D. | 1992 | Bulgarian Turkish emigration and return | No health outcomes |
| Watkins, J. F.; Leinbach, T. R.; Falconer, K. F. | 1993 | Women, family, and work in Indonesian transmigration | No health outcomes |
| Yadava, K. N.; Singh, R. B. | 1991 | A probability model for the distribution of the number of migrants at the household level | No health outcomes |
| Kreibaum, Merle | 2016 | Their suffering, our burden? How Congolese refugees affect the Ugandan population | No health outcomes |
| Lazareva, Olga | 2015 | Russian migrants to Russia: assimilation and local labor market effects | No health outcomes |
| Li, Weibing; Zhang, Kaixia | 2019 | Does Air Pollution Crowd Out Foreign Direct Investment Inflows? Evidence from a Quasi-natural Experiment in China | No health outcomes |
| Makovec, Mattia; Purnamasari, Ririn; Sandi, Matteo; Savitri, Astrid | 2016 | Intended vs. unintended consequences of migration restriction policies: evidence from a natural experiment in Indonesia | No health outcomes |
| Makovec, Mattia; Purnamasari, Ririn S; Sandi, Matteo; Savitri, Astrid R | 2018 | Intended versus unintended consequences of migration restriction policies: evidence from a natural experiment in Indonesia | No health outcomes |
| Martén, Linna; Hainmueller, Jens; Hangartner, Dominik | 2019 | Ethnic networks can foster the economic integration of refugees | No health outcomes |
| Mastrorocco, Nicola; Minale, Luigi | 2016 | Information and crime perceptions: evidence from a natural experiment | No health outcomes |
| Maystadt, Jean-Francois | 2012 | Poverty Reduction in a Refugee-Hosting Economy. A Natural Experiment | No health outcomes |
| McCain, Barbara; Simmons, Kevin M; Willner, Jonathon | 2012 | Tornadoes As A Natural Experiment In Rational Choice | No health outcomes |

| | | | |
|---|---|---|---|
| McIntosh, Molly Fifer | 2008 | Measuring the labor market impacts of Hurricane Katrina migration: Evidence from Houston, Texas | No health outcomes |
| McKenzie, David; Gibson, John; Stillman, Steven | 2007 | Moving to opportunity, leaving behind what? Evaluating the initial effects of a migration policy on incomes and poverty in source areas | No health outcomes |
| McKenzie, David; Stillman, Steven; Gibson, John | 2010 | How important is selection? Experimental vs. non-experimental measures of the income gains from migration | No health outcomes |
| Mesoudi, Alex; Magid, Kesson; Hussain, Delwar | 2016 | How do people become WEIRD? Migration reveals the cultural transmission mechanisms underlying variation in psychological processes | No health outcomes |
| Montalvo, Jose G | 2011 | Voting after the bombings: A natural experiment on the effect of terrorist attacks on democratic elections | No health outcomes |
| Orbeta Jr, Aniceto | 2008 | Economic impact of international migration and remittances on Philippine households: What we thought we knew, what we need to know | No health outcomes |
| Parsons, Christopher; Vézina, Pierre-• Louis | 2018 | Migrant networks and trade: The Vietnamese boat people as a natural experiment | No health outcomes |
| Pedersen, Peder J; Pytlikova, Mariola | 2008 | EU Enlargement: Migration Flows from Central and Eastern Europe Into the Nordic Countries: Exploiting a Natural Experiment | No health outcomes |
| Phan, Tuan Q; Airoldi, Edoardo M | 2015 | A natural experiment of social network formation and dynamics | No health outcomes |
| Ramachandran, Rajesh | 2012 | Language use in education and primary schooling attainment: evidence from a natural experiment in Ethiopia | No health outcomes |
| Roth, Christopher; Sumarto, Sudarno | 2015 | Does education increase interethnic and interreligious tolerance? Evidence from a natural experiment | No health outcomes |
| Ruiz, Isabel; Vargas-Silva, Carlos | 2015 | The labor market impacts of forced migration | No health outcomes |
| Schlosser, Analia | 2005 | Public preschool and the labor supply of Arab mothers: Evidence from a natural experiment | No health outcomes |
| Schrank, Andrew | 2006 | Labor standards and human resources: A natural experiment in an unlikely laboratory | No health outcomes |
| Spaan, Ernst; van Naerssen, Ton | 2018 | Migration decision-making and migration industry in the Indonesia - Malaysia corridor | No health outcomes |
| Tumen, Semih | 2016 | The economic impact of Syrian refugees on host countries: Quasi-experimental evidence from Turkey | No health outcomes |
| Valentova, Marie; Callens, Marie-Sophie | 2018 | Did the escalation of the financial crisis of 2008 affect the perception of immigration-related threats? A natural experiment | No health outcomes |
| Venta, Amanda | 2020 | Attachment Facilitates Acculturative Learning and Adversity Moderates: Validating the Theory of Epistemic Trust in a Natural Experiment | No health outcomes |
| Jordan, Bill; Brown, Philip | 2006 | The Sangatte work-visa holders: a natural experiment in immigration policy | No health outcomes |
| Hui, Kai-Lung; Png, Ivan PL | 2012 | Digital Divide? Evidence from a Federal Natural Experiment | No health outcomes |
| Kirk, David S | 2008 | Lessons from Hurricane Katrina: A Natural Experiment of the Effect of Residential Change on Recidivism | No health outcomes |
| Gibson, John; Mackenzie, D; Rohorua, Halahingano; Stillman, Steven | 2006 | Natural Experiment Evidence on Whether Selection Bias Overstates the Gains from Migration | No health outcomes |

| | | | |
|---|---|---|---|
| Elsner, Benjamin | 2011 | Does Emigration Benefit the Stayers? The EU Enlargement as a Natural Experiment: Evidence from Lithuania | No health outcomes |
| Kirk, David S | 2008 | The Effect of Hurricane Katrina on Prisoner Reentry in Louisiana: A Natural Experiment | No health outcomes |
| Patulny, Roger; Siminski, Peter; Mendolia, Silvia | 2015 | The front line of social capital creation - A natural experiment in symbolic interaction | No health outcomes |
| Mikula K; Pytlikova M. | 2020 | Air Pollution and Migration – exploiting a natural experiment from the Czech Republic | No health outcomes |
| Choudhury, Prithwiraj | 2016 | Return migration and geography of innovation in MNEs: a natural experiment of knowledge production by local workers reporting to return migrants | No health outcomes |
| Ceritoglu, Evren; Yunculer, H Burcu Gurcihan; Torun, Huzeyfe; Tumen, Semih | 2017 | The impact of Syrian refugees on natives labor market outcomes in Turkey: evidence from a quasi-experimental design | No health outcomes |
| Amuedo-Dorantes, Catalina; Pozo, Susan | 2006 | Migration, remittances, and male and female employment patterns | No health outcomes |
| Carlino, G. A.; Mills, E. S. | 1987 | The determinants of county growth | No health outcomes |
| Balkan, Binnur; Tumen, Semih | 2016 | Immigration and prices: quasi-experimental evidence from Syrian refugees in Turkey | No health outcomes |
| van de Crommenacker, Janske; Komdeur, Jan; Burke, Terry; Richardson, David S | 2011 | Spatio-• temporal variation in territory quality and oxidative status: a natural experiment in the Seychelles warbler (Acrocephalus sechellensis) | No health outcomes |
| Sell, R. R. | 1983 | Transferred jobs: a neglected aspect of migration and occupational change | No health outcomes |
| Sarris, A. H.; Zografakis, S. | 1999 | A computable general equilibrium assessment of the impact of illegal immigration on the Greek economy | No health outcomes |
| Beguin, H. | 1982 | The effect of urban spatial structure on residential mobility | No health outcomes |
| Marbach, Moritz; Hainmueller, Jens; Hangartner, Dominik | 2018 | The long-term impact of employment bans on the economic integration of refugees | No health outcomes |
| Antman, Francisca M | 2011 | International migration and gender discrimination among children left behind | No health outcomes |
| Gezie, L. D.; Worku, A.; Kebede, Y.; Gebeyehu, A. | 2019 | Sexual violence at each stage of human trafficking cycle and associated factors: a retrospective cohort study on Ethiopian female returnees via three major trafficking corridors | No health outcomes |
| Gichunge, Catherine; Somerset, Shawn; Harris, Neil | 2016 | Using a Household Food Inventory to Assess the Availability of Traditional Vegetables among Resettled African Refugees | No health outcomes |
| Borra, Cristina; Pons-Pons, Jerònia; Vilar-Rodríguez, Margarita | 2020 | Austerity, healthcare provision, and health outcomes in Spain | No migrant population |
| Friedman, Abigail S.; Venkataramani, Atheendar S. | 2021 | Chilling Effects: US Immigration Enforcement And Health Care Seeking Among Hispanic Adults | No migrant population |
| Bertoli, Paola; Grembi, Veronica; Nguyen, The Linh Bao | 2021 | Birth outcomes in hard times among minority ethnic groups | No migrant population |
| Hossain, Altaf; Alam, Md. Jahangir; Haque, Md. Rezaul | 2021 | Effects of riverbank erosion on mental health of the affected people in Bangladesh | No migrant population |
| Fonseca-Rodriguez, Osvaldo; Gustafsson, Per E.; San Sebastian, Miguel; Connolly, Anne-Marie Fors | 2021 | Spatial clustering and contextual factors associated with hospitalisation and deaths due to COVID-19 in Sweden: a geospatial nationwide ecological study | No migrant population |

| | | | |
|---|---|---|---|
| Baez, Javier E | 2011 | Civil wars beyond their borders: The human capital and health consequences of hosting refugees | No migrant population |
| Quis, Johanna; Reif, Simon | 2017 | Health effects of instruction intensity: Evidence from a natural experiment in German high-schools | No migrant population |
| Nakhaie, R.; Arnold, R. | 2010 | A four year (1996-2000) analysis of social capital and health status of Canadians: The difference that love makes | No migrant population |
| Yemelyanau, Maksim; Amialchuk, Aliaksandr; Ali, Mir M | 2012 | Evidence from the chernobyl nuclear accident: the effect on health, education, and labor market outcomes in belarus | No migrant population |
| Gibson, John; McKenzie, David; Stillman, Steven | 2011 | The impacts of international migration on remaining household members: omnibus results from a migration lottery program | No migrant population |
| Loevinsohn, Michael | 2015 | The 2001-03 famine and the dynamics of HIV in Malawi: A natural experiment | No migrant population |
| Koksal, Aycan; Wohlgenant, Michael | 2013 | Interdependence of tobacco and alcohol consumption: a natural experiment approach | No migrant population |
| Krueger, Alan B; Kuziemko, Ilyana | 2013 | The demand for health insurance among uninsured Americans: Results of a survey experiment and implications for policy | No migrant population |
| Mattingly, Daniel C | 2017 | Colonial legacies and state institutions in China: Evidence from a natural experiment | No migrant population |
| McKenzie, David | 2015 | Learning about migration through experiments | No migrant population |
| Meng, Xin; Qian, Nancy | 2009 | The long term consequences of famine on survivors: evidence from a unique natural experiment using China's great famine | No migrant population |
| Moen, Phyllis; Kelly, Erin; Chermack, Kelly | 2009 | Learning from a natural experiment: Studying a corporate work-time policy initiative | No migrant population |
| Mölenberg, Famke JM; Noordzij, J Mark; Burdorf, Alex; van Lenthe, Frank J | 2019 | New physical activity spaces in deprived neighborhoods: Does it change outdoor play and sedentary behavior? A natural experiment | No migrant population |
| Ong, Qiyan; Theseira, Walter | 2016 | Does choosing jobs based on income risk lead to higher job satisfaction in the long run? Evidence from the natural experiment of German reunification | No migrant population |
| Pantano, Juan | 2007 | Unwanted fertility, contraceptive technology and crime: Exploiting a natural experiment in access to the pill | No migrant population |
| Pesonen, Anu-• Katriina; Räikkönen, Katri; Heinonen, Kati; Kajantie, Eero; Forsén, Tom; Eriksson, Johan G | 2008 | Reproductive traits following a parent-child separation trauma during childhood: a natural experiment during World War II | No migrant population |
| Ponomareva, Katia | 2016 | Maternity Leave and Long-Run Health Outcomes of Children: A Natural Experiment in Soviet Russia | No migrant population |
| Monstad, Karin; Propper, Carol; Salvanes, Kjell G | 2008 | Education and fertility: Evidence from a natural experiment | No migrant population |
| Simonet, Fabienne; Wilkins, Russell; Labranche, Elena; Smylie, Janet; Heaman, Maureen; Martens, Patricia; Fraser, William D; Minich, Katherine; Wu, Yuquan; Carry, Catherine | 2009 | Primary birthing attendants and birth outcomes in remote Inuit communities - a natural -œexperiment-• in Nunavik, Canada | No migrant population |
| Snodgrass, Jeffrey G; Lacy, Michael G; Upadhyay, Chakrapani | 2017 | Developing culturally sensitive affect scales for global mental health research and practice: Emotional balance, not named syndromes, in Indian Adivasi subjective well-being | No migrant population |
| Palència, Laia; Malmusi, Davide; De Moortel, Deborah; Artazcoz, Lucía; Backhans, Mona; Vanroelen, Christophe; Borrell, Carme | 2014 | The influence of gender equality policies on gender inequalities in health in Europe | No migrant population |
| Stevenson, Betsey; Wolfers, Justin | 2013 | Subjective well-being and income: Is there any evidence of satiation? | No migrant population |

| | | | |
|---|---|---|---|
| Terraube, Julien; Fernández-Llamazares, Ã• lvaro; Cabeza, Mar | 2017 | The role of protected areas in supporting human health: a call to broaden the assessment of conservation outcomes | No migrant population |
| Van Gelder, Jean-• Louis | 2013 | Then I'll Huff, and I'll Puff, and I'll...: A natural experiment on property titling, housing improvement and the psychology of tenure security | No migrant population |
| Vargas, Andres J | 2011 | The effect of social security contributions on coverage and wages: a gender perspective using a natural experiment from Colombia | No migrant population |
| Wang, Jia | 2016 | Village Migration, Kinship Networks, and Subjective Well-Being among Adults Staying Behind in Rural China | No migrant population |
| Xu, Hongwei; Li, Lydia; Zhang, Zhenmei; Liu, Jinyu | 2016 | Is natural experiment a cure? Re-examining the long-term health effects of China's 1959-1961 famine | No migrant population |
| Hofmann, Sarah; Mühlenweg, Andrea | 2018 | Learning intensity effects in students' mental and physical health - Evidence from a large scale natural experiment in Germany | No migrant population |
| Huang, Cheng; Phillips, Michael R; Zhang, Yali; Zhang, Jingxuan; Shi, Qichang; Song, Zhiqiang; Ding, Zhijie; Pang, Shutao; Martorell, Reynaldo | 2013 | Malnutrition in early life and adult mental health: evidence from a natural experiment | No migrant population |
| Hanna, Rema; Oliva, Paulina | 2015 | The effect of pollution on labor supply: Evidence from a natural experiment in Mexico City | No migrant population |
| Dow, William H; Schmeer, Kammi K | 2003 | Health insurance and child mortality in Costa Rica | No migrant population |
| Carliner, G.; Robinson, C.; Tomes, N. | 1984 | Lifetime models of female labor supply, wage rates, and fertility | No migrant population |
| Bluedorn, John C; Cascio, Elizabeth | 2005 | Education and intergenerational mobility: Evidence from a natural experiment in Puerto Rico | No migrant population |
| Bae, H. | 1987 | Changing family structure and aging issues | No migrant population |
| Azam, Mehtabul | 2011 | The impact of Indian job guarantee scheme on labor market outcomes: Evidence from a natural experiment | No migrant population |
| Antwi, James; Phillips, David | 2011 | Wages and Health Worker Retention: Evidence from Public Sector Wage Reforms in Ghana | No migrant population |
| Anderson, Benjamin C | 2010 | Consumption Smoothing Responses to Natural Disasters: Evidence from a Quasi-Natural Experiment in Nicaragua | No migrant population |
| Algan, Yann; Do, Quoc-Anh; Dalvit, Nicolò; Le Chapelain, Alexis; Zenou, Yves | 2015 | How social networks shape our beliefs: A natural experiment among future French politicians | No migrant population |
| Aksoy, Ozan; Billari, Francesco C | 2018 | Political Islam, marriage, and fertility: evidence from a natural experiment | No migrant population |
| Échevin, Damien; Fortin, Bernard | 2014 | Physician payment mechanisms, hospital length of stay and risk of readmission: evidence from a natural experiment | No migrant population |
| Herttua, Kimmo | 2010 | The Effects of the 2004 Reduction in the Price of Alcohol on Alcohol-Related Harm in Finland - a Natural Experiment Based on Register Data | No migrant population |
| Ólafsdóttir, Thorhildur; Hrafnkelsson, Birgir; Thorgeirsson, Gudmundur; Á• sgeirsdóttir, Tinna Laufey | 2016 | The tax-free year in Iceland: A natural experiment to explore the impact of a short-term increase in labor supply on the risk of heart attacks | No migrant population |
| Cunningham, Scott; Kendall, Todd D | 2011 | Men in transit and prostitution: Using political conventions as a natural experiment | No migrant population |
| Chaudhury, Nazmul; Hammer, Jeffrey; Kremer, Michael; Muralidharan, Karthik; Rogers, F Halsey | 2006 | Missing in action: teacher and health worker absence in developing countries | No migrant population |

| | | | |
|---|---|---|---|
| Carson, S. A. | 2015 | Biological Conditions and Economic Development | No migrant population |
| Bönisch, Peter; Hyll, Walter | 2015 | Television Role Models and Fertility - Evidence from a Natural Experiment | No migrant population |
| Berger, Daniel | 2009 | Taxes, institutions and local governance: evidence from a natural experiment in colonial Nigeria | No migrant population |
| Baez, Javier E; Santos, Indhira V | 2007 | Children's vulnerability to weather shocks: A natural disaster as a natural experiment | No migrant population |
| Astell-Burt, Thomas; Feng, Xiaoqi; Kolt, Gregory S; Jalaludin, Bin | 2015 | Does rising crime lead to increasing distress? Longitudinal analysis of a natural experiment with dynamic objective neighbourhood measures | No migrant population |
| Aoki, Yu | 2011 | An Outcome of Free Labour Supply: Effect of Volunteer Work on Mortality | No migrant population |
| Aoki, Yu | 2011 | Does volunteering reduce mortality? Evidence from a natural experiment in japan | No migrant population |
| White, James; Greene, Giles; Farewell, Daniel; Dunstan, Frank; Rodgers, Sarah; Lyons, Ronan A; Humphreys, Ioan; John, Ann; Webster, Chris; Phillips, Ceri J | 2017 | Improving mental health through the regeneration of deprived neighborhoods: a natural experiment | No migrant population |
| White, James; Greene, Giles; Dunstan, Frank; Rodgers, Sarah; Lyons, Ronan A; Humphreys, Ioan; John, Ann; Webster, Chris; Palmer, Stephen; Elliott, Eva | 2014 | The Communities First (ComFi) study: protocol for a prospective controlled quasi-experimental study to evaluate the impact of area-wide regeneration on mental health and social cohesion in deprived communities | No migrant population |
| Vandoros, Sotiris; Avendano, Mauricio; Kawachi, Ichiro | 2019 | The EU referendum and mental health in the short term: a natural experiment using antidepressant prescriptions in England | No migrant population |
| Torche, Florencia | 2011 | The effect of maternal stress on birth outcomes: exploiting a natural experiment | No migrant population |
| Thern, Emelie; de Munter, Jeroen; Hemmingsson, Tomas; Davey Smith, George; Ramstedt, Mats; Tynelius, Per; Rasmussen, Finn | 2017 | Effects of increased alcohol availability during adolescence on the risk of all-• cause and cause-• specific disability pension: a natural experiment | No migrant population |
| Peres, MA; Peres, KG; Barbato, PR; Höfelmann, DA | 2016 | Access to fluoridated water and adult dental caries: a natural experiment | No migrant population |
| Loopstra, Rachel; Reeves, Aaron; McKee, Martin; Stuckler, David | 2016 | Food insecurity and social protection in Europe: quasi-natural experiment of Europe's great recessions 2004-2012 | No migrant population |
| Hsin, O.; La Greca, A. M.; Valenzuela, J.; Moine, C. T.; Delamater, A. | 2010 | Adherence and Glycemic Control among Hispanic Youth with Type 1 Diabetes: Role of Family Involvement and Acculturation | No migrant population |
| Ball, K.; Mishra, G.; Crawford, D. | 2002 | Which aspects of socioeconomic status are related to obesity among men and women? | No migrant population |
| He, Ping; Liu, Li; Salas, JM Ian; Guo, Chao; Cheng, Yunfei; Chen, Gong; Zheng, Xiaoying | 2018 | Prenatal malnutrition and adult cognitive impairment: a natural experiment from the 1959-1961 Chinese famine | No migrant population |
| Droomers, Mariël; Jongeneel-Grimen, Birthe; Kramer, Daniëlle; de Vries, Sjerp; Kremers, Stef; Bruggink, Jan-Willem; van Oers, Hans; Kunst, Anton E; Stronks, Karien | 2016 | The impact of intervening in green space in Dutch deprived neighbourhoods on physical activity and general health: results from the quasi-experimental URBAN40 study | No migrant population |
| Braam, Arjan W.; van Ommeren, Omar W. H. R.; van Buuren, Melissa L.; Laan, Wijnand; Smeets, Hugo M.; Engelhard, Iris M. | 2016 | Local Geographical Distribution of Acute Involuntary Psychiatric Admissions in Subdistricts In and Around Utrecht, the Netherlands | No migrant population |
| Case, Anne; Paxson, Christina | 2008 | Height, health, and cognitive function at older ages | No migrant population |
| Chen, H. J.; Wang, Y. F. | 2014 | The changing food outlet distributions and local contextual factors in the United States | No migrant population |
| Dongyang, Yang; Chengdong, Xu; Jinfeng, Wang; Yong, Zhao; Yang, Dongyang; Xu, Chengdong; Wang, Jinfeng; Zhao, Yong | 2017 | Spatiotemporal epidemic characteristics and risk factor analysis of malaria in Yunnan Province, China | No migrant population |

| | | | | |
|---|---|---|---|---|
| Crosse, E. A.; Alder, R. J.; Ostbye, T.; Campbell, M. K.; Crosse, E. A.; Alder, R. J.; Ostbye, T.; Campbell, M. K. | 1997 | Small area variation in low birthweight: looking beyond socioeconomic predictors | No migrant population | |
| Czaderny, K. | 2019 | Lung cancer mortality in historical context: How stable are spatial patterns of smoking over time? | No migrant population | |
| Egan, Matt; Katikireddi, Srinivasa Vittal; Kearns, Ade; Tannahill, Carol; Kalacs, Martins; Bond, Lyndal | 2013 | Health effects of neighborhood demolition and housing improvement: a prospective controlled study of 2 natural experiments in urban renewal | No migrant population | |
| Fernandez, M. D.; Gaspe, M. S.; Gurtler, R. E. | 2019 | Inequalities in the social determinants of health and Chagas disease transmission risk in indigenous and creole households in the Argentine Chaco | No migrant population | |
| Glazier, R. H.; Creatore, M. I.; Cortinois, A. A.; Agha, M. M.; Moineddin, R.; Glazier, Richard H.; Creatore, Maria I.; Cortinois, Andrea A.; Agha, Mohammad M.; Moineddin, Rahim | 2004 | Neighbourhood recent immigration and hospitalization in Toronto, Canada...Proceedings of the National Symposium on Immigrant Health in Canada | No migrant population | |
| Han, Bing; Chen, Yi; Cheng, Jing; Li, Qin; Zhu, Chunfang; Chen, Yingchao; Xia, Fangzhen; Wang, Ningjian; Lu, Yingli | 2018 | Comparison of the Prevalence of Metabolic Disease Between Two Types of Urbanization in China | No migrant population | |
| De Neve, Jan-Walter; Fink, Günther; Subramanian, SV; Moyo, Sikhulile; Bor, Jacob | 2015 | Length of secondary schooling and risk of HIV infection in Botswana: evidence from a natural experiment | No migrant population | |
| Glenn, L. L.; Beck, R. W.; Burkett, G. L. | 1998 | Effect of a transient, geographically localised economic recovery on community health and income studied with longitudinal household cohort interview method | No migrant population | |
| Ajefu, Joseph B.; Ogebe, Joseph O. | 2021 | The effects of international remittances on expenditure patterns of the left-behind households in Sub-Saharan Africa | No migrant population | |
| Jirmanus LZ,Ranker L,Touw S,Mahmood R,Kimball SL,Hanchate A,Lasser KE | 2022 | Impact of United States 2017 Immigration Policy changes on missed appointments at two Massachusetts Safety-Net Hospitals | No migrant population | |
| Haque, Rabiul; Parr, Nick; Muhidin, Salut | 2020 | Climate-related displacement, impoverishment and healthcare accessibility in mainland Bangladesh | No natural experiment | |
| Schmalbach, Bjarne; Schmalbach, Ileana; Kasinger, Christoph; Petrowski, Katja; Braehler, Elmar; Zenger, Markus; Stoebel-Richter, Yve; Richter, Ernst Peter; Berth, Hendrik | 2021 | Psychological and Socio-Economical Determinants of Health: The Case of Inner German Migration | No natural experiment | |
| Powell, Tara M.; Li, Shang-Ju; Hsiao, Yuan; Thompson, Michelle; Farraj, Aseel; Abdoh, Mariam; Farraj, Rami | 2021 | An integrated physical and mental health awareness education intervention to reduce non-communicable diseases among Syrian refugees and Jordanians in host communities: A natural experiment study | No natural experiment | claims to be natural experiment, but no random allocation |
| Yang, Min; Hagenauer, Julian; Dijst, Martin; Helbich, Marco | 2021 | Assessing the perceived changes in neighborhood physical and social environments and how they are associated with Chinese internal migrants' mental health | No natural experiment | |
| Solberg, Øivind; Vaez, Marjan; Johnson-Singh, Charisse M.; Saboonchi, Fredrik | 2020 | Asylum-seekers' psychosocial situation: A diathesis for post-migratory stress and mental health disorders? | No natural experiment | |
| Khan, Sanjida; Haque, Shamsul | 2021 | Trauma, mental health, and everyday functioning among Rohingya refugee people living in short- and long-term resettlements | No natural experiment | |
| Holmager, Therese Lucia Friis; Lophaven, Søren Nymand; Mortensen, Laust Hvas; Lynge, Elsebeth | 2021 | Does Lolland-Falster make people sick, or do sick people move to Lolland-Falster? An example of selective migration and mortality in Denmark, 1968-2017 | No natural experiment | |
| Fares, Hani; Puig-Junoy, Jaume | 2021 | Inequity and benefit incidence analysis in healthcare use among Syrian refugees in Egypt | No natural experiment | |
| Dangmann, Cecilie; Solberg, Oivind; Andersen, Per Normann | 2021 | Health-related quality of life in refugee youth and the mediating role of mental distress and post-migration stressors | No natural experiment | |
| Berry, John W.; Hou, Feng | 2021 | Immigrant acculturation and wellbeing across generations and settlement contexts in Canada | No natural experiment | |

| | | | | |
|---|---|---|---|---|
| Hawkes, Clare; Norris, Kimberley; Joyce, Janine; Paton, Douglas | 2021 | Individuals of refugee background resettled in regional and rural Australia: A systematic review of mental health research | No natural experiment | |
| Peterson, Cynthia; Poudel-Tandukar, Kalpana; Sanger, Kirk; Jacelon, Cynthia S. | 2020 | Improving Mental Health in Refugee Populations: A Review of Intervention Studies Conducted in the United States | No natural experiment | |
| Hakimi, Razia; Kheirkhah, Masoomeh; Abolghasemi, Jamileh; Hakimi, Masumah | 2021 | Sex education and Afghan migrant adolescent women | No natural experiment | |
| Lyles, Emily; Chua, Stephen; Barham, Yasmeen; Pfieffer-Mundt, Kayla; Spiegel, Paul; Burton, Ann; Doocy, Shannon | 2021 | Improving diabetes control for Syrian refugees in Jordan: a longitudinal cohort study comparing the effects of cash transfers and health education interventions | No natural experiment | claims to be natural experiment, but is an intervention study |
| Wanigaratne, Susitha; Wiedmeyer, Mei-ling; Brown, Hilary K.; Guttmann, Astrid; Urquia, Marcelo L. | 2020 | Induced abortion according to immigrants' birthplace: a population-based cohort study | No natural experiment | |
| Russo, Rienna; Kwon, Simona; Tsui, Jennifer; Yi, Stella S. | 2020 | Immigrants Prioritize Language Over Geography When Accessing Health Care | No natural experiment | |
| Ghattas, Hala; Choufani, Jowel; Jamaluddine, Zeina; Masterson, Amelia Reese; Sahyoun, Nadine R. | 2020 | Linking women-led community kitchens to school food programmes: lessons learned from the Healthy Kitchens, Healthy Children intervention in Palestinian refugees in Lebanon | No natural experiment | claims to be natural experiment, but is an intervention study |
| Dane, Sharon; Haslam, Catherine; Jetten, Jolanda; Liu, Shuang; Gallois, Cindy; Le Tran, Tran Nghi | 2020 | The benefits of ethnic activity group participation on older immigrant well-being and host country adjustment | No natural experiment | claims to be natural experiment, but is an intervention study |
| Endrst, J. | 1979 | Botswana's self-help community | No natural experiment | |
| Basu, Sarah | 2012 | Mental health concerns for Indian women | No natural experiment | |
| Al Obaidi, A. K. | 2010 | Iraqi psychiatrist in exile helping distressed iraqi refugee children in egypt in non-clinical settings | No natural experiment | |
| Lindencrona, F.; Ekblad, S.; Hauff, E.; Lindencrona, Fredrik; Ekblad, Solvig; Hauff, Edvard | 2008 | Mental health of recently resettled refugees from the Middle East in Sweden: the impact of pre-resettlement trauma, resettlement stress and capacity to handle stress | No natural experiment | |
| Eisenberg, Katherine W.; van Wijngaarden, Edwin; Fisher, Susan G.; Korfmacher, Katrina S.; Campbell, James R.; Fernandez, I. Diana; Cochran, Jennifer; Geltman, Paul L. | 2011 | Blood lead levels of refugee children resettled in Massachusetts, 2000 to 2007 | No natural experiment | |
| Snodgrass, Jeffrey G; Upadhyay, Chakrapani; Debnath, Debashis; Lacy, Michael G | 2016 | The mental health costs of human displacement: A natural experiment involving indigenous Indian conservation refugees | No natural experiment | claims to be natural experiment, but no random allocation |
| Mizen, Amy; Song, Jiao; Fry, Richard; Akbari, Ashley; Berridge, Damon; Parker, Sarah C; Johnson, Rhodri; Lovell, Rebecca; Lyons, Ronan A; Nieuwenhuijsen, Mark | 2019 | Longitudinal access and exposure to green-blue spaces and individual-level mental health and well-being: protocol for a longitudinal, population-wide record-linked natural experiment | No natural experiment | Protocol |
| Diez, Elia; Lopez, Maria J.; Perez, Gloria; Garcia-Subirats, Irene; Nebot, Laia; Carreras, Ramon; Villalbi, Joan R. | 2020 | Impact of a community contraceptive counselling intervention on adolescent fertility rates: a quasi-experimental study | No natural experiment | claims to be natural experiment, but is an intervention study |
| Craig, Peter; Cooper, C; Gunnell, D; Haw, S; Lawson, K; Macintyre, S; Ogilvie, D; Petticrew, M; Reeves, B; Sutton, M | 2010 | Using natural experiments to evaluate population health interventions | No natural experiment | |
| Hjern, A.; Angel, B.; Jeppson, O. | 1998 | Political violence, family stress and mental health of refugee children in exile | No natural experiment | |
| Khoshmohabat, Hadi; Motamedi, Mohammad Hosein Kalantar; Saghafinia, Masoud; Shams, Amin | 2014 | Immigration for health care in Iran: burden or blessing? | No natural experiment | |
| Blight, K. J.; Ekblad, S.; Persson, J.; Ekberg, J. | 2006 | Mental health, employment and gender. Cross-sectional evidence in a sample of refugees from Bosnia-Herzegovina living in two Swedish regions | No natural experiment | |

| | | | |
|---|---|---|---|
| Beiser, M. N. M.; Hou, F. | 2006 | Ethnic identity, resettlement stress and depressive affect among Southeast Asian refugees in Canada | No natural experiment |
| Correa-Velez, Ignacio; Gifford, Sandra M.; McMichael, Celia | 2015 | The persistence of predictors of wellbeing among refugee youth eight years after resettlement in Melbourne, Australia | No natural experiment |
| Franzini, L.; Fernandez-Esquer, M. E. | 2004 | Socioeconomic, cultural, and personal influences on health outcomes in low income Mexican-origin individuals in Texas | No natural experiment |
| Finnvold, Jon Erik; Ugreninov, Elisabeth | 2018 | Refugees' admission to mental health institutions in Norway: Is there an ethnic density effect? | No natural experiment |
| Clemens, Michael A; Özden, Çağlar; Rapoport, Hillel | 2014 | Migration and development research is moving far beyond remittances | No natural experiment |
| Gottlieb, N.; Weinstein, T.; Mink, J.; Ghebrezghiabher, H. M.; Sultan, Z.; Reichlin, R. | 2017 | Applying a community-based participatory research approach to improve access to healthcare for Eritrean asylum-seekers in Israel: a pilot study | No natural experiment |
| Garnier, D.; Simondon, K. B.; Hoarau, T.; Benefice, E. | 2003 | Impact of the health and living conditions of migrant and non-migrant Senegalese adolescent girls on their nutritional status and growth | No natural experiment |
| Fang, Z.; Sakellariou, C. | 2016 | Social Insurance, Income and Subjective Well-Being of Rural Migrants in China-An Application of Unconditional Quantile Regression | No natural experiment |
| Fishman, S. H.; Morgan, S. P.; Hummer, R. A. | 2018 | Smoking and Variation in the Hispanic Paradox: A Comparison of Low Birthweight Across 33 US States | No natural experiment |
| Ziersch, Anna; Due, Clemence; Walsh, Moira | 2020 | Discrimination: a health hazard for people from refugee and asylum-seeking backgrounds resettled in Australia | No natural experiment |
| Zöller, Bengt; Li, Xinjun; Sundquist, Jan; Sundquist, Kristina | 2013 | Neighbourhood deprivation and hospitalization for atrial fibrillation in Sweden | No natural experiment |
| Whitsett, David; Sherman, Martin F. | 2017 | Do resettlement variables predict psychiatric treatment outcomes in a sample of asylum-seeking survivors of torture? | No natural experiment |
| Schick, M.; Zumwald, A.; Knopfli, B.; Nickerson, A.; Bryant, R. A.; Schnyder, U.; Muller, J.; Morina, N. | 2016 | Challenging future, challenging past: the relationship of social integration and psychological impairment in traumatized refugees | No natural experiment |
| Saposnik, G.; Redelmeier, D. A.; Lu, H.; Lonn, E.; Fuller-Thomson, E.; Ray, J. G. | 2010 | Risk of premature stroke in recent immigrants (PRESARIO): Population-based matched cohort study | No natural experiment |
| Oppedal, B.; Idsoe, T. | 2015 | The role of social support in the acculturation and mental health of unaccompanied minor asylum seekers | No natural experiment |
| Miszkurka, M.; Goulet, L.; Zunzunegui, M. V.; Miszkurka, Malgorzata; Goulet, Lise; Zunzunegui, Maria Victoria | 2012 | Antenatal depressive symptoms among Canadian-born and immigrant women in Quebec: differential exposure and vulnerability to contextual risk factors | No natural experiment |
| Lumley, Mia; Katsikitis, Mary; Statham, Dixie | 2018 | Depression, Anxiety, and Acculturative Stress Among Resettled Bhutanese Refugees in Australia | No natural experiment |
| Maximova, K.; Krahn, H.; Maximova, Katerina; Krahn, Harvey | 2010 | Health status of refugees settled in Alberta: changes since arrival | No natural experiment |
| Lie, B. | 2002 | A 3-year follow-up study of psychosocial functioning and general symptoms in settled refugees | No natural experiment |
| Lau, Winnie; Silove, Derrick; Edwards, Ben; Forbes, David; Bryant, Richard; McFarlane, Alexander; Hadzi-Pavlovic, Dusan; Steel, Zachary; Nickerson, Angela; Van Hooff, Miranda; Felmingham, Kim; Cowlishaw, Sean; Alkemade, Nathan; Kartal, Dzenana; O'Donnell, Meaghan; O'Donnell, Meaghan | 2018 | Adjustment of refugee children and adolescents in Australia: outcomes from wave three of the Building a New Life in Australia study | No natural experiment |
| Kotey, Stanley; Carrico, Ruth; Wiemken, Timothy Lee; Furmanek, Stephen; Bosson, Rahel; Nyantakyi, Florence; VanHeiden, Sarah; Mattingly, William; Zierold, Kristina M. | 2018 | Elevated Blood Lead Levels by Length of Time From Resettlement to Health Screening in Kentucky Refugee Children | No natural experiment |

| | | | | |
|---|---|---|---|---|
| Lamkaddem, Majda; Essink-Bot, Marie-Louise; Devillé, Walter; Gerritsen, Annette; Stronks, Karien | 2015 | Health changes of refugees from Afghanistan, Iran and Somalia: the role of residence status and experienced living difficulties in the resettlement process | No natural experiment | |
| Melzer, Silvia Maja; Muffels, Ruud J | 2012 | Migrant's pursuit of happiness. The impact of adaption, social comparison and relative deprivation: Evidence from a" natural" experiment | No natural experiment | Claims to be natural experiment, but no random allocation |
| Koyama, S.; Aida, J.; Kawachi, I.; Kondo, N.; Subramanian, S. V.; Ito, K.; Kobashi, G.; Masuno, K.; Kondo, K.; Osaka, K. | 2014 | Social Support Improves Mental Health among the Victims Relocated to Temporary Housing following the Great East Japan Earthquake and Tsunami | No natural experiment | |
| Miranda, P. Y.; Schulz, A. J.; Israel, B. A.; Gonzalez, H. M. | 2011 | Context of Entry and Number of Depressive Symptoms in an Older Mexican-Origin Immigrant Population | No natural experiment | |
| Fritjers, P; Johnston, David W; Meng, Xin | 2009 | The mental health cost of long working hours: the case of rural Chinese migrants | No natural experiment | |
| Schooling, C Mary; Lam, Tai Hing; Ho, Sai Yin; Mak, Kwok Hang; Leung, Gabriel M | 2008 | Does economic development contribute to sex differences in ischaemic heart disease mortality? Hong Kong as a natural experiment using a case-control study | No natural experiment | Claims to be natural experiment, but no random allocation |
| Dong, Hao; Lee, James Z. | 2014 | Kinship matters: Long-term mortality consequences of childhood migration, historical evidence from northeast China, 1792-1909* | No natural experiment | |
| Giacco, Domenico; Bird, Victoria Jane; McCrone, Paul; Lorant, Vincent; Nicaise, Pablo; Pfennig, Andrea; Bauer, Michael; Ruggeri, Mirella; Lasalvia, Antonio; Moskalewicz, Jacek | 2015 | Specialised teams or personal continuity across inpatient and outpatient mental healthcare? Study protocol for a natural experiment | No natural experiment | Protocol |
| Hiday, V. A. | 1978 | Migration, urbanization, and fertility in the Philippines | No natural experiment | |
| Brewer, Mackenzie; Kimbro, Rachel Tolbert | 2014 | Neighborhood context and immigrant children's physical activity | No natural experiment | |
| Irfan, M. | 1986 | Migration and development in Pakistan: some selected issues | No natural experiment | |
| Hope, K. R. | 1989 | Managing rapid urbanization in the third world: some aspects of policy | No natural experiment | |
| Helliwell, John F; Bonikowska, Aneta; Shiplett, Hugh | 2016 | Migration as a test of the happiness set point hypothesis: Evidence from Immigration to Canada | No natural experiment | |
| Connolly, S.; O'Reilly, D. | 2007 | The contribution of migration to changes in the distribution of health over time: five-year follow-up study in Northern Ireland | No natural experiment | |
| O'Reilly, D.; Stevenson, M. | 2003 | Selective migration from deprived areas in Northern Ireland and the spatial distribution of inequalities: implications for monitoring health and inequalities in health | No natural experiment | |
| Correa-Velez, Ignacio; Gifford, Sandra M.; Barnett, Adrian G. | 2010 | Longing to belong: social inclusion and wellbeing among youth with refugee backgrounds in the first three years in Melbourne, Australia | No natural experiment | |
| Calderon, R. | 1983 | Inbreeding, migration and age at marriage in rural Toledo, Spain | No natural experiment | |
| Bozorgmehr, K.; Razum, O.; Szecsenyi, J.; Maier, W.; Stock, C. | 2017 | Regional deprivation is associated with the distribution of vulnerable asylum seekers: a nationwide small area analysis in Germany | No natural experiment | |
| Beiser, Morton; Hou, Feng | 2014 | Chronic health conditions, labour market participation and resource consumption among immigrant and native-born residents of Canada | No natural experiment | |
| Atkins, D. N.; Held, M. L.; Lindley, L. C. | 2018 | The impact of expanded health insurance coverage for unauthorized pregnant women on prenatal care utilization | No natural experiment | |
| Antai, D. | 2010 | Migration and child immunization in Nigeria: individual- and community-level contexts | No natural experiment | |

| | | | |
|---|---|---|---|
| Buchmueller, Thimo; Lembcke, Hanna; Busch, Julian; Kumsta, Robert; Wolf, Oliver T.; Leyendecker, Birgit | 2020 | Exploring hair steroid concentrations in asylum seekers, internally displaced refugees, and immigrants | No natural experiment |
| Haj-Younes, Jasmin; Stromme, Elisabeth Marie; Igland, Jannicke; Abildsnes, Eirik; Kumar, Bernadette; Hasha, Wegdan; Diaz, Esperanza | 2021 | Use of health care services among Syrian refugees migrating to Norway: a prospective longitudinal study | No natural experiment |
| Palladino, C.; Bello, J. M.; Perez-Hoyos, S.; Resino, R.; Guille, S.; Garcia, D.; Gurbindo, M. D.; Ramos, J. T.; de Jose, M. I.; Mellado, M. J.; Munoz-Fernadez, M. A. | 2008 | Spatial pattern of HIV-1 mother-to-child-transmission in Madrid (Spain) from 1980 till now: demographic and socioeconomic factors | No natural experiment |
| Parnia, A.; Chakravartty, D.; Wiseman, C. L. S.; Archbold, J.; Copes, R.; Zawar, N.; Chen, S. X.; Cole, D. C. | 2018 | Environmental factors associated with blood lead among newcomer women from South and East Asia in the Greater Toronto Area | No natural experiment |
| Lacey, K. K.; Park, J.; Briggs, A. Q.; Jackson, J. S. | 2019 | National origins, social context, timing of migration and the physical and mental health of Caribbeans living in and outside of Canada | No natural experiment |
| Lin, C.; Rodgers, Y. V. | 2019 | Social Disadvantage and Children's Nutritional Status in Rural-Urban Migrant Households | No natural experiment |
| Yalim, A. C. | 2020 | The Impacts of Contextual Factors on Psychosocial Wellbeing of Syrian Refugees: Findings from Turkey and the United States | No natural experiment |
| Koenig, W. D. | 1988 | Internal migration in the contemporary United States: comparison of measures and partitioning of stages | No natural experiment |
| Krishnaraj, M. | 1999 | Globalisation and women in India | No natural experiment |
| Kuumba, M. B. | 1993 | Perpetuating neo-colonialism through population control: South Africa and the United States | No natural experiment |
| Lawton, R. | 1987 | Peopling the past | No natural experiment |
| Lee, K.; Walt, G. | 1995 | Linking national and global population agendas: case studies from eight developing countries | No natural experiment |
| Levine, N. | 1980 | Antiurbanization: an implicit development policy in Turkey | No natural experiment |
| Lodha, R.; Dash, N. R.; Kapil, A.; Kabra, S. K. | 2000 | Diphtheria in urban slums in north India | No natural experiment |
| Ma, R. | 1993 | County town -- jian-zhi town differentials and migration to towns in China | No natural experiment |
| Mahmoud, M. E. | 1983 | Sudanese emigration to Saudi Arabia | No natural experiment |
| Meredith, W. H. | 1993 | China's family planning policy today | No natural experiment |
| Morauta, L.; Ryan, D. | 1982 | From temporary to permanent townsmen: migrants from the Malalaua District, Papua New Guinea | No natural experiment |
| Teitelbaum, M. S. | 1974 | Population and development: is a consensus possible? | No natural experiment |
| Thiesenhusen, W. C. | 1991 | Have agricultural economists neglected poverty issues? | No natural experiment |
| Trovato, F. | 1992 | Violent and accidental mortality among four immigrant groups in Canada, 1970-1972 | No natural experiment |
| Tsui, A. O.; Ragsdale, T. A.; Shirwa, A. I. | 1991 | The settlement of Somali nomads | No natural experiment |
| Rossini, R. E. | 1983 | Women as labor force in agriculture; the case of the State of S. Paulo, Brazil | No natural experiment |

| | | | | |
|---|---|---|---|---|
| Sathar, Z. A. | 1987 | Seeking explanations for high levels of infant mortality in Pakistan | No natural experiment | |
| Sathar, Z. A.; Kiani, M. F. | 1986 | Delayed marriages in Pakistan | No natural experiment | |
| Shami, S.; McCann, L. | 1993 | The social implications of population displacement and resettlement in the Middle East. Conference report | No natural experiment | |
| Shao, Cenyi; Meng, Xuehui; Cui, Shichen; Wang, Jingru; Li, Chengcheng | 2016 | Income-related health inequality of migrant workers in China and its decomposition: An analysis based on the 2012 China Labor-force Dynamics Survey data | No natural experiment | |
| Simon, J. L. | 1982 | A scheme to promote the world's economic development with migration | No natural experiment | |
| Wulf, D.; Willson, P. D. | 1984 | Global politics in Mexico City | No natural experiment | |
| Xu, B. | 1989 | Characteristics of population migration and measures to control in Beijing | No natural experiment | |
| Young, C. | 1992 | Pitfalls in comparing immigrants with the Australian-born population with particular reference to socioeconomic status | No natural experiment | |
| Koning, Pierre | 2016 | Privatizing sick pay: Does it work? | No natural experiment | |
| Kureková, Lucia | 2011 | From job search to skill search: Political economy of labor migration in central and Eastern Europe | No natural experiment | |
| Lee, Ronald; Miller, Timothy | 2000 | Immigration, social security, and broader fiscal impacts | No natural experiment | |
| Leon, David A | 2008 | Cities, urbanization and health | No natural experiment | |
| Lilford, Richard J; Oyebode, Oyinlola; Satterthwaite, David; Melendez-Torres, GJ; Chen, Yen-Fu; Mberu, Blessing; Watson, Samuel I; Sartori, Jo; Ndugwa, Robert; Caiaffa, Waleska | 2017 | Improving the health and welfare of people who live in slums | No natural experiment | |
| McKee, Martin; Reeves, Aaron; Clair, Amy; Stuckler, David | 2017 | Living on the edge: precariousness and why it matters for health | No natural experiment | |
| McKee, Martin; Stuckler, David | 2017 | The Deferred Action for Childhood Arrivals programme: a quasi-experiment in giving hope to migrants | No natural experiment | Commentary |
| McKenzie, David; Yang, Dean | 2010 | Experimental approaches in migration studies | No natural experiment | |
| McLaren, Lindsay; Hawe, Penelope | 2005 | Ecological perspectives in health research | No natural experiment | |
| Melzer, Silvia Maja | 2011 | Does Migration Make You Happy? The Influence of Migration on Subjective Well-Being | No natural experiment | |
| Neerup Handlos, Line; Fog Olwig, Karen; Bygbjerg, Ib Christian; Norredam, Marie | 2016 | Return Migrants' Experience of Access to Care in Corrupt Healthcare Systems: The Bosnian Example | No natural experiment | |
| Nunez-De La Mora, Alejandra; Chatterton, Robert T; Choudhury, Osul A; Napolitano, Dora A; Bentley, Gillian R | 2007 | Childhood conditions influence adult progesterone levels | No natural experiment | |
| Paloyo, Alfredo R; Reichert, Arndt R; Reuss-Borst, Monika; Tauchmann, Harald | 2015 | Who responds to financial incentives for weight loss? Evidence from a randomized controlled trial | No natural experiment | |
| Petticrew, Mark; Cummins, Steven; Ferrell, Catherine; Findlay, Anne; Higgins, Cassie; Hoy, Caroline; Kearns, Adrian; Sparks, Leigh | 2005 | Natural experiments: an underused tool for public health? | No natural experiment | Commentary |

| | | | | |
|---|---|---|---|---|
| Rellstab, Sara; Pecoraro, Marco; Holly, Alberto; Wanner, Philippe; Renard, Karine | 2016 | The Migrant Health Gap and the Role of Labour Market Status: Evidence from Switzerland | No natural experiment | |
| Reshmi, RS; Unisa, Sayeed; Jose, Juby Ann | 2014 | Women Migrants and their Mental Health: A Study of Working Women Hostellers in Mumbai | No natural experiment | |
| Rook, Graham A | 2013 | Regulation of the immune system by biodiversity from the natural environment: an ecosystem service essential to health | No natural experiment | |
| Sasin, Marcin J; McKenzie, David | 2007 | Migration, remittances, poverty, and human capital: conceptual and empirical challenges | No natural experiment | |
| López-Cevallos, Daniel F.; Chunhuei, Chi | 2012 | Migration, remittances, and health care utilization in Ecuador | No natural experiment | |
| Lee, H.; Brown, S. L.; Mitchell, M. M.; Schiraldi, G. R. | 2008 | Correlates of resilience in the face of adversity for Korean women immigrating to the US | No natural experiment | |
| Moh'd, Rabi'u Isah; Ajefu, Joseph Boniface | 2017 | Understanding the relationship between health and internal migration in the United Kingdom | No natural experiment | |
| Wilson, R. | 2001 | Asylum seekers: is there a proper housing response? | No natural experiment | |
| Rajamanoharan, S.; Monteiro, E. F.; Maw, R.; Carne, C. A.; Robinson, A.; Rajamanoharan, Sasikala; Monteiro, Eric F.; Maw, Raymond; Carne, Christopher A.; Robinson, Angela | 2004 | Genitourinary medicine/HIV services for persons with insecure immigration or seeking asylum in the United Kingdom: a British Co-operative Clinical Group survey | No natural experiment | |
| Tumen, Semih | 2015 | The use of natural experiments in migration research | No natural experiment | |
| Varheim, Andreas | 2014 | Trust and the role of the public library in the integration of refugees: The case of a Northern Norwegian city | No natural experiment | |
| Yang, Min; Dijst, Martin; Helbich, Marco | 2019 | Migration trajectories and their relationship to mental health among internal migrants in urban China: A sequence alignment approach | No natural experiment | |
| Newman, R. D.; Mnzava, A.; Szilagyi, Z. | 2013 | Mosquito larval source management: evaluating evidence in the context of practice and policy | No natural experiment | |
| Schooling, C Mary; Lam, Tai Hing; Thomas, G Neil; Cowling, Benjamin J; Heys, Michelle; Janus, Edward D; Leung, Gabriel M; Hong Kong Cardiovascular Risk Factor Prevalence Study Steering Committee | 2007 | Growth environment and sex differences in lipids, body shape and diabetes risk | No natural experiment | Claims to be natural experiment, but no random allocation |
| Choi, J. Y. | 2009 | Contextual effects on health care access among immigrants: lessons from three ethnic communities in Hawaii | No natural experiment | |
| Shelley, D.; Fahs, M.; Yerneni, R.; Das, D.; Nguyen, N.; Hung, D.; Burton, D.; Chin, M.; Chang, M. D.; Cummings, K. M.; Shelley, Donna; Fahs, Marianne; Yerneni, Rajeev; Das, Dhiman; Nguyen, Nam; Hung, Dorothy; Burton, Dee; Chin, Margaret; Chang, Ming-der; Cummings, K. Michael | 2008 | Effectiveness of tobacco control among Chinese Americans: a comparative analysis of policy approaches versus community-based programs | No natural experiment | claims to be natural experiment, but is an intervention study |
| Melzer, Silvia; Muffels, Ruud | 2012 | Migrant's Pursuit of Happiness-The Impact of Adaptation, Social Comparison and Relative Deprivation: Evidence from a Natural Experiment | No natural experiment | Duplicate |
| Khosravi, Roghayeh; Azman, Azlinda; Ayasreh, Emad Abdallah Mustafa; Khosravi, Sahar | 2019 | Can a building social capital intervention improve the mental health of international students? A non-randomized quasi-experimental study | No natural experiment | Claims to be natural experiment, but Intervention is designed by researchers |
| Astell-Burt, Thomas; Feng, Xiaoqi; Kolt, Gregory S | 2016 | Large-scale investment in green space as an intervention for physical activity, mental and cardiometabolic health: study protocol for a quasi-experimental evaluation of a natural experiment | No natural experiment | Protocol |

| | | | |
|---|---|---|---|
| Kim, Jungho | 2016 | Hängen Bildungsstand und Geburtenhäufigkeit von Frauen zusammen? | No natural experiment |
| Kanchanachitra, C.; Lindelow, M.; Johnston, T.; Hanvoravongchai, P.; Lorenzo, F. M.; Huong, N. L.; Wilopo, S. A.; Dela Rosa, J. F. | 2011 | Human resources for health in southeast Asia: shortages, distributional challenges, and international trade in health services | No natural experiment |
| Islam, M. N. | 1983 | The efficiency of interprovincial migration in Canada, 1961-1978 | No natural experiment |
| Havinga, I. C.; Mohammad, F.; Cohen, S. I. | 1986 | Intergenerational mobility and long-term socio-economic change in Pakistan | No natural experiment |
| Hatzenbuehler, Mark L | 2010 | Social factors as determinants of mental health disparities in LGB populations: Implications for public policy | No natural experiment |
| Hammar, N; Kaprio, J; Hagström, U; Alfredsson, L; Koskenvuo, Markku; Hammar, T | 2002 | Migration and mortality: a 20 year follow up of Finnish twin pairs with migrant co-twins in Sweden | No natural experiment |
| Green, S. C. | 1978 | Migrant adjustment in Seoul, Korea: employment and housing | No natural experiment |
| Grant, M. | 1995 | Movement patterns and the medium-sized city. Tenants on the move in Gweru, Zimbabwe | No natural experiment |
| Kim, D. S. | 1994 | The demographic transition in the Korean peninsula, 1910-1990: South and North Korea compared | No natural experiment |
| Khlat, Myriam; Guillot, Michel | 2017 | Health and mortality patterns among migrants in France | No natural experiment |
| Khan, Z. | 1991 | Are breastfeeding patterns in Pakistan changing? | No natural experiment |
| Khalaf, M. | 1993 | The Lebanese woman and the labor market | No natural experiment |
| Kauppi, Carol; Forchuk, Cheryl; Montgomery, Phyllis; Edwards, Betty; Davie, Samantha; Rudnick, Abraham | 2015 | Migration, Homelessness, and Health Among Psychiatric Survivors in Northern and Southern Ontario | No natural experiment |
| Kaprio, Jaakko; Koskenvuo, Markku | 2002 | Genetic and environmental factors in complex diseases: the older Finnish Twin Cohort | No natural experiment |
| Huijts, Tim; Kraaykamp, Gerbert | 2012 | Immigrants' health in Europe: a cross-classified multilevel approach to examine origin country, destination country, and community effects | No natural experiment |
| Janevic, T.; Borrell, L. N.; Savitz, D. A.; Echeverria, S. E.; Rundle, A. | 2014 | Ethnic enclaves and gestational diabetes among immigrant women in New York City | No natural experiment |
| Johnson, M. A.; Marchi, K. S. | 2009 | Segmented assimilation theory and perinatal health disparities among women of Mexican descent | No natural experiment |
| Dunlavy, A. C.; Garcy, A. M.; Rostila, M. | 2016 | Educational mismatch and health status among foreign-born workers in Sweden | No natural experiment |
| Chen, Juan; Chen, Shuo; Landry, Pierre F. | 2013 | Migration, environmental hazards, and health outcomes in China | No natural experiment |
| Chen, Juan | 2011 | Internal migration and health: Re-examining the healthy migrant phenomenon in China | No natural experiment |
| Cabieses, Báltica; Uphoff, Eleonora; Pinart, Mariona; Antó, Josep Maria; Wright, John | 2014 | A systematic review on the development of asthma and allergic diseases in relation to international immigration: the leading role of the environment confirmed | No natural experiment |
| Halliday, F. | 1982 | Labour migration in the Arab world: the ugly face of the new economic order | No natural experiment |

| | | | |
|---|---|---|---|
| Grzywacz, J. G.; Quandt, S. A.; Chen, H.; Isom, S.; Kiang, L.; Vallejos, Q.; Arcury, T. A.; Grzywacz, Joseph G.; Quandt, Sara A.; Chen, Haiying; Isom, Scott; Kiang, Lisa; Vallejos, Quirina; Arcury, Thomas A. | 2010 | Depressive symptoms among Latino farmworkers across the agricultural season: Structural and situational influences | No natural experiment |
| Giulietti, Corrado | 2014 | The welfare magnet hypothesis and the welfare take-up of migrants | No natural experiment |
| Kim, Karen; Quinn, Michal; Lam, Helen | 2018 | Promoting Colorectal Cancer Screening in Foreign-Born Chinese-American Women: Does Racial/Ethnic and Language Concordance Matter? | No natural experiment |
| Kaucher, S.; Deckert, A.; Becher, H.; Winkler, V. | 2017 | Migration pattern and mortality of ethnic German migrants from the former Soviet Union: a cohort study in Germany | No natural experiment |
| Jensen, T. K.; Skardalsmo, E. M. B.; Fjermestad, K. W. | 2014 | Development of mental health problems - a follow-up study of unaccompanied refugee minors | No natural experiment |
| Hauff, E.; Vaglum, P. | 1995 | ORGANIZED VIOLENCE AND THE STRESS OF EXILE - PREDICTORS OF MENTAL-HEALTH IN A COMMUNITY COHORT OF VIETNAMESE REFUGEES 3 YEARS AFTER RESETTLEMENT | No natural experiment |
| Gresenz, C. R.; Derose, K. P.; Ruder, T.; Escarce, J. J. | 2012 | Health Care Experiences of Hispanics in New and Traditional US Destinations | No natural experiment |
| Garg, Teevrat | 2019 | Ecosystems and human health: The local benefits of forest cover in Indonesia | No natural experiment |
| Gandhi, Mamta; Narang, Kavita; Kaur, Manmeet | 2014 | Gaps in availability, utilization and expectations of people from health care services: A study of resettlement colony, Chandigarh | No natural experiment |
| Gallart, Albert; Cruz, Félix; Zabalegui, Adelaida | 2013 | Factors influencing burden among non-professional immigrant caregivers: a case-control study | No natural experiment |
| Gakuba, Théogène-Octave | 2015 | Young african refugees in urban context (abidjan, dakar, geneva) psychosocial aspects and resilience | No natural experiment |
| Faskunger, J.; Eriksson, U.; Johansson, S. E.; Sundquist, K.; Sundquist, J. | 2009 | Risk of obesity in immigrants compared with Swedes in two deprived neighbourhoods | No natural experiment |
| Dustmann, Christian; Weiss, Yoram | 2007 | Return migration: theory and empirical evidence from the UK | No natural experiment |
| De, Prabal K; Dench, Daniel | 2016 | Mental Health Effects of Internal Migration - Evidence from Urban Bangladesh | No natural experiment |
| Das, Jishnu; Do, Quy-Toan; Friedman, Jed; McKenzie, David; Scott, Kinnon | 2007 | Mental health and poverty in developing countries: Revisiting the relationship | No natural experiment |
| Das, Jishnu; Do, Quy-Toan; Friedman, Jed; McKenzie, David | 2009 | Mental health patterns and consequences: results from survey data in five developing countries | No natural experiment |
| Cooke, Thomas J; Speirs, Karen | 2005 | Migration and employment among the civilian spouses of military personnel | No natural experiment |
| Choe, E. H. | 1985 | Volume and stream of migrants from the National Migration Survey | No natural experiment |
| Bhalotra, Sonia; Clots-Figueras, Irma | 2014 | Health and the political agency of women | No natural experiment |
| Besley, Timothy; Kudamatsu, Masayuki | 2006 | Health and democracy | No natural experiment |
| Barghadouch, A.; Carlsson, J.; Norredam, M. | 2018 | Psychiatric Disorders and Predictors Hereof Among Refugee Children in Early Adulthood: A Register-Based Cohort Study | No natural experiment |
| Banerjee, B. | 1983 | Social networks in the migration process: empirical evidence on chain migration in India | No natural experiment |

| | | | |
|---|---|---|---|
| Gelbard, A.; Haub, C.; Kent, M. M. | 1999 | World population beyond six billion | No natural experiment |
| Loret de Mola, C.; Stanojevic, S.; Ruiz, P.; Gilman, R. H.; Smeeth, L.; Miranda, J. J.; Loret de Mola, Christian; Stanojevic, Sanja; Ruiz, Paulo; Gilman, Robert H.; Smeeth, Liam; Miranda, J. Jaime | 2012 | The effect of rural-to-urban migration on social capital and common mental disorders: PERU MIGRANT study | No natural experiment |
| Hrabovszky, J. P.; Miyan, K. | 1987 | Population growth and land use in Nepal: "the great turnabout" | No natural experiment |
| Gee, Gilbert C.; de Castro, A. B.; Wang, May C.; Crespi, Catherine M.; Morey, Brittany N.; Kaori, Fujishiro | 2015 | Feasibility of Conducting a Longitudinal, Transnational Study of Filipino Migrants to the United States: A Dual-Cohort Design | No natural experiment |
| Gakuba, Théogène-Octave; Sall, Mohamadou; Fokou, Gilbert; Kouakou, Christiane; Amalaman, Martin; Kone, Solange | 2015 | Mental Health and Resilience of Young African Women Refugees in Urban Context (Abidjan - Ivory Coast and Daka - Senegal) | No natural experiment |
| Fritz, M. V.; Chin, D.; DeMarinis, V. | 2008 | Stressors, anxiety, acculturation and adjustment among international and North American students | No natural experiment |
| Evlampidou, I.; Danis, K.; Lenglet, A.; Tseroni, M.; Theocharopoulos, Y.; Panagiotopoulos, T. | 2015 | Malaria knowledge, attitudes and practices among migrants from malaria-endemic countries in Evrotas, Laconia, Greece, 2013 | No natural experiment |
| Erickson, P. I. | 1994 | Lessons from a repeat pregnancy prevention program for Hispanic teenage mothers in east Los Angeles | No natural experiment |
| Edwards, D. B. | 1986 | Marginality and migration: cultural dimensions of the Afghan refugee problem | No natural experiment |
| Eastwood, D. A. | 1983 | Reality of delusion: migrant perception of levels of living and opportunity in Venezuela, 1961-1971 | No natural experiment |
| Cebotari, Victor; Siegel, Melissa; Mazzucato, Valentina | 2018 | Migration and child health in Moldova and Georgia | No natural experiment |
| Ahearn, F. L.; Noble, J. H. | 2004 | Post-civil war adaptation and need in Managua, Nicaragua | No natural experiment |
| Si, Si; Peters, Susan; Reid, Alison | 2018 | ariations in mesothelioma mortality rates among migrants to Australia and Australian-born | No natural experiment |
| Rasmussen, A.; Annan, J. | 2010 | Predicting Stress Related to Basic Needs and Safety in Darfur Refugee Camps: A Structural and Social Ecological Analysis | No natural experiment |
| Randell, H. | 2016 | The short-term impacts of development-induced displacement on wealth and subjective well-being in the Brazilian Amazon | No natural experiment |
| Nishikiori, N.; Abe, T.; Costa, D. G. M.; Dharmaratne, S. D.; Kunii, O.; Moji, K. | 2006 | Who died as a result of the tsunami? Risk factors of mortality among internally displaced persons in Sri Lanka: a retrospective cohort analysis | No natural experiment |
| Ro, Annie; Fleischer, Nancy L.; Blebu, Bridgette | 2016 | An examination of health selection among U.S. immigrants using multi-national data | No natural experiment |
| Jones, Cheryl B; Sherwood, Gwen | 2014 | The globalization of the nursing workforce: Pulling the pieces together | No natural experiment |
| Um, M. Y.; Chi, I.; Kim, H. J.; Palinkas, L. A.; Kim, J. Y. | 2015 | Correlates of depressive symptoms among North Korean refugees adapting to South Korean society: The moderating role of perceived discrimination | No natural experiment |
| Termorshuizen, Fabian; Heerdink, Eibert R.; Selten, Jean-Paul | 2018 | The impact of ethnic density on dispensing of antipsychotic and antidepressant medication among immigrants in the Netherlands | No natural experiment |
| Steel, Zachary; Momartin, Shakeh; Silove, Derrick; Coello, Marianio; Aroche, Jorge; Tay, Kuo Wei | 2011 | Two year psychosocial and mental health outcomes for refugees subjected to restrictive or supportive immigration policies | No natural experiment |
| Raude, J.; Setbon, M. | 2009 | The role of environmental and individual factors in the social epidemiology of chikungunya disease on Mayotte Island | No natural experiment |

| | | | |
|---|---|---|---|
| Rees, Susan J.; Fisher, Jane R.; Steel, Zachary; Mohsin, Mohammed; Nadar, Nawal; Moussa, Batool; Hassoun, Fatima; Yousif, Mariam; Krishna, Yalini; Khalil, Batoul; Mugo, Jok; Tay, Alvin Kuowei; Klein, Louis; Silove, Derrick | 2019 | Prevalence and Risk Factors of Major Depressive Disorder Among Women at Public Antenatal Clinics From Refugee, Conflict-Affected, and Australian-Born Backgrounds | No natural experiment |
| Remennick, L. I. | 2002 | Immigrants from Chernobyl-affected areas in Israel: the link between health and social adjustment | No natural experiment |
| Donato, K. M.; Kanaiaupuni, S. M.; Stainback, M. | 2003 | Sex differences in child health: Effects of Mexico-US migration | No natural experiment |
| Dona, G. | 2010 | Rethinking well-being: from contexts to processes | No natural experiment |
| Desai, V. | 1994 | Migration and labour characteristics of slum dwellers in Bombay | No natural experiment |
| Daw, M. A.; El-Bouzedi, A.; Ahmed, M. O.; Dau, A. A.; Agnan, M. M.; Libyan Study Grp, Hepatitis; Hiv, | 2016 | Epidemiology of hepatitis C virus and genotype distribution in immigrants crossing to Europe from North and sub-Saharan Africa | No natural experiment |
| Crofts, J. P.; Gelb, D.; Andrews, N.; Delpech, V.; Watson, J. M.; Abubakar, I. | 2008 | Investigating tuberculosis trends in England | No natural experiment |
| Liu, Lee | 2019 | China's dusty lung crisis: Rural-urban health inequity as social and spatial injustice | No natural experiment |
| Macfarlane, A.; Singleton, C.; Green, E. | 2009 | Language barriers in health and social care consultations in the community: A comparative study of responses in Ireland and England | No natural experiment |
| Meleis, A. I. | 1979 | The health care system of Kuwait: the social paradoxes | No natural experiment |
| Cruz, M.; D'Ayala, P. G.; Marcus, E.; McElroy, J. L.; Rossi, O. | 1987 | The demographic dynamics of small island societies | No natural experiment |
| Collinson, Mark A; White, Michael J; Bocquier, Philippe; McGarvey, Stephen T; Afolabi, Sulaimon A; Clark, Samuel J; Kahn, Kathleen; Tollman, Stephen M | 2014 | Migration and the epidemiological transition: insights from the Agincourt sub-district of northeast South Africa | No natural experiment |
| Connolly, S.; Rosato, M.; O'Reilly, D. | 2011 | The effect of population movement on the spatial distribution of socio-economic and health status: Analysis using the Northern Ireland mortality study | No natural experiment |
| Collinson, M. A.; Tollman, S. M.; Kahn, K. | 2007 | Migration, settlement change and health in post-apartheid South Africa: triangulating health and demographic surveillance with national census data | No natural experiment |
| Cleveland, Janet; Rousseau, CeÌ• cile; Kronick, Rachel | 2012 | The harmful effects of detention and family separation on asylum seekers' mental health in the context of Bill C-31 | No natural experiment |
| Clemens, Michael A; Özden, Çağlar; Rapoport, Hillel | 2015 | Reprint of: Migration and development research is moving far beyond remittances | No natural experiment |
| Clarke, J. I. | 1969 | Population policies and dynamics in Tunisia | No natural experiment |
| Clark, S. J.; Collinson, M. A.; Kahn, K.; Drullinger, K.; Tollman, S. M. | 2007 | Returning home to die: circular labour migration and mortality in South Africa | No natural experiment |
| Christopher, K. A. | 1998 | Determinants of psychological well-being in immigrants | No natural experiment |
| Chen, Joyce; Kosec, Katrina; Mueller, Valerie | 2019 | Moving to despair? Migration and well-being in Pakistan | No natural experiment |
| Chen, C.; Liu, S. F. | 1997 | Migration into and out of Taiwan, 1895-1944 | No natural experiment |

| | | | |
|---|---|---|---|
| Cayuela, A.; Martinez, J. M.; Ronda, E.; Delclos, G. L.; Conway, S. | 2018 | Assessing the influence of working hours on general health by migrant status and family structure: the case of Ecuadorian-, Colombian-, and Spanish-born workers in Spain | No natural experiment |
| Ashford, L. S.; Haws, J. M. | 1992 | Family planning program sustainability: threat or opportunity? | No natural experiment |
| Ambler, Kate | 2019 | Migration and remittances in Central America: New evidence and pathways for future research | No natural experiment |
| Alter, George; Oris, Michel | 2005 | Childhood conditions, migration, and mortality: Migrants and natives in 19th-• century cities | No natural experiment |
| Alati, Rosa; Najman, Jake M; Shuttlewood, Gregory J; Williams, Gail M; Bor, William | 2003 | Changes in mental health status amongst children of migrants to Australia: a longitudinal study | No natural experiment |
| Alcantara, C.; Chen, C. N.; Alegria, M. | 2014 | Do post-migration perceptions of social mobility matter for Latino immigrant health? | No natural experiment |
| Åkerman, Eva; Larsson, Elin C.; Essén, Birgitta; Westerling, Ragnar | 2019 | A missed opportunity? Lack of knowledge about sexual and reproductive health services among immigrant women in Sweden | No natural experiment |
| Adedeji, A.; Bullinger, Monika | 2019 | Subjective integration and quality of life of Sub-Saharan African migrants in Germany | No natural experiment |
| Acevedo-Garcia, Dolores; Almeida, Joanna | 2012 | Special issue introduction: place, migration and health | No natural experiment |
| Koehn, P. H. | 2006 | Transnational migration, state policy and local clinician treatment of asylum seekers and resettled migrants: comparative perspectives on reception center and community health care practice in Finland | No natural experiment |
| Kim, Isok | 2016 | Beyond Trauma: Post-resettlement Factors and Mental Health Outcomes Among Latino and Asian Refugees in the United States | No natural experiment |
| Kendall, Jacob; Anglewicz, Philip | 2017 | Characteristics Associated With Migration Among Older Women and Men in Rural Malawi | No natural experiment |
| Kauhl, B.; Maier, W.; Schweikart, J.; Keste, A.; Moskwyn, M. | 2018 | Who is where at risk for Chronic Obstructive Pulmonary Disease? A spatial epidemiological analysis of health insurance claims for COPD in Northeastern Germany | No natural experiment |
| Kaestner, R.; Malamud, O. | 2014 | SELF-SELECTION AND INTERNATIONAL MIGRATION: NEW EVIDENCE FROM MEXICO | No natural experiment |
| Joy, Edward J. M.; Green, Rosemary; Agrawal, Sutapa; Aleksandrowicz, Lukasz; Bowen, Liza; Kinra, Sanjay; Macdiarmid, Jennie I.; Haines, Andy; Dangour, Alan D. | 2017 | Dietary patterns and non-communicable disease risk in Indian adults: secondary analysis of Indian Migration Study data | No natural experiment |
| Jarallah, Y.; Baxter, J. | 2019 | Gender disparities and psychological distress among humanitarian migrants in Australia: a moderating role of migration pathway? | No natural experiment |
| Husain, F.; Anderson, M.; Lopes Cardozo, B.; Becknell, K.; Blanton, C.; Araki, D.; Vithana, E. K.; Husain, Farah; Anderson, Mark; Lopes Cardozo, Barbara; Becknell, Kristin; Blanton, Curtis; Araki, Diane; Vithana, Eeshara Kottegoda | 2011 | Prevalence of war-related mental health conditions and association with displacement status in postwar Jaffna District, Sri Lanka | No natural experiment |
| Jang, Yuri; Yoon, Hyunwoo; Rhee, Min-• Kyoung; Park, Nan Sook; Chiriboga, David A.; Kim, Miyong T. | 2019 | Factors associated with dental service use of older Korean Americans | No natural experiment |
| Im, Hyuk; Lee, Ki; Lee, Hyo | 2014 | Acculturation Stress and Mental Health Among the Marriage Migrant Women in Busan, South Korea | No natural experiment |
| Huq-hussain, S. | 1995 | Fighting poverty: the economic adjustment of female migrants in Dhaka | No natural experiment |

| | | | |
|---|---|---|---|
| Hulanicka, B.; Gronkiewicz, L.; Zietkiewicz, B. | 1999 | Stature of boys post World War II migrants | No natural experiment |
| Hemphill, E.; Raine, K.; Spence, J. C.; Smoyer-Tomic, K. E. | 2008 | Exploring obesogenic food environments in Edmonton, Canada: The association between socioeconomic factors and fast-food outlet access | No natural experiment |
| Hammarstedt, M. | 2000 | The receipt of transfer payments by immigrants in Sweden | No natural experiment |
| Hajizadeh, M.; Campbell, M. K.; Sarma, S. | 2016 | A Spatial Econometric Analysis of Adult Obesity: Evidence from Canada | No natural experiment |
| Guo, Man; Sabbagh Steinberg, Nadia; Dong, Xinqi; Tiwari, Agnes | 2019 | Is family relations related to health service utilisation among older immigrants: Evidence from Chinese elderly in the United States | No natural experiment |
| Greenwood, M. J. | 1971 | An analysis of the determinants of internal labor mobility in India | No natural experiment |
| Greenwood, M. J. | 1969 | The determinants of labor migration in Egypt | No natural experiment |
| Danke, K.; Blecher, C.; Bardehle, D.; Cremer, D.; Razum, O. | 2008 | Small Area Analysis of Infant Mortality in Bielefeld with Special Consideration of the Migration Satutus of Parents, 2000-2006 | No natural experiment |
| Graves, P. E. | 1983 | Migration with a composite amenity: the role of rents | No natural experiment |
| Frenkel, S. J.; Li, M.; Restubog, S. L. D. | 2012 | Management, Organizational Justice and Emotional Exhaustion among Chinese Migrant Workers: Evidence from two Manufacturing Firms | No natural experiment |
| De Grande, Hannelore; Vandenheede, Hadewijch; Gadeyne, Sylvie; Deboosere, Patrick | 2014 | Health status and mortality rates of adolescents and young adults in the Brussels-Capital Region: differences according to region of origin and migration history | No natural experiment |
| Creatore, M. I.; Moineddin, R.; Booth, G.; Manuel, D. H.; DesMeules, M.; McDermott, S.; Glazier, R. H. | 2010 | Age- and sex-related prevalence of diabetes mellitus among immigrants to Ontario, Canada | No natural experiment |
| Crawshaw, Alison F.; Pareek, Manish; Were, John; Schillinger, Steffen; Gorbacheva, Olga; Wickramage, Kolitha P.; Mandal, Sema; Delpech, Valerie; Gill, Noel; Kirkbride, Hilary; Zenner, Dominik | 2018 | Infectious disease testing of UK-bound refugees: a population-based, cross-sectional study | No natural experiment |
| Cooper, S.; Enticott, J. C.; Shawyer, F.; Meadows, G. | 2019 | Determinants of Mental Illness Among Humanitarian Migrants: Longitudinal Analysis of Findings From the First Three Waves of a Large Cohort Study | No natural experiment |
| Choi, H.; Rauh, V.; Garfinkel, R.; Tu, Y.; Perera, F. P. | 2008 | Prenatal exposure to airborne polycyclic aromatic hydrocarbons and risk of intrauterine growth restriction | No natural experiment |
| Chen, X. M.; Zhang, Q.; Wang, J. Y.; Liu, J.; Zhang, W. B.; Qi, S.; Xu, H.; Li, C.; Zhang, J. S.; Zhao, H. T.; Meng, S. S.; Li, D.; Lu, H. Y.; Aschner, M.; Li, B.; Yin, H.; Chen, J. Y.; Luo, W. J. | 2017 | Cognitive and neuroimaging changes in healthy immigrants upon relocation to a high altitude: A panel study | No natural experiment |
| Buu, A.; Mansour, M.; Wang, J.; Refior, S. K.; Fitzgerald, H. E.; Zucker, R. A. | 2007 | Alcoholism effects on social migration and neighborhood effects on alcoholism over the course of 12 years | No natural experiment |
| Zhu, L.; Xu, P. | 2015 | The Politics of Welfare Exclusion: Immigration and Disparity in Medicaid Coverage | No natural experiment |
| Zhou, G. F.; Lo, E.; Zhong, D. B.; Wang, X. M.; Wang, Y.; Malla, S.; Lee, M. C.; Yang, Z. Q.; Cui, L. W.; Yan, G. Y. | 2016 | Impact of interventions on malaria in internally displaced persons along the China-Myanmar border: 2011-2014 | No natural experiment |
| Zheng, Y.; Lamoureux, E. L.; Ikram, M. K.; Mitchell, P.; Wang, J. J.; Younan, C.; Anuar, A. R.; Tai, E. S.; Wong, T. Y. | 2012 | Impact of migration and acculturation on prevalence of type 2 diabetes and related eye complications in indians living in a newly urbanised society | No natural experiment |

| | | | |
|---|---|---|---|
| Gee, Gilbert C.; de Castro, A. B.; Crespi, Catherine M.; Wang, May C.; Llave, Karen; Brindle, Eleanor; Lee, Nanette R.; Kabamalan, Maria Midea M.; Hing, Anna K. | 2018 | Health of Philippine Emigrants Study (HoPES): study design and rationale | No natural experiment |
| Ikhile, Ifunanya; Anderson, Claire; McGrath, Simon; Bridges, Stephanie | 2018 | Is the Global Pharmacy Workforce Issue All About Numbers? | No natural experiment |
| Zarulli, V. | 2016 | Post-War Migration Flows and Disparities in Mortality from Age 50 Years Onwards: the Case of Turin in Italy | No natural experiment |
| Yuan, Si-Yang; Freeman, Ruth | 2011 | Can social support in the guise of an oral health education intervention promote mother-"infant bonding in Chinese immigrant mothers and their infants? | No natural experiment |
| Beiser, Morton; Puente-Duran, Sofia; Hou, Feng | 2015 | Cultural distance and emotional problems among immigrant and refugee youth in Canada: Findings from the New Canadian Child and Youth Study (NCCYS) | No natural experiment |
| Beiser, M.; Zilber, N.; Simich, L.; Youngmann, R.; Zohar, A. H.; Taa, B.; Hou, F. | 2011 | Regional effects on the mental health of immigrant children: Results from the New Canadian Children and Youth Study (NCCYS) | No natural experiment |
| Arthur, J. A. | 1991 | Interregional migration of labor in Ghana, West Africa: determinants, consequences and policy intervention | No natural experiment |
| Xiao, Y. Y.; Zhao, N. Q.; Yu, M.; Zhao, M.; Zhong, J. M.; Gong, W. W.; Hu, R. Y. | 2013 | Factors Associated with Severe Deliberate Self-Harm among Chinese Internal Migrants | No natural experiment |
| Woods, R. | 1984 | Population studies | No natural experiment |
| Wild, Verina | 2012 | Migration and Health: Discovering New Territory for Bioethics | No natural experiment |
| Wen, Ming; Maloney, Thomas N. | 2014 | Neighborhood socioeconomic status and BMI differences by immigrant and legal status: evidence from Utah | No natural experiment |
| Wanna, C. P.; Seehuus, M.; Mazzulla, E.; Fondacaro, K. | 2019 | A house is not a home: Modeling the effects of social support and connection within resettled refugee populations | No natural experiment |
| Wang, Bo-Ram; Kwon, Young Dae; Jeon, Wootack; Noh, Jin-Won | 2015 | Factors associated with the frequency of physician visits among North Korean defectors residing in South Korea: a cross-sectional study | No natural experiment |
| Walton, E. | 2015 | Making Sense of Asian American Ethnic Neighborhoods: A Typology and Application to Health | No natural experiment |
| Waldinger, R. | 1989 | Immigration and urban change | No natural experiment |
| Wakabayashi, K. | 1990 | Migration from rural to urban areas in China | No natural experiment |
| Wai, Khin Thet; Kyaw, Myat Phone; Oo, Tin; Zaw, PeThet; Nyunt, Myat Htut; Thida, Moe; Kyaw, Thar Tun | 2014 | Spatial distribution, work patterns, and perception towards malaria interventions among temporary mobile/migrant workers in artemisinin resistance containment zone | No natural experiment |
| Vearey, Joanna | 2012 | Learning from HIV: Exploring migration and health in South Africa | No natural experiment |
| Van Praag, B. M. | 1988 | The notion of population economics | No natural experiment |
| Urquia, M. L.; Frank, J. W.; Glazier, R. H.; Moineddin, R.; Matheson, F. I.; Gagnon, A. J. | 2009 | Neighborhood Context and Infant Birthweight Among Recent Immigrant Mothers: A Multilevel Analysis | No natural experiment |
| Tseliou, F.; Maguire, A.; Donnelly, M.; O'Reilly, D. | 2016 | The impact of childhood residential mobility on mental health outcomes in adolescence and early adulthood: a record linkage study | No natural experiment |
| Thomas, Duncan; Frankenberg, Elizabeth | 2002 | Health, nutrition and prosperity: a microeconomic perspective | No natural experiment |

| | | | |
|---|---|---|---|
| Tunstall, H.; Shortt, N. K.; Pearce, J. R.; Mitchell, R. J. | 2015 | Difficult Life Events, Selective Migration and Spatial Inequalities in Mental Health in the UK | No natural experiment |
| Tsoupakis, E.; Tziafetas, G. | 1983 | Statistical analysis of the immigration flow in Greece | No natural experiment |
| Tozer, M.; Khawaja, N. G.; Schweitzer, R. | 2018 | Protective Factors Contributing to Wellbeing Among Refugee Youth in Australia | No natural experiment |
| Torres, Jacqueline M; Casey, Joan A | 2017 | The centrality of social ties to climate migration and mental health | No natural experiment |
| Tessaring, M. | 1988 | Demographic aspects of educational expansion and labour-force development in the Federal Republic of Germany | No natural experiment |
| Svendsen, Erik R; Runkle, Jennifer R; Dhara, Venkata Ramana; Lin, Shao; Naboka, Marina; Mousseau, Timothy A; Bennett, Charles L | 2012 | Epidemiologic methods lessons learned from environmental public health disasters: Chernobyl, the World Trade Center, Bhopal, and Graniteville, South Carolina | No natural experiment |
| Suwan, P.; Grossman, J. | 1987 | Assessment of primary health care resources in Thailand | No natural experiment |
| Short, Susan E; Mollborn, Stefanie | 2015 | Social determinants and health behaviors: conceptual frames and empirical advances | No natural experiment |
| Satterthwaite, D. | 1993 | The impact on health of urban environments | No natural experiment |
| Razum, Oliver; Breckenkamp, Jürgen; Fauser, Margit | 2019 | Transnational ties, endowment with capital, and health of immigrants in Germany: cross-sectional study | No natural experiment |
| Probst, Janice C; Moore, Charity G; Glover, Saundra H; Samuels, Michael E | 2004 | Person and place: the compounding effects of race/ethnicity and rurality on health | No natural experiment |
| Piwowarczyk, L.; Keane, T. M.; Lincoln, A. | 2008 | Hunger: The silent epidemic among asylum seekers and resettled refugees | No natural experiment |
| Petrelli, Alessio; Di Napoli, Anteo; Rossi, Alessandra; Costanzo, Gianfranco; Mirisola, Concetta; Gargiulo, Lidia | 2017 | The variation in the health status of immigrants and Italians during the global crisis and the role of socioeconomic factors | No natural experiment |
| Perez-Carceles, Maria D.; Medina, Maria D.; Perez-Flores, Domingo; Noguera, Jose A.; Pereniguez, Juan E.; Madrigal, Manuel; Luna, Aurelio | 2014 | Screening for hazardous drinking in migrant workers in southeastern Spain | No natural experiment |
| Brunswic, E. | 1993 | Editorial | No natural experiment |
| Lee, C. I.; Smith, L. S.; Shwe Oo, E. K.; Scharschmidt, B. C.; Whichard, E.; Kler, T.; Lee, T. J.; Richards, A. K. | 2009 | Internally displaced human resources for health: villager health worker partnerships to scale up a malaria control programme in active conflict areas of eastern Burma | No natural experiment |
| Nielsen, Signe Smith; Hempler, Nana Folmann; Krasnik, Allan | 2013 | Issues to consider when measuring and applying socioeconomic position quantitatively in immigrant health research | No natural experiment |
| Money, J. | 1997 | No vacancy: the political geography of immigration control in advanced industrial countries | No natural experiment |
| Monasta, L.; Andersson, N.; Ledogar, R. J.; Cockcroft, A. | 2008 | Minority health and small numbers epidemiology: a case study of living conditions and the health of children in 5 foreign Romá camps in Italy | No natural experiment |
| Milas, S. | 1984 | Population crisis and desertification in the Sudano-Sahelian region | No natural experiment |
| Merkin, Sharon Stein; Arditi-Babchuk, Hadar; Shohat, Tamy | 2015 | Neighborhood socioeconomic status and self-rated health in Israel: the Israel National Health Interview Survey | No natural experiment |
| Ackermann Rau, Sabine; Sakarya, Sibel; Abel, Thomas | 2014 | When to see a doctor for common health problems: distribution patterns of functional health literacy across migrant populations in Switzerland | No natural experiment |

| | | | | |
|---|---|---|---|---|
| Mellor, J. W. | 1991 | Agricultural links to nonagricultural growth: urbanization, employment, poverty | No natural experiment | |
| McKee, Martin; Stuckler, David | 2018 | Revisiting the corporate and commercial determinants of health | No natural experiment | |
| Marshall, G. N.; Schell, T. L.; Elliott, M. N.; Berthold, S. M.; Chun, C.; Marshall, Grant N.; Schell, Terry L.; Elliott, Marc N.; Berthold, S. Megan; Chun, Chi-Ah | 2005 | Mental health of Cambodian refugees 2 decades after resettlement in the United States | No natural experiment | |
| Malmusi, Davide | 2015 | Immigrants' health and health inequality by type of integration policies in European countries | No natural experiment | |
| Liu, D.; Tsegai, D.; Litaker, D.; von Braun, J. | 2015 | Under regional characteristics of rural China: a clearer view on the performance of the New Rural Cooperative Medical Scheme | No natural experiment | |
| Lofters, Aisha K.; Moineddin, Rahim; Hwang, Stephen W.; Glazier, Richard H. | 2011 | Predictors of low cervical cancer screening among immigrant women in Ontario, Canada | No natural experiment | |
| Liddell, B. J.; Nickerson, A.; Felmingham, K. L.; Malhi, G. S.; Cheung, J.; Den, M.; Askovic, M.; Coello, M.; Aroche, J.; Bryant, R. A. | 2019 | Complex Posttraumatic Stress Disorder Symptom Profiles in Traumatized Refugees | No natural experiment | |
| Lev-Wiesel, R.; Kaufman, R. | 2004 | Personal characteristics, unemployment, and anxiety among highly educated immigrants | No natural experiment | |
| Reus-Pons, Matias; Vandenheede, Hadewijch; Janssen, Fanny; Kibele, Eva U. B. | 2016 | Differences in mortality between groups of older migrants and older non-migrants in Belgium, 2001-09 | No natural experiment | |
| Hussen, Hozan Ismael; Persson, Martina; Moradi, Tahereh | 2013 | The trends and the risk of type 1 diabetes over the past 40 years: an analysis by birth cohorts and by parental migration background in Sweden | No natural experiment | |
| Heaton, T. B.; Leoprapai, B.; Cardona, R. | 1983 | Families, jobs and mobility: a comparison of migration streams in Thailand and Colombia | No natural experiment | |
| Almond, Douglas; Mazumder, Bhashkar | 2005 | The 1918 influenza pandemic and subsequent health outcomes: an analysis of SIPP data | No natural experiment | |
| Lv, N.; Brown, J. L. | 2011 | Impact of a nutrition education program to increase intake of calcium-rich foods by chinese-american women | No natural experiment | |
| Labrecque, J. A.; Kyle, R. P.; Joseph, L.; Bernatsky, S. | 2016 | Health-selective migration among patients with rheumatoid arthritis in Quebec :a cohort study using administrative data | No natural experiment | |
| Lazar-Neto, Felippe; Louzada, Andressa C. Sposato; de Moura, Ricardo Faé; Calixto, Fernando Morelli; Castro, Marcia C. | 2018 | Depression and Its Correlates Among Brazilian Immigrants in Massachusetts, USA | No natural experiment | |
| Daher, A. M.; Ibrahim, H. S.; Daher, T. M.; Anbori, A. K. | 2011 | Health related quality of life among Iraqi immigrants settled in Malaysia | No natural experiment | |
| Borrell, Carme; Palència, Laia; Bartoll, Xavier; Ikram, Umar; Malmusi, Davide | 2015 | Perceived Discrimination and Health among Immigrants in Europe According to National Integration Policies | No natural experiment | |
| Agyemang, C.; Meeks, K.; Beune, E.; Owusu-Dabo, E.; Mockenhaupt, F. P.; Addo, J.; de Graft Aikins, A.; Bahendeka, S.; Danquah, I.; Schulze, M. B.; et al., | 2016 | Obesity and type 2 diabetes in sub-Saharan Africans - Is the burden in today's Africa similar to African migrants in Europe? The RODAM study | No natural experiment | |
| Aldridge, R. W.; Zenner, D.; White, P. J.; Muzyamba, M. C.; Loutet, M.; Dhavan, P.; Mosca, D.; Hayward, A. C.; Abubakar, I. | 2016 | Prevalence of and risk factors for active tuberculosis in migrants screened before entry to the UK: a population-based cross-sectional study | No natural experiment | |
| Amin, R.; Mittendorfer-Rutz, E.; Mehlum, L.; Runeson, B.; Helgesson, M.; Tinghog, P.; Bjorkenstam, E.; Holmes, E. A.; Qin, P. | 2021 | Does country of resettlement influence the risk of suicide in refugees? A case-control study in Sweden and Norway | No natural experiment | |
| Zhuang, Xiao Yu; Wong, Daniel Fu Keung; Ng, Ting Kin; Poon, Ada | 2020 | Effectiveness of Mental Health First Aid for Chinese-Speaking International Students in Melbourne | No natural experiment | Claims to be natural experiment, but no random allocation |
| Havumaki, J.; Meza, R.; Phares, C. R.; Date, K.; Eisenberg, M. C. | 2019 | Comparing alternative cholera vaccination strategies in Maela refugee camp: using a transmission model in public health practice | No natural experiment | |

| | | | |
|---|---|---|---|
| Beiser, M.; Hou, F. | 2016 | Mental Health Effects of Premigration Trauma and Postmigration Discrimination on Refugee Youth in Canada | No natural experiment |
| MacDowell, H.; Pyakurel, S.; Acharya, J.; Morrison-Beedy, D.; Kue, J. | 2019 | Perceptions Toward Mental Illness and Seeking Psychological Help among Bhutanese Refugees Resettled in the US | No natural experiment |
| Madsen, C.; Gehring, U.; Walker, S. E.; Brunekreef, B.; Stigum, H.; Naess, O.; Nafstad, P. | 2010 | Ambient air pollution exposure, residential mobility and term birth weight in Oslo, Norway | No natural experiment |
| Mehta, Neil; Elo, Irma; Engelman, Michal; Lauderdale, Diane; Kestenbaum, Bert; Mehta, Neil K.; Elo, Irma T.; Lauderdale, Diane S.; Kestenbaum, Bert M. | 2016 | Life Expectancy Among U.S.-born and Foreign-born Older Adults in the United States: Estimates From Linked Social Security and Medicare Data | No natural experiment |
| Merten, S.; Wyss, C.; Ackermann-Liebrich, U. | 2007 | Caesarean sections and breastfeeding initiation among migrants in Switzerland | No natural experiment |
| Miller, Lauren; Robinson, Jonnell; Cibula, Donald | 2016 | Healthy Immigrant Effect: Preterm Births Among Immigrants and Refugees in Syracuse, NY | No natural experiment |
| Monnat, S. M. | 2017 | The New Destination Disadvantage: Disparities in Hispanic Health Insurance Coverage Rates in Metropolitan and Nonmetropolitan New and Established Destinations | No natural experiment |
| Lee, S.; Choi, S. H.; Proulx, L.; Cornwell, J. | 2015 | Community Integration of Burmese Refugees in the United States | No natural experiment |
| Kuroda, Yujiro; Iwasa, Hajime; Goto, Aya; Yoshida, Kazuki; Matsuda, Kumiko; Iwamitsu, Yumi; Yasumura, Seiji | 2017 | Occurrence of depressive tendency and associated social factors among elderly persons forced by the Great East Japan Earthquake and nuclear disaster to live as long-term evacuees: a prospective cohort study | No natural experiment |
| Knoblauch, A. M.; Divall, M. J.; Owuor, M.; Musunka, G.; Pascall, A.; Nduna, K.; Ng'uni, H.; Utzinger, J.; Winkler, M. S. | 2018 | Selected indicators and determinants of women's health in the vicinity of a copper mine development in northwestern Zambia | No natural experiment |
| Kauhl, B.; Pieper, J.; Schweikart, J.; Keste, A.; Moskwyn, M. | 2018 | Spatial Distribution of Type 2 Diabetes Mellitus in Berlin: Application of a Geographically Weighted Regression Analysis to Identify Location-Specific Risk Groups | No natural experiment |
| Maskileyson, Dina | 2019 | Health trajectories of immigrants in the United States: Does income inequality of country of origin matter? | No natural experiment |
| McLaughlin, Robert H. | 2012 | Criteria for Medical Repatriation and the Context of Inadequate Access to Care | No natural experiment |
| Mood, C.; Jonsson, J. O.; Laftman, S. B. | 2016 | Immigrant Integration and Youth Mental Health in Four European Countries | No natural experiment |
| Murray, K. E. | 2010 | Sudanese Perspectives on Resettlement in Australia | No natural experiment |
| Park, K.; Cho, Y.; Yoon, I. J. | 2009 | Social inclusion and length of stay as determinants of health among North Korean refugees in South Korea | No natural experiment |
| Pieterse, S.; Ismail, S. | 2003 | Nutritional risk factors for older refugees | No natural experiment |
| Reid, Alison; Peters, Susan; Felipe, Nieves; Lenguerrand, Erik; Harding, Seeromanie | 2016 | The impact of migration on deaths and hospital admissions from work-related injuries in Australia | No natural experiment |
| Sartorius, B.; Kahn, K.; Collinson, M. A.; Sartorius, K.; Tollman, S. M. | 2013 | Dying in their prime: determinants and space-time risk of adult mortality in rural South Africa | No natural experiment |
| Silove, D.; Liddell, B.; Rees, S.; Chey, T.; Nickerson, A.; Tam, N.; Zwi, A. B.; Brooks, R.; Sila, L. L.; Steel, Z. | 2014 | Effects of recurrent violence on post-traumatic stress disorder and severe distress in conflict-affected Timor-Leste: a 6-year longitudinal study | No natural experiment |
| Silverberg, J. I.; Simpson, E. L.; Durkin, H. G.; Joks, R. | 2013 | Prevalence of Allergic Disease in Foreign-Born American Children | No natural experiment |

| | | | |
|---|---|---|---|
| Tan, N. X.; Messina, J. P.; Yang, L. G.; Yang, B.; Emch, M.; Chen, X. S.; Cohen, M. S.; Tucker, J. D. | 2011 | A Spatial Analysis of County-level Variation in Syphilis and Gonorrhea in Guangdong Province, China | No natural experiment |
| Thapa, S. B.; Dalgard, O. S.; Claussen, B.; Sandvik, L.; Hauff, E. | 2007 | Psychological distress among immigrants from high- and low-income countries: Findings from the Oslo Health Study | No natural experiment |
| Tinghog, P.; Hemmingsson, T.; Lundberg, I. | 2007 | To what extent may the association between immigrant status and mental illness be explained by socioeconomic factors? | No natural experiment |
| Urmi, A. Z.; Leung, D. T.; Wilkinson, V.; Miah, M. A. A.; Rahman, M.; Azim, T. | 2015 | Profile of an HIV testing and counseling unit in Bangladesh: majority of new diagnoses among returning migrant workers and spouses | No natural experiment |
| Vahabi, M.; Lofters, A.; Kumar, M.; Glazier, R. H. | 2016 | Breast cancer screening disparities among immigrant women by world region of origin: a population-based study in Ontario, Canada | No natural experiment |
| Vilar-Compte, Mireya; Macinko, James; Weitzman, Beth C.; Avendaño-Villela, Carlos M. | 2019 | Short relative leg length is associated with overweight and obesity in Mexican immigrant women | No natural experiment |
| Vives, A.; Vanroelen, C.; Amable, M.; Ferrer, M.; Moncada, S.; Llorens, C.; Muntaner, C.; Benavides, F. G.; Benach, J. | 2011 | EMPLOYMENT PRECARIOUSNESS IN SPAIN: PREVALENCE, SOCIAL DISTRIBUTION, AND POPULATION-ATTRIBUTABLE RISK PERCENT OF POOR MENTAL HEALTH | No natural experiment |
| Ellis, B. H.; Lankau, E. W.; Ao, T.; Benson, M. A.; Miller, A. B.; Shetty, S.; Cardozo, B. L.; Geltman, P. L.; Cochran, J. | 2015 | Understanding Bhutanese Refugee Suicide Through the Interpersonal-Psychological Theory of Suicidal Behavior | No natural experiment |
| Elsafti, Abdallah Mohamed; van Berlaer, Gerlant; Al Safadi, Mohammad; Debacker, Michel; Buyl, Ronald; Redwan, Atef; Hubloue, Ives | 2016 | Children in the Syrian Civil War: the Familial, Educational, and Public Health Impact of Ongoing Violence | No natural experiment |
| Faturiyele, Iyiola; Karletsos, Dimitris; Ntene-Sealiete, Keletso; Musekiwa, Alfred; Khabo, Mantiti; Mariti, Marethabile; Mahasha, Phetole; Xulu, Thembisile; Pisa, Pedro T.; on behalf of, Equip Innovation for Health Team | 2018 | Access to HIV care and treatment for migrants between Lesotho and South Africa: a mixed methods study | No natural experiment |
| Feikin, D. R.; Adazu, K.; Obor, D.; Ogwang, S.; Vulule, J.; Hamel, M. J.; Laserson, K. | 2010 | Mortality and health among internally displaced persons in western Kenya following post-election violence, 2008: novel use of demographic surveillance | No natural experiment |
| Gilliland, J. A.; Shah, T. I.; Clark, A.; Sibbald, S.; Seabrook, J. A. | 2019 | A geospatial approach to understanding inequalities in accessibility to primary care among vulnerable populations | No natural experiment |
| Giraudo, Massimiliano; Bena, Antonella; Costa, Giuseppe | 2017 | Migrant workers in Italy: an analysis of injury risk taking into account occupational characteristics and job tenure | No natural experiment |
| Liang, Y.; Lu, W. Y.; Wu, W. | 2014 | Are social security policies for Chinese landless farmers really effective on health in the process of Chinese rapid urbanization? a study on the effect of social security policies for Chinese landless farmers on their health-related quality of life | No natural experiment |
| Yang, Min; Dijst, Martin; Helbich, Marco | 2018 | Mental health among migrants in Shenzhen, China: does it matter whether the migrant population is identified by hukou or birthplace? | No natural experiment |
| Ma, J.; Mitchell, G.; Dong, G. P.; Zhang, W. Z. | 2017 | Inequality in Beijing: A Spatial Multilevel Analysis of Perceived Environmental Hazard and Self-Rated Health | No natural experiment |
| Markovic, M.; Manderson, L.; Kelaher, M. | 2002 | The health of immigrant women: Queensland women from the former Yugoslavia | No natural experiment |
| Méjean, C.; Deschamps, V.; Bellin-Lestienne, C.; Oleko, A.; Darmon, N.; Hercberg, S.; Serge, H.; Castetbon, K.; Katia, C.; Méjean, C.; Deschamps, V.; Bellin-Lestienne, C.; Oleko, A.; Darmon, N.; Hercberg, S.; Serge, H.; Castetbon, K.; Katia, C. | 2010 | Associations of socioeconomic factors with inadequate dietary intake in food aid users in France (The ABENA study 2004-2005) | No natural experiment |
| Mudrazija, Stipica; López-Ortega, Mariana; Vega, William A.; Gutiérrez Robledo, Luis Miguel; Sribney, William | 2016 | Household Composition and Longitudinal Health Outcomes for Older Mexican Return Migrants | No natural experiment |

| | | | |
|---|---|---|---|
| Myers, W. P.; Westenhouse, J. L.; Flood, J.; Riley, L. W. | 2006 | An ecological study of tuberculosis transmission in California | No natural experiment |
| Ndetei, D. M. | 2008 | Retention or migration of mental health workers: psychiatrists in Kenya | No natural experiment |
| Owoeye, Olabisi; Khawaja, Manzar; Kinsella, Anthony; Russell, Vincent | 2011 | Counter-urbanisation during Ireland's 'Celtic Tiger' period - mental health implications | No natural experiment |
| Panjari, M.; Koplin, J. J.; Dharmage, S. C.; Peters, R. L.; Gurrin, L. C.; Sawyer, S. M.; McWilliam, V.; Eckert, J. K.; Vicendese, D.; Erbas, B.; Matheson, M. C.; Tang, M. L. K.; Douglass, J.; Ponsonby, A. L.; Dwyer, T.; Goldfeld, S.; Allen, K. J. | 2016 | Nut allergy prevalence and differences between Asian-born children and Australian-born children of Asian descent: a state-wide survey of children at primary school entry in Victoria, Australia | No natural experiment |
| Patel, S.; Schechter, M. T.; Sewankambo, N. K.; Atim, S.; Kiwanuka, N.; Spittal, P. M. | 2014 | Lost in Transition: HIV Prevalence and Correlates of Infection among Young People Living in Post-Emergency Phase Transit Camps in Gulu District, Northern Uganda | No natural experiment |
| Sulaiman-Hill, C. M. R.; Thompson, S. C. | 2012 | Afghan and Kurdish refugees, 8-20 years after resettlement, still experience psychological distress and challenges to well being | No natural experiment |
| Thapa, S. B.; Hauff, E. | 2005 | Gender differences in factors associated with psychological distress among immigrants from low- and middle-income countries - Findings from the Oslo Health Study | No natural experiment |
| Topal, K.; Eser, E.; Sanberk, I.; Bayliss, E.; Saatci, E. | 2012 | Challenges in access to health services and its impact on quality of life: a randomised population-based survey within Turkish speaking immigrants in London | No natural experiment |
| Wang, B. R.; Yu, S.; Noh, J. W.; Kwon, Y. D. | 2014 | Factors associated with self-rated health among North Korean defectors residing in South Korea | No natural experiment |
| Wong, Eunice; Marshall, Grant; Schell, Terry; Elliott, Marc; Babey, Susan; Hambarsoomians, Katrin | 2011 | The Unusually Poor Physical Health Status of Cambodian Refugees Two Decades After Resettlement | No natural experiment |
| Wong, F. K.; Chang, Y. L.; He, X. S.; Wong, Fu Keung Daniel; Chang, Ying Li; He, Xue Song | 2009 | Correlates of psychological wellbeing of children of migrant workers in Shanghai, China | No natural experiment |
| Yarnell, Christopher J.; Longdi, Fu; Manuel, Doug; Tanuseputro, Peter; Stukel, Therese; Pinto, Ruxandra; Scales, Damon C.; Laupacis, Andreas; Fowler, Robert A.; Fu, Longdi | 2017 | Association Between Immigrant Status and End-of-Life Care in Ontario, Canada | No natural experiment |
| Luque, John; Tarasenko, Yelena; Reyes-Garcia, Claudia; Alfonso, Moya; Suazo, Norma; Rebing, Laura; Ferris, Daron; Luque, John S.; Tarasenko, Yelena N.; Alfonso, Moya L.; Ferris, Daron G. | 2017 | Salud es Vida: a Cervical Cancer Screening Intervention for Rural Latina Immigrant Women | No natural experiment |
| Lupone, Christina D.; Daniels, Danielle; Lammert, Dawn; Borsuk, Robyn; Hobart, Travis; Lane, Sandra; Shaw, Andrea | 2020 | Lead Exposure in Newly Resettled Pediatric Refugees in Syracuse, NY | No natural experiment |
| Islam, M. M.; Sallu, S.; Hubacek, K.; Paavola, J. | 2014 | Migrating to tackle climate variability and change? Insights from coastal fishing communities in Bangladesh | No natural experiment |
| Kuntz, B.; Lampert, T. | 2013 | How Healthy is the Lifestyle of Adolescents in Germany? Results from the German Health Interview and Examination Survey for Children and Adolescents (KiGGS) | No natural experiment |
| El Ghaziri, Nahema; Blaser, Jérémie; Darwiche, Joëlle; Suris, Joan-Carles; Sanchis Zozaya, Javier; Marion-Veyron, Régis; Spini, Dario; Bodenmann, Patrick | 2019 | Protocol of a longitudinal study on the specific needs of Syrian refugee families in Switzerland | No natural experiment |
| Mbago, M. C. | 1994 | Some correlates of child mortality in the refugee populated regions in Tanzania | No natural experiment |
| Santavirta, Torsten | 2016 | Invited commentary: The long term impact of forced migration during childhood on adult health | No natural experiment |

| | | | |
|---|---|---|---|
| Teariki, Mary Anne | 2017 | Housing and health of kiribati migrants living in New Zealand | No natural experiment |
| Gentilini, Valeria; Bodini, Chiara; Di Girolamo, Chiara; Campione, Ilaria; Cavazza, Gabriele; Marzaroli, Paolo; Musti, Muriel; Perlangeli, Vincenza; Pandolfi, Paolo; Pizzi, Lorenzo; Riccio, Martina | 2020 | An ecological study on health inequalities in the city of Bologna (Emilia-Romagna Region, Northern Italy): bridging knowledge and action | No natural experiment |
| Lichter, Daniel T.; Johnson, Kenneth M. | 2021 | Opportunity and Place: Latino Children and America's Future | No natural experiment |
| Acevedo-Garcia, D. | 2001 | Zip code-level risk factors for tuberculosis: neighborhood environment and residential segregation in New Jersey, 1985-1992 | No natural experiment |
| Andrade, L. H.; Wang, Y. P.; Andreoni, S.; Silveira, C. M.; Alexandrino-Silva, C.; Siu, E. R.; Nishimura, R.; Anthony, J. C.; Gattaz, W. F.; Kessler, R. C.; Viana, M. C. | 2012 | Mental Disorders in Megacities: Findings from the Sao Paulo Megacity Mental Health Survey, Brazil | No natural experiment |
| Barreto, T. V.; Rodrigues, L. C. | 1992 | FACTORS INFLUENCING CHILDHOOD IMMUNIZATION IN AN URBAN AREA OF BRAZIL | No natural experiment |
| Bozorgmehr, K.; Jahn, R. | 2019 | Adverse health effects of restrictive migration policies: building the evidence base to change practice | No natural experiment |
| Barber, J.; Guo, M.; Nguyen, L. T.; Thomas, R.; Turin, T. C.; Vaska, M.; Naugler, C.; Coapt, | 2017 | Sociodemographic Correlates of Clinical Laboratory Test Expenditures in a Major Canadian City | No natural experiment |
| Abouzeid, M.; Philpot, B.; Janus, E. D.; Coates, M. J.; Dunbar, J. A. | 2013 | Type 2 diabetes prevalence varies by socio-economic status within and between migrant groups: analysis and implications for Australia | No natural experiment |
| Bakhtiari, Elyas; Olafsdottir, Sigrun; Beckfield, Jason | 2018 | Institutions, Incorporation, and Inequality: The Case of Minority Health Inequalities in Europe | No natural experiment |
| Affronti, Mario; Affronti, Andrea; Pagano, Salvatore; Soresi, Maurizio; Giannitrapani, Lydia; Valenti, Miriam; La Spada, Emanuele; Montalto, Giuseppe | 2013 | The health of irregular and illegal immigrants: analysis of day-hospital admissions in a department of migration medicine | No natural experiment |
| Ager, A.; Ager, W.; Long, L. | 1995 | The differential experience of Mozambican refugee women and men | No natural experiment |
| Agren, G.; Romelsjo, A. | 1992 | MORTALITY IN ALCOHOL-RELATED DISEASES IN SWEDEN DURING 1971-80 IN RELATION TO OCCUPATION, MARITAL-STATUS AND CITIZENSHIP IN 1970 | No natural experiment |
| Amirkhanian, Yuri; Kuznetsova, Anna; Kelly, Jeffrey; DiFranceisco, Wayne; Musatov, Vladimir; Avsukevich, Natalya; Chaika, Nikolay; McAuliffe, Timothy | 2011 | Male Labor Migrants in Russia: HIV Risk Behavior Levels, Contextual Factors, and Prevention Needs | No natural experiment |
| Amrhein, C. G.; Mackinnon, R. D. | 1984 | An elementary simulation model of the job matching process within an interregional setting | No natural experiment |
| Antón, J. I.; Muñoz de Bustillo, R.; Antón, José-Ignacio; Muñoz de Bustillo, Rafael | 2010 | Health care utilisation and immigration in Spain | No natural experiment |
| Bakker, L.; Cheung, S. Y.; Phillimore, J. | 2016 | The Asylum-Integration Paradox: Comparing Asylum Support Systems and Refugee Integration in The Netherlands and the UK | No natural experiment |
| Banke-Thomas, Aduragbemi; Agbemenu, Kafuli; Johnson-Agbakwu, Crista | 2019 | Factors Associated with Access to Maternal and Reproductive Health Care among Somali Refugee Women Resettled in Ohio, United States: A Cross-Sectional Survey | No natural experiment |
| Bao, H. J.; Fang, Y.; Ye, Q. Y.; Peng, Y. | 2018 | Investigating Social Welfare Change in Urban Village Transformation: A Rural Migrant Perspective | No natural experiment |
| Bardenheier, Barbara H.; Pavkov, Meda E.; Winston, Carla A.; Klosovsky, Alex; Yen, Catherine; Benoit, Stephen; Gravenstein, Stefan; Posey, Drew L.; Phares, Christina R. | 2019 | Prevalence of Tuberculosis Disease Among Adult US-Bound Refugees with Chronic Kidney Disease | No natural experiment |

| | | | |
|---|---|---|---|
| Barreto, T. V.; Rodrigues, L. C. | 1992 | Factors influencing childhood immunisation in an urban area of Brazil | No natural experiment |
| Beecher, B.; Reeves, J.; Eggertsen, L.; Furuto, S. | 2010 | International students' views about transferability in social work education and practice | No natural experiment |
| Beere, Paul; Keeling, Sally; Jamieson, Hamish | 2019 | Ageing, loneliness, and the geographic distribution of New Zealand's interRAI-HC cohort | No natural experiment |
| Begam, N Shamim; Srinivasan, Kannan; Mini, GK | 2016 | Is migration affecting prevalence, awareness, treatment and control of hypertension of men in Kerala, India? | No natural experiment |
| Beiser, M. | 2009 | Resettling refugees and safeguarding their mental health: lessons learned from the Canadian Refugee Resettlement Project | No natural experiment |
| Beiser, M.; Hamilton, H.; Rummens, J. A.; Oxman-Martinez, J.; Ogilvie, L.; Humphrey, C.; Armstrong, R.; Beiser, Morton; Hamilton, Hayley; Rummens, Joanna Anneke; Oxman-Martinez, Jacqueline; Ogilvie, Linda; Humphrey, Chuck; Armstrong, Robert | 2010 | Predictors of emotional problems and physical aggression among children of Hong Kong Chinese, Mainland Chinese and Filipino immigrants to Canada | No natural experiment |
| Bhugra, Dinesh | 2004 | Migration and mental health | No natural experiment |
| Bhui, K.; Abdi, A.; Abdi, M.; Pereira, S.; Dualeh, M.; Robertson, D.; Sathyamoorthy, G.; Ismail, H. | 2003 | Traumatic events, migration characteristics and psychiatric symptoms among Somali refugees - Preliminary communication | No natural experiment |
| Borodkin, F. M.; Alfiorov, V. M. | 1982 | A demoeconomic model of the rural sector | No natural experiment |
| Brazil, N. | 2017 | Spatial Variation in the Hispanic Paradox: Mortality Rates in New and Established Hispanic US Destinations | No natural experiment |
| Brisbois, M. D.; Silva, H. O.; Pereira, H. R.; Sethares, K. A. | 2016 | Self-perceived health and quality of life among Azorean deportees: a cross sectional descriptive study | No natural experiment |
| Chesnais, J. C. | 1988 | Population trends in the European Community, 1960-1986 | No natural experiment |
| Curtis, S. E.; Ogden, P. E. | 1986 | Bangladeshis in London: a challenge to welfare | No natural experiment |
| Dalgard, O. S.; Thapa, S. B.; Hauff, E.; McCubbin, M.; Syed, H. R. | 2006 | Immigration, lack of control and psychological distress: Findings from the Oslo Health Study | No natural experiment |
| Davidson, G. R.; Murray, K. E.; Schweitzer, R. | 2008 | Review of refugee mental health and wellbeing: Australian perspectives | No natural experiment |
| Joshi, Chandni; Russell, Grant; I. Hao Cheng; Kay, Margaret; Pottie, Kevin; Alston, Margaret; Smith, Mitchell; Chan, Bibiana; Vasi, Shiva; Lo, Winston; Wahidi, Sayed Shukrullah; Harris, Mark F. | 2013 | A narrative synthesis of the impact of primary health care delivery models for refugees in resettlement countries on access, quality and coordination | No natural experiment |
| Dufour, Darna L; Piperata, Barbara A | 2004 | Rural-• to-• urban migration in Latin America: An update and thoughts on the model | No natural experiment |
| Bond, A. R. | 1985 | Northern settlement family-style: labor planning and population policy in Noril'sk | No natural experiment |
| Boukhemis, K.; Zeghiche, A. | 1988 | Appraisal of rural-urban migration determinants: a case study of Constantine, Algeria | No natural experiment |
| Boulogne, Roxane; Jougla, Eric; Breem, Yves; Kunst, Anton E.; Rey, Grégoire | 2012 | Mortality differences between the foreign-born and locally-born population in France (2004 - 2007) | No natural experiment |
| Bourne, Paul Andrew; Hudson-Davis, Angela; Sharpe-Pryce, Charlene; Clarke, Jeffery; Solan, Ikhalfani; Rhule, Joan; Francis, Cynthia; Watson-Coleman, Olive; Sharma, Anushree; Campbell-Smith, Janinne | 2014 | Does International Migration influence mortality pattern and what role does murder and economics play | No natural experiment |

| | | | |
|---|---|---|---|
| Brabäck, Lennart; Vogt, Hartmut; Hjern, Anders | 2011 | Migration and asthma medication in international adoptees and immigrant families in Sweden | No natural experiment |
| Briem, Christopher; Morrison, Peter A | 2004 | How Migration Flows Shape the Elderly Population of Metropolitan Pittsburgh | No natural experiment |
| Kanamori, Mariko; Kondo, Naoki; Juarez, Sol P.; Cederstrom, Agneta; Stickley, Andrew; Rostila, Mikael | 2021 | Does increased migration affect the rural-urban divide in suicide? A register-based repeated cohort study in Sweden from 1991 to 2015 | No natural experiment |
| James, Poppy; Iyer, Aarti; Webb, Thomas L. | 2019 | The impact of post-• migration stressors on refugees' emotional distress and health: A longitudinal analysis | No natural experiment |
| Sundquist, K.; Frank, G. | 2004 | Urbanization and hospital admission rates for alcohol and drug abuse: a follow-up study of 4.5 million women and men in Sweden | No natural experiment |
| Priebe, S.; Matanov, A.; Gavrilovic, J. J.; McCrone, P.; Ljubotina, D.; Knezevic, G.; Kucukalic, A.; Franciskovic, T.; Schutzwohl, M. | 2009 | Consequences of Untreated Posttraumatic Stress Disorder Following War in Former Yugoslavia: Morbidity, Subjective Quality of Life, and Care Costs | No natural experiment |
| Tekeli-Yesil, Sidika; Isik, Esra; Unal, Yesim; Aljomaa Almossa, Fuad; Konsuk Unlu, Hande; Aker, Ahmet Tamer | 2018 | Determinants of Mental Disorders in Syrian Refugees in Turkey Versus Internally Displaced Persons in Syria | No natural experiment |
| Thela, Lindokuhle; Tomita, Andrew; Maharaj, Varsha; Mhlongo, Mpho; Burns, Jonathan K. | 2017 | Counting the cost of Afrophobia: Post-migration adaptation and mental health challenges of African refugees in South Africa | No natural experiment |
| Gillespie, Sarah; Cardeli, Emma; Sideridis, Georgios; Issa, Osob; Ellis, B. Heidi | 2020 | Residential mobility, mental health, and community violence exposure among Somali refugees and immigrants in North America | No natural experiment |
| Bogic, M.; Ajdukovic, D.; Bremner, S.; Franciskovic, T.; Galeazzi, G. M.; Kucukalic, A.; Lecic-Tosevski, D.; Morina, N.; Popovski, M.; Schutzwohl, M.; Wang, D. L.; Priebe, S. | 2012 | Factors associated with mental disorders in long-settled war refugees: refugees from the former Yugoslavia in Germany, Italy and the UK | No natural experiment |
| Lyles, Emily; Arhem, Jakob; El Khoury, Ghada; Trujillo, Antonio; Spiegel, Paul; Burton, Ann; Doocy, Shannon | 2021 | Multi-purpose cash transfers and health among vulnerable Syrian refugees in Lebanon: a prospective cohort study | No natural experiment |
| Mayer, Adam; Lopez, Maria Claudia; Johansen, Igor Cavallini; Moran, Emilio | 2021 | Hydropower, Social Capital, Community Impacts, and Self-Rated Health in the Amazon* | No natural experiment |
| Ye, Xin; Zhu, Dawei; He, Ping | 2021 | The Long-Term Impact of Adversity in Adolescence on Health in Middle and Older Adulthood: A Natural Experiment From the Chinese Send-Down Movement | No natural experiment |
| Dondero, Molly; Altman, Claire E. | 2020 | Immigrant policies as health policies: State immigrant policy climates and health provider visits among U.S. immigrants | No natural experiment |
| Burrows, Kate; Pelupessy, Dicky C.; Khoshnood, Kaveh; Bell, Michelle L. | 2021 | Environmental Displacement and Mental Well-Being in Banjarnegara, Indonesia | No natural experiment |
| Alegria M,Cruz-Gonzalez M,Alvarez K,Canino G,Duarte C,Bird H,Ramos-Olazagasti M,Markle SL,O'Malley I,Vila D,Shrout PE | 2022 | How Ethnic Minority Context Alters the Risk for Developing Mental Health Disorders and Psychological Distress for Latinx Young Adults | No natural experiment |
| Fakhoury J,Burton-Jeangros C,Consoli L,Duvoisin A,Jackson Y | 2022 | Association Between Residence Status Regularization and Access to Healthcare for Undocumented Migrants in Switzerland: A Panel Study | No natural experiment |
| Farnham A,Winkler MS,Zabre HR,Divall MJ,Fink G,Knoblauch A | 2022 | Spatial mobility and large-scale resource extraction: An analysis of community well-being and health in a copper mining area of Zambia | No natural experiment |
| Mahadevan R,Jayasinghe M | | Are Factors Associated with Adult Refugees' Settlement different from Well-Being? A Longitudinal Study focusing on Gender and Age in Australia | No natural experiment |
| Nickerson A,Kashyap S,Keegan D,Edwards B,Forrest W,Bryant RA,O'Donnell M,Felmingham K,McFarlane AC,Tol WA,Lenferink L,Hoffman J,Liddell BJ | 2022 | Impact of displacement context on psychological distress in refugees resettled in Australia: a longitudinal population-based study | No natural experiment |
| O'Donnell AW,Paolini S,Stuart J | | Distinct trajectories of psychological distress among resettled refugees: Community acceptance predicts resilience while low ingroup social support predicts clinical distress | No natural experiment |

| | | | | |
|---|---|---|---|---|
| Piancharoen P,Kosiyaporn H,Suphanchaimat R | 2022 | Equity of Social Health Insurance Coverage for Migrants in Thailand: A Concentration Index Analysis | No natural experiment | |
| Piccoli L,Wanner P | 2022 | The political determinants of the health of undocumented immigrants: a comparative analysis of mortality patterns in Switzerland | No natural experiment | |
| Shovaz FA,Mahmoodabadi HZ,Salehzadeh M | 2022 | Effectiveness of life skills training based on self-care on mental health and quality of life of married Afghan women in Iran | No natural experiment | Claims to be a quasi-experimental study, but is an intervention study. |
| Tong YY,Kim J | 2022 | Adolescents' exposure to classmates from non-immigrant families and adulthood volunteerism | No natural experiment | |
| Wei LL,Yang Y,Zhang J,Si LJ | 2022 | Rural-urban migration, family arrangement, and children's welfare: Evidence from China's rural areas | No natural experiment | IV: Population mobility is not an indicator for family arrangements |

#### Table S4: Adapted EPHPP

Adaptations of the Quality assessment tool for quantitative studies of the Effective Public Health Practice Project (EPHPP)

| Component | Question | Adaptation |
|---|---|---|
| **A) Selection Bias** | *(Q1) Are the individuals selected to participate in the study likely to be representative of the target population?*<br>Very likely<br>Somewhat likely<br>Not likely<br>Can't tell<br>*(Q2) What percentage of selected individuals agreed to participate (survey study) or were included in the sample selection (register study)?*<br>80 - 100% agreement<br>60 – 79% agreement<br>less than 60% agreement<br>Not applicable<br>Can't tell | No adaptation |
| **B) Study Design** | *Indicate the study design*<br>1 Randomized controlled trial<br>2 Controlled clinical trial<br>3 Cohort analytic (two group pre + post)<br>4 Case-control<br>5 Cohort (one group pre + post (before and after))<br>6 Interrupted time series<br>7 Other specify ___________________________<br>8 Can't tell<br><br>*Was the study described as randomized? If NO, go to Component C.*<br>No Yes<br>*If Yes, was the method of randomization described? (See dictionary)*<br>No Yes<br>*If Yes, was the method appropriate? (See dictionary)*<br>No Yes | *(Q1) Indicate the study design*<br>➔ Study designs adjusted for natural experiment studies as follows:<br>• Cohort analytic (two groups pre + post)<br>• Cohort (one group pre + post)<br>• Cross-sectional case-control<br>• Cross-sectional<br>• Repeated cross-sectional<br>• Interrupted time series<br>• Ecological study<br>• Other specify ______________________________<br>• Can't tell<br><br>*The three questions about randomization were summarised:*<br>➔ Adjusted as follows:<br>*(Q2) Were there any limitations in the random allocation of participants to the exposure through the natural experiment?*<br>Answer options:<br>No limitations<br>Few limitations<br>Several limitations<br>Can't tell |
| **C) Confounders** | *(Q1) Were there important differences between groups prior to the intervention?*<br>Yes<br>No<br>Can't tell<br><br>*The following are examples of confounders:*<br>Race<br>Sex<br>Marital status/family<br>Age<br>SES (income or class)<br>Education<br>Health status<br>Pre-intervention score on outcome measure<br><br>*(Q2) If yes, indicate the percentage of relevant confounders that were controlled (either in the design (e.g. stratification, matching) or analysis)?*<br>80 – 100% (most)<br>60 – 79% (some)<br>Less than 60% (few or none)<br>Can't Tell | No adaptation |
| **D) Blinding** | *(Q1) Was (were) the outcome assessor(s) aware of the intervention or exposure status of participants?*<br>1 Yes<br>2 No<br>3 Can't tell | Component deleted, not applicable. |

| | | |
|---|---|---|
| | *(Q2) Were the study participants aware of the research question?*<br>1 Yes<br>2 No<br>3 Can't tell | |
| **E) Data Collection Methods** | *(Q1) Were data collection tools shown to be valid?*<br>Yes<br>No<br>Can't tell<br><br>*(Q2) Were data collection tools shown to be reliable?*<br>Yes<br>No<br>Can't tell | No adaptation |
| **F) Withdrawls and Drop-outs** | *(Q1) Were withdrawals and drop-outs reported in terms of numbers and/or reasons per group?*<br>Yes<br>No<br>Can't tell<br>Not Applicable (i.e. one time surveys or interviews)<br><br>*(Q2) Indicate the percentage of participants completing the study. (If the percentage differs by groups, record the lowest).*<br>80 -100%<br>60 - 79%<br>less than 60%<br>Can't tell<br>Not Applicable (i.e. Retrospective case-control) | No adaptation, however hint for reviewers:<br>*Only applicable for longitudinal studies and refers to the drop-outs during follow-up. Adherence to treatment should be captured in the "intervention integrity" section below.* |
| **G) Intervention Integrity** | *(Q1) What percentage of participants received the allocated intervention or exposure of interest?*<br>1 80 -100%<br>2 60 - 79%<br>3 less than 60%<br>4 Can't tell<br><br>*(Q2) Was the consistency of the intervention measured?*<br>1 Yes<br>2 No<br>3 Can't tell<br><br>*(Q3) Is it likely that subjects received an unintended intervention (contamination or co-intervention) that may influence the results?*<br>4 Yes<br>5 No<br>6 Can't tell | No adaptation |
| **H) Analyses** | *(Q1) Indicate the unit of allocation (circle one)*<br>community<br>organization/institution<br>practice/office<br>individual<br><br>*(Q2) Indicate the unit of analysis (circle one)*<br>community<br>organization/institution<br>practice/office<br>individual<br><br>*(Q3) Are the statistical methods appropriate for the study design?*<br>1 Yes<br>2 No<br>3 Can't tell<br><br>*(Q4) Is the analysis performed by intervention allocation status (i.e. intention to treat) rather than the actual intervention received?*<br>1 Yes<br>2 No<br>3 Can't tell | No adaptation |

### Table S5: Results of the quality appraisal using the adapted EPHPP

| Quality appraisal (EPHPP) | A: Selection bias | | | B: Study Design | | | C: Confounders | | | E: Data collection method | | | F: Withdrawals and drop-outs | | | G: Intervention integrity | | | H: Analyses | | | | |
|---|---|---|---|---|---|---|---|---|---|---|---|---|---|---|---|---|---|---|---|---|---|---|---|
| **Author(s)** | **Q1** | **Q2** | **RATING** | **Q1** | **Q2** | **RATING** | **Q1** | **Q2** | **RATING** | **Q1** | **Q2** | **RATING** | **Q1** | **Q2** | **RATING** | **Q1** | **Q2** | **Q3** | **Q1** | **Q2** | **Q3** | **Q4** | **GLOBAL RATING** |
| Almquist & Miething 2022 | Very likely | Not applicable | Moderate | Repeated cross-sectional | Few limitations | Moderate | Yes | (60-79%) some | Moderate | Yes | Yes | Strong | Not applicable | Not applicable | | 80-100% | No | Yes | Individual | Community | Yes | Yes | Moderate |
| Black et al. 2015 | Somewhat likely | Can't tell | Weak | Cohort analytic | Few limitations | Moderate | Yes | Can't tell | Weak | Can't tell | Can't tell | Weak | Can't tell | Can't tell | Weak | Can't tell | Can't tell | Yes | Individual | Individual | Yes | Yes | Weak |
| Boje-Kovacs et al. 2022 | Very likely | 80-100% agreement | Strong | Cohort analytic | Few limitations | Strong | No | (80-100%) most | Strong | Yes | Yes | Strong | Can't tell | Can't tell | Weak | 80-100% | Yes | Yes | Community | Individual | Yes | Yes | Strong |
| Bozorgmehr & Razum 2015 | Somewhat likely | Not applicable | Moderate | Ecological study | Few limitations | Moderate | Yes | 60-79% (some) | Moderate | Can't tell | Can't tell | | Not applicable | Not applicable | | Less than 60% | Yes | Can't tell | Community | Community | Yes | Yes | Moderate |
| East & Friedson 2020a | Somewhat likely | Can't tell | Moderate | Cohort | Few limitations | Moderate | Can't tell | 60-79% (some) | Moderate | Yes | Yes | Strong | Can't tell | Can't tell | Moderate | 80-100% | Can't tell | Yes | Community | Individual | Yes | Yes | Moderate |
| East 2020b | Can't tell | Can't tell | Weak | Cohort analytic | Few limitations | Moderate | Can't tell | 80-100% (most) | Strong | Yes | Yes | Strong | Can't tell | Can't tell | Weak | Can't tell | Yes | Yes | Community | Individual | Yes | Yes | Moderate |
| Erdmann et al. 2021 | Very likely | Less than 60% agreement | Weak | Cohort | Few limitations | Moderate | Not applicable | Can't tell | Weak | Yes | Yes | Strong | Not applicable | Not applicable | | 80-100% | No | Yes | Individual | Individual | Yes | Yes | Weak |
| Fletcher et al. 2021 | Very likely | Can't tell | Moderate | Cohort analytic | Few limitations | Moderate | Yes | 80-100% (most) | Strong | Can't tell | Can't tell | Moderate | Yes | 80-100% | Strong | 80-100% | No | Yes | Organization/ Institution | Individual | Yes | No | Strong |
| Foverskov et al. 2022a | Very likely | 80-100% agreement | Strong | Cohort analytic | Few limitations | Moderate | No | (80-100%) most | Strong | Yes | Yes | Strong | No | Can't tell | Weak | 80-100% | Yes | Yes | Individual | Individual | Yes | Yes | Strong |
| Foverskov et al. 2022b | Very likely | 80-100% agreement | Strong | Cohort analytic | Few limitations | Moderate | No | (80-100%) most | Strong | Yes | Yes | Strong | No | Can't tell | Weak | 80-100% | Yes | Yes | Individual | Individual | Yes | Yes | Strong |
| Frey 2022 | Very likely | Can't tell | Moderate | Cross-sectional | Few limitations | Moderate | Can't tell | Can't tell | Weak | Yes | Yes | Strong | Not applicable | Not applicable | | 80-100% | No | Yes | Community | Individual | Yes | No | Weak |
| Fu & VanLandingham 2010 | Somewhat likely | 60-79% agreement | Moderate | Cohort analytic | Few limitations | Moderate | Yes | 80-100% (most) | Moderate | Yes | No | Moderate | Not applicable | Not applicable | | 80-100% | Yes | Yes | Individual | Individual | Yes | Yes | Moderate |

| | | | | | | | | | | | | | | | | | | | | | | | |
|---|---|---|---|---|---|---|---|---|---|---|---|---|---|---|---|---|---|---|---|---|---|---|---|
| Fu & VanLandingham 2012a | Somewhat likely | 60-79% agreement | Moderate | Cohort analytic | Few limitations | Moderate | Yes | 60-79% (some) | Moderate | Yes | No | Moderate | Not applicable | Not applicable | | 80-100% | Yes | Yes | Individual | Individual | Yes | Yes | Moderate |
| Fu & VanLandingham 2012b | Somewhat likely | 60-79% agreement | Moderate | Cohort analytic | Few limitations | Moderate | Yes | 60-79% (some) | Moderate | Yes | No | Moderate | Not applicable | Not applicable | | 80-100% | Yes | Yes | Individual | Individual | Yes | Yes | Moderate |
| Giacco et al. 2018 | Somewhat likely | Less than 60% agreement | Moderate | Cohort analytic | Few limitations | Moderate | Yes | 80-100% (most) | Strong | Yes | Yes | Strong | Yes | 80-100% | Strong | Can't tell | Can't tell | Can't tell | Organization/ Institution | Individual | Yes | Yes | Moderate |
| Gibson et al. 2010 | Very likely | 80 - 100% agreement | Strong | Cohort analytic | Few limitations | Moderate | Can't tell | Can't tell | Weak | yes | yes | Strong | Not applicable | Not applicable | Weak | 80-100% | No | No | Community | Individual | Yes | Yes | Moderate |
| Gibson et al. 2013 | Very likely | 80 - 100% agreement | Strong | Cohort analytic | Few limitations | Moderate | Yes | 80-100% (most) | Strong | Yes | Can't tell | Moderate | Not applicable | Not applicable | Weak | 60-79% | No | Can't tell | Community | Individual | Yes | Yes | Moderate |
| Grönqvist et al. 2012 | Very likely | 80 - 100% agreement | Strong | Cohort analytic | No limitations | Strong | Yes | 80-100% (most) | Strong | Yes | Yes | Strong | Yes | Less than 60% | Weak | 80-100% | Yes | No | Individual | Individual | Yes | Yes | Strong |
| Hainmüller et al. 2017 | Very likely | 80 - 100% agreement | Strong | Cohort analytic | Few limitations | Moderate | No | 80-100% (most) | Strong | Yes | Yes | Strong | Can't tell | Can't tell | Weak | Can't tell | No | Yes | Individual | Individual | Yes | Yes | Strong |
| Hajdu & Hajdu 2015 | Somewhat likely | Not applicable | Moderate | Cross-sectional | Several limitations | Weak | Yes | 60-79% (some) | Moderate | Yes | Yes | Strong | Not applicable | Not applicable | | 80-100% | Can't tell | Yes | Community | Individual | Yes | Can't tell | Weak |
| Hamad et al. 2020 | Very likely | 80 - 100% agreement | Strong | Cohort analytic | Few limitations | Strong | Yes | 80-100% (most) | Strong | Yes | Yes | Strong | Not applicable | Not applicable | | 80-100% | Yes | No | Individual | Individual | Yes | No | Strong |
| Hamilton et al. 2021 | Somewhat likely | Less than 60% agreement | Weak | Cohort analytic | Few limitations | Moderate | Yes | 80-100% (most) | Strong | Yes | Yes | Strong | Not applicable | Not applicable | | Unclear | No | Yes | Individual | Individual | Yes | Yes | Moderate |
| Honkaniemi et al. 2021 | Very likely | 80 - 100% agreement | Strong | Cohort analytic | No limitations | Strong | Yes | 80-100% (most) | Strong | Yes | Yes | Strong | Not applicable | Not applicable | | 80-100% | Yes | Yes | Individual | Individual | Yes | No | Moderate |
| Hori & Schafer 2010 | Very likely | 80 - 100% agreement | Strong | Cross-sectional | Several limitations | Weak | Yes | 60-79% (some) | Moderate | Yes | No | Moderate | Not applicable | Not applicable | | | No | No | Community | Individual | Can't tell | No | Weak |
| Hwang et al. 2011 | Somewhat likely | 80 - 100% agreement | Strong | Cohort analytic | Few limitations | Moderate | Yes | 80-100% (most) | Strong | Yes | Yes | Strong | Yes | 60-79% | Moderate | Less than 60% | Can't tell | No | Community | Individual | Yes | No | Moderate |

| | | | | | | | | | | | | | | | | | | | | | | | |
|---|---|---|---|---|---|---|---|---|---|---|---|---|---|---|---|---|---|---|---|---|---|---|---|
| Jaschke & Kosyakova 2021 | Somewhat likely | Less than 60% agreement | Moderate | Cohort analytic | Few limitations | Moderate | Yes | 80-100% (most) | Strong | Yes | Yes | Strong | Yes | Less than 60% | Moderate | 80-100% | Can't tell | Can't tell | Individual | Individual | Yes | Yes | Moderate |
| Juanmarti Mestres et al. 2020 | Somewhat likely | Can't tell | Moderate | Ecological study | No limitations | Strong | Yes | 80-100% (most) | Strong | Yes | Yes | Strong | Not applicable | Not applicable | | 80-100% | Can't tell | Can't tell | Community | Individual | Yes | Yes | Strong |
| Kaushal 2007 | Somewhat likely | Can't tell | Moderate | Cohort analytic | Few limitations | Moderate | Yes | 60-79% (some) | Moderate | No | Yes | Moderate | Can't tell | Can't tell | Weak | Can't tell | No | Yes | Community | Individual | Yes | Can't tell | Moderate |
| Lopez et al. 2016 | Very likely | Can't tell | Moderate | Cross-sectional | Few limitations | Moderate | Yes | 60-79% (some) | Moderate | No | Yes | Moderate | Not applicable | Not applicable | | Can't tell | No | Yes | Community | Individual | Yes | No | Weak |
| Lu et al. 2022 | Very likely | Can't tell | Moderate | Cross-sectional | Several limitations | Weak | Can't tell | 80-100% (most) | Moderate | Yes | Yes | Strong | Not applicable | Not applicable | | 80-100% | Yes | Yes | Individual | Individual | Yes | No | Weak |
| Mezuk et al. 2019 | Very likely | Can't tell | Moderate | Cohort analytic | Few limitations | Moderate | Yes | 60-79% (some) | Moderate | No | Yes | Moderate | Can't tell | Can't tell | Weak | Can't tell | Can't tell | Yes | Community | Individual | Yes | No | Weak |
| Raphael et al. 2020 | Very likely | 80 - 100% agreement | Strong | Cohort analytic | No limitations | Strong | No | 80-100% (most) | Strong | Yes | Yes | Strong | Can't tell | Can't tell | Weak | 80-100% | No | No | Individual | Individual | Yes | Yes | Strong |
| Reiss et al. 2012 | Very likely | 80 - 100% agreement | Strong | Cohort analytic | Few limitations | Moderate | Yes | Less than 60% (few or none) | Weak | Yes | Yes | Strong | Yes | 80-100% | Strong | 80-100% | Yes | Yes | Individual | Individual | Yes | Yes | Moderate |
| Schober & Zocher 2022 | Very likely | 80-100% agreement | Strong | Cohort analytic | Few limitations | Strong | No | 60-79% (some) | Moderate | Yes | Yes | Strong | Can't tell | Can't tell | Weak | 80-100% | No | Yes | Community | Individual | Yes | No | Moderate |
| Stillman et al. 2009 | Very likely | less than 60% agreement | Weak | Cohort analytic | No limitations | Moderate | Yes | 80-100% (most) | Strong | Yes | Yes | Strong | Not applicable | Not applicable | | 80-100% | Can't tell | Can't tell | Community | Individual | Yes | Yes | Moderate |
| Stillman et al. 2012 | Very likely | Less than 60% agreement | Weak | Cohort analytic | Few limitations | Moderate | Yes | 80-100% (most) | Strong | Yes | Yes | Strong | Not applicable | Not applicable | | 80-100% | Can't tell | Can't tell | Community | Individual | Yes | Yes | Moderate |
| Stillman et al. 2015 | Very likely | Less than 60% agreement | Weak | Cohort analytic | No limitations | Strong | Yes | 80-100% (most) | Strong | Yes | Yes | Strong | Yes | 60-79% | Moderate | 80-100% | Can't tell | Can't tell | Community | Community | Yes | No | Strong |
| Swartz et al. 2017 | Very likely | 80 - 100% agreement | Strong | Cohort analytic | Few limitations | Moderate | Can't tell | 60-79% (some) | Moderate | Yes | Yes | Strong | Can't tell | Can't tell | Weak | Can't tell | No | Yes | Community | Individual | Yes | Yes | Moderate |

| | | | | | | | | | | | | | | | | | | | | | | | |
|---|---|---|---|---|---|---|---|---|---|---|---|---|---|---|---|---|---|---|---|---|---|---|---|
| Toomey et al. 2014 | Somewhat likely | Not applicable | Moderate | Cohort | Several limitations | Weak | Yes | Less than 60% (few or none) | Weak | No | Yes | Moderate | Can't tell | Can't tell | Weak | Can't tell | No | Can't tell | Community | Individual | Yes | Can't tell | Weak |
| Torres et al. 2022 | Very likely | 80-100% agreement | Strong | Cohort analytic | Few limitations | Strong | No | (80-100%) most | Strong | Yes | Yes | Strong | Not applicable | Not applicable | | 80-100% | No | Can't tell | Individual | Individual | Yes | Yes | Strong |
| Venkataramani et al. 2017 | Somewhat likely | Less than 60% agreement | Weak | Cohort analytic | Few limitations | Moderate | Yes | Less than 60% (few or none) | Weak | Yes | Yes | Strong | Not applicable | Not applicable | | 80-100% | Yes | Yes | Individual | Individual | Yes | Yes | Moderate |
| Wenner et al. 2020 | Somewhat likely | 80 - 100% agreement | Moderate | Ecological study | Few limitations | Moderate | Yes | Less than 60% (few or none) | Weak | Yes | Yes | Strong | No | Can't tell | Weak | 80-100% | No | No | Individual | Community | Yes | Yes | Moderate |
| Wenner et al. 2022 | Very likely | Less than 60% agreement | Weak | Cross-sectional case-control | Can't tell | Weak | No | 60-79% (some) | Moderate | No | Yes | Moderate | Not applicable | Not applicable | | 80-100% | No | Yes | Individual | Individual | Yes | No | Weak |
| White et al. 2016 | Very likely | 80 - 100% agreement | Strong | Cohort analytic | Few limitations | Moderate | Yes | 60-79% (some) | Moderate | Yes | Yes | Strong | Not applicable | Not applicable | Weak | 80-100% | Can't tell | Can't tell | Community | Individual | Yes | Yes | Moderate |
| Worth 1963 | Can't tell | Less than 60% agreement | Weak | Cohort analytic | Several limitations | Weak | Can't tell | Less than 60% (few or none) | Weak | Can't tell | Can't tell | Moderate | Not applicable | Not applicable | | 80-100% | Yes | Yes | Individual | Individual | | Yes | Weak |
| Ye & Rodriguez 2021 | Very likely | Can't tell | Weak | Cohort analytic | Few limitations | Moderate | Yes | Can't tell | Weak | Can't tell | Can't tell | Weak | Can't tell | Can't tell | Weak | Can't tell | Yes | Yes | Community | Individual | Yes | Yes | Weak |

#### Table S6: Additional details of included studies

| Author & Year of publication | Title | Event giving rise to natural experiment | Country of study | Geographical level | total sample size | % of sample migrants | Type of study | Statistical methods* | Causal pathways considered (theoretical/ empirical) | Using migrants as an example for wider issues |
|---|---|---|---|---|---|---|---|---|---|---|
| Almquist & Miething 2022 | The impact of an unemployment insurance reform on incidence rates of hospitalisation due to alcohol-related disorders: a quasi-experimental study of heterogeneous effects across ethnic background, educational level, employment status, and sex in Sweden | social policy (restriction of health insurance policy) | Sweden | country | 5009832 | 25% | repeated cross-sectional | Regression discontinuity | Theoretical: compensation of income loss, investment in health-promoting goods and activites<br>Empirical: none | migrants as relevant subgroup (-> axis of marginalisation) |
| Black et al. 2015 | The Impact of the Great Migration on Mortality of African Americans: Evidence from the Deep South | geographical proximity to facilitator of migration | United States of America | country | 828179 | not reported | cohort analytic | Instrumental variable<br>Intention-to-treat | Theoretical: move from rural to urban areas<br>Empirical: evidence for increased smoking and drinking affecting rates of COPD, cirrhosis, chronic liver disease and cancer | migrant-specific |
| Boje-Kovacs et al. 2022 | Neighborhoods and mental health – evidence from a natural experiment in the public social housing sector | social policy (housing programme) | Denmark | regional (Copenhagen municipality) | 8175 | 47% | cohort analytic | Instrumental variable<br>Intention-to-treat | Theoretical: mental health affected by social interactions; mental health affected by structural or institutional factors.<br>Empirical: structural factors (housing quality, local area, health care fixed effects) | migrants as relevant subgroup (-> axis of marginalisation) |
| Bozorgmehr & Razum 2015 | Effect of Restricting Access to Health Care on Health Expenditures among Asylum-Seekers and Refugees: A Quasi-Experimental Study in Germany, 1994–2013 | healthcare access policy | Germany | country | 5 688 823 person-years | 100% | ecological cross-sectional study; ecological cohort analytic | Regression adjustment; Regression discontinuity<br>Intention-to-treat | Theoretical: entitlement restrictions prevent cheaper primary care and increase costly emergency care; negative health effects of collective accommodation centres<br>Empirical: higher costs in collective accommodation centres | migrant-specific research question, but results may be extrapolated (-> effect of restrictions) |
| East & Friedson 2020 | An Apple a Day? Adult food stamp eligibility and health care utilization among immigrants | social policy (food stamps) | USA | country | 11674 | 100% | cohort analytic | Difference in differences<br>Intention-to-treat | Theoretical: increased food consumption, household resources<br>Empirical: affordability of care (no effect) | migrants as example |
| East 2020 | The Effect of Food Stamps on Children's Health - Evidence from Immigrants' Changing Eligibility | social policy (food stamps) | USA | country | 5949 | 100% | cohort analytic | Difference in differences<br>Intention-to-treat | Theoretical: increased food consumption, improved nutritional content, child care & family stress, life-course effects from nutrition in utero and infant age<br>Empirical: increased food consumption, life-course | migrants as example |

| | | | | | | | | | | |
|---|---|---|---|---|---|---|---|---|---|---|
| | | | | | | | | | effects from nutrition in utero and infant age | |
| Erdmann et al. 2021 | Using independent cross-sectional survey data to predict post-migration health trajectories among refugees by estimating transition probabilities and their variances | migration policy (residential assignment) | Germany | regional (federal state) | 560 | 100% | repeated cross-sectional used to approximate longitudinal design | Transition probabilities | Theoretical: differing levels of noise and chemical pollution, connectivity and access to green spaces, infrastructure development, social connectivity, and accessibility of health care services<br>Empirical: none | migrants as example |
| Fletcher | The effects of foreign-born peers in US High Schools and Middle Schools | variation across grades within schools | USA | country | 78546 | 10% | cross-sectional study, cohort analytic | Regression adjustment | Theoretical: fewer risk behaviours, better health behaviours fewer behavioural problems among foreign-born students; Social ties to US-born as form of upward mobility; insulation through foreign-born; US-born increasing discrimination and harassment<br>Empirical: friendship networks (less risky behaviours among foreign-born peers), positive effects largest in small groups | migrant-specific |
| Foverskov et al. 2022a | Neighbourhood socioeconomic disadvantage and psychiatric disorders among refugees: a population-based, quasi-experimental study in Denmark | migration policy (residential assignment) | Denmark | country | 42067 | 100% | cohort analytic | regression adjustment intention-to-treat | Theoretical: exposure to neighbourhood disorder, crime, noise, and lack of amenities; lack of social support and cohesion<br>Empirical: none | migrant-specific research question, but results may be extrapolated (-> effect of neighbourhood disadvantage) |
| Foverskov et al. 2022b | Risk of Psychiatric Disorders Among Refugee Children and Adolescents Living in Disadvantaged Neighborhoods | migration policy (residential assignment) | Denmark | country | 18709 | 100% | cohort analytic | regression adjustment intention-to-treat | Theoretical: socioeconomic opportunities, health behaviours, social support, and collective efficacy<br>Empirical: none | migrant-specific research question, but results may be extrapolated (-> effect of neighbourhood disadvantage) |
| Frey | Getting under the Skin: The Impact of Terrorist Attacks on Native and Immigrant Sentiment | terrorist attack | Germany | country | 1033 | 100% | cross-sectional study | Regression adjustment | Theoretical: increase in anti-refugee sentiment, hostility & discrimination<br>Empirical: increase in anti-refugee sentiment, hostility & discrimination | migrants as example |
| Fu & VanLandingham 2012a | Mental Health Consequences of International Migration for Vietnamese Americans and the Mediating Effects of Physical Health and Social Networks: Results from a Natural Experiment Approach | migration policy (repatriation) | Vietnam/ USA | city | 703 | 18% | cohort analytic | Regression adjustment | Theoretical: acculturation. Incl. The acquisition of dietary patterns and a more sedentary lifestyle; psychological stresses associated with immigration (suggested protective factor language acquisition) | migrant-specific |

| | | | | | | | | | | |
|---|---|---|---|---|---|---|---|---|---|---|
| | | | | | | | | | Empirical: negative effect of acculturation, positive effect of language acquisition | |
| Fu & VanLandingham 2012b | Disentangling the Effects of Migration, Selection and Acculturation on Weight and Body Fat Distribution: Results from a Natural Experiment involving Vietnamese Americans, Returnees, and Never-leavers | migration policy (repatriation) | Vietnam/ USA | city | 709 | 18% | cohort analytic | Regression adjustment Mediation analysis via Structural Equation Modelling | Theoretical: disruption/strengthening of social networks, physical health Empirical: social networks and physical health buffer negative effects of migration | migrant-specific |
| Fu & VanLandingham 2010 | Mental and Physical Health Consequences of Repatriation for Vietnamese Returnees: A Natural Experiment Approach | migration policy (repatriation) | Vietnam/ USA | city | 709 | 18% | cohort analytic | Regression adjustment | Theoretical: predisposing factors, migration experience, physical and social environment after migration Empirical: length of stay in asylum camps, length of time back in Vietnam, number of communities liven in since returning, community reaction to return | migrant-specific |
| Giacco et al. 2018 | The same or different psychiatrists for in- and out-patient treatment? A multi-country natural experiment | healthcare/ service organisation | Belgium, England, Germany, Italy and Poland | hospital | 6369 | 14% | cohort analytic | Regression adjustment Intention-to-treat | Theoretical: fragmentation of specialisation Empirical: none | migrants as relevant subgroup (-> axis of marginalisation) |
| Gibson et al. 2010 | What happens to diet and child health when migration splits households? Evidence from a migration lottery program | migration policy (visa programme) | New Zealand, Tonga | country | 669 | 27% | cohort analytic | Instrumental variable | Theoretical: lower income, changes income composition & household size; relative prices of different foods in destination and home country Empirical: dietary change in children left behind dependent on lower income rather than change in household size or income composition | migrant-specific |
| Gibson et al. 2013 | Natural Experiment evidence on the effect of Migration on blood pressure and Hypertension | migration policy (visa programme) | New Zealand, Tonga | country | 638 | 25% | cohort analytic | Instrumental variable | Theoretical: hypertension triggered by anxiety and change in diet (esp. Sodium)/ physical activity Empirical: increased hypertension through stress and sodium | migrant-specific |
| Grönqvist et al. 2012 | Income inequality and health: Lessons from a refugee residential assignment program | migration policy (residential assignment) | Sweden | country | 65595 | 100% | cohort analytic | Regression adjustment | Theoretical: strong vs. weak income inequality hypothesis Empirical: none | migrants as example |
| Hainmueller et al. 2017 | Protecting unauthorized immigrant mothers improves their children’s mental health | migration policy (visa programme) | USA | regional (Oregon) | 5653 | 100% | cohort analytic | Regression discontinuity | Theoretical: reduced threat of deportation Empirical: none | migrant-specific |

| | | | | | | | | | | |
|---|---|---|---|---|---|---|---|---|---|---|
| | | | | | | | | Intention-to-treat | | |
| Hajdu & Hajdu 2015 | The Impact of Culture on Well-Being: Evidence from a Natural Experiment | international migration | 34 European countries for at least one round and 16 countries for all five rounds | Continental (Multiple European countries) | 12085 | 100% | cross-sectional study | Regression adjustment | Theoretical: individualism/ collectivism, interpersonal and institutional trust<br>Empirical: none | migrants as example |
| Hamad et al. 2020 | Association of Neighborhood Disadvantage with Cardiovascular Risk Factors and Events Among Refugees in Denmark | migration policy (residential assignment) | Denmark | municipaliti es/ parishes | 49305 | 100% | cohort analytic | Regression adjustment | Theoretical: limited walkability, availability of nutritious food, reduced employment opportunities, greater crime rates<br>Empirical: none | migrants as example |
| Hamilton et al. 2021 | DACA's Association With Birth Outcomes Among Mexican-Origin Mothers in the United States | migration policy (visa programme) | USA | country | 72613 | 100% | cohort analytic | Difference in differences<br>Intention-to-treat | Theoretical: reduced threat of deportation, provision of economic opportunity<br>Empirical: none | migrant-specific |
| Honkaniemi et al. 2021 | Psychiatric consequences of a father's leave policy by nativity: a quasi-experimental study in Sweden | social policy (parental leave) | Sweden | country | 198589 | 17% | cohort analytic | Regression discontinuit y<br>Intention-to-treat | Theoretical: biological (ie, hormonal) changes, psycho-social changes, work–family balance, family relationships, physical activity, decreased alcohol consumption<br>Empirical: none | migrants as relevant subgroup (-> axis of marginalisation) |
| Hori & Schafer 2010 | Social costs of displacement in Louisiana after Hurricanes Katrina and Rita | natural disaster | USA | regional | 10347 | 19% | cross-sectional study | Regression adjustment | Theoretical: disruption of social and economic networks<br>Empirical: none | migrant-specific |
| Hwang et al. 2011 | The Short-Term Impact of Involuntary Migration in China's Three Gorges: A Prospective Study | infrastructure development | China | regional | 770 | 55% | cohort analytic | Difference in differences | Theoretical: dismantling of social networks and economic circumstances<br>Empirical: none | migrant-specific |
| Jaschke & Kosyakova 2021 | Does facilitated and Early Access to the Healthcare System Improve Refugees' Health Outcomes? Evidence from a Natural Experiment in Germany | healthcare access policy | Germany | country | 5922 | 100% | cross-sectional study; cohort analytic | Regression adjustment<br>Intention-to-treat | Theoretical: practical hurdles of visiting a doctor, undiagnosed mental health issues during flight<br>Empirical: literacy (worse score without eHC access), post-migration stress act as moderators | migrant-specific research question, but results may be extrapolated (-> effect of restrictions) |
| Juanmarti Mestres et al. 2020 | The deadly effects of losing health insurance | migration policy (restricted | Spain | country | not reported | not reported | ecologica l cohort analytic | Difference in differences | Theoretical: treatment interruptions, lower diagnosis rates, no access to timely | migrant-specific research question, but results may be |

| | | | | | | | | | | |
|---|---|---|---|---|---|---|---|---|---|---|
| | | health care access) | | | | | | Intention-to-treat | treatment<br>Empirical: none | extrapolated (-> effect of restrictions) |
| Kaushal 2007 | Do food stamps cause obesity? Evidence from immigrant experience | social policy (food stamps) | USA | country | 489266 | 15% | cohort analytic | Difference in differences Intention-to-treat | Theoretical: could cause obesity if people would buy less food if they received cash, but food stamps might also lead to healthier food and lower stress<br>Empirical: none | migrants as example |
| Lopez et al. 2016 | Health Implications of an Immigration Raid: Findings from a Latino Community in the Midwestern United States | migration policy (migration enforcement) | USA | regional | 487 | 83% | cross-sectional study | Regression adjustment | Theoretical: community distrust in authorities, perception of public spaces as risky<br>Empirical: none | migrant-specific |
| Lu et al. 2022 | Heterogeneous Impact of Social Integration on the Health of Rural-to-Urban Migrants in China | variation in dialects | China | country | 117446 | 100% | cross-sectional study | Instrumenta l variable | Theoretical: changed health consciousness or behaviour, increased utilisation of health services<br>Empirical: none | migrant-specific |
| Mezuk et al. 2019 | Immigrant enclaves and risk of drug involvement among asylum-seeking immigrants in Sweden: A quasi-experimental study | migration policy (residential assignment) | Sweden | country | 1091328 | 5% | cohort analytic | Regression adjustment | Theoretical: strengthened social capital in areas of high migrant density and stability of social environments act as protective factors<br>Empirical: none | migrants as example |
| Raphael et al. 2020 | Neighborhood Deprivation and Mental Health Among Immigrants to Sweden | migration policy (residential assignment) | Sweden | country | 145310 | 33% | cohort analytic | Regression adjustment | Theoretical: Discrimination and fewer opportunities for integration, financial stability<br>Empirical: neighbourhood percent immigrants | migrants as example |
| Reiss et al. 2012 | Assessing the effect of regional deprivation on mortality avoiding compositional bias: a natural experiment | migration policy (residential assignment) | Germany | regional | 32661 | 100% | cohort analytic | Regression adjustment | Theoretical: none<br>Empirical: none | migrants as example |
| Schober & Zocher 2022 | Health-Care Utilization of Refugees: Evidence from Austria | migration policy (residential assignment) | Austria | regional (Upper Austria) | 9771 | 100% | cohort analytic | Regression adjustment | Theoretical: different availability of information, norms and attitudes among peers, presence of NGOs and professional support, differing regional practice styles<br>Empirical: none | migrant-specific |
| Stillman et al. 2015 | Miserable Migrants? Natural Experiment Evidence on International Migration and Subjective Well-Being | migration policy (visa programme) | New Zealand, Tonga | country | 254 | 40% | cohort analytic | Instrumenta l variable | Theoretical: relative positions fall after migration; lower relative income leads to unhappiness<br>Empirical: none | migrant-specific |
| Stillman et al. 2009 | Migration and mental health: Evidence from a natural experiment | migration policy (visa programme) | New Zealand, Tonga | country | 497 | 40% | cohort analytic | Instrumenta l variable | Theoretical: stressful migration process, positive impact from remittances and change of cultural setting | migrant-specific |

| | | | | | | | | | | |
|---|---|---|---|---|---|---|---|---|---|---|
| | | | | | | | | | Empirical: changes in income and employment explain only small part of mental health improvement | |
| Stillman et al. 2012 | The impact of immigration on child health: experimental evidence from a migration lottery program | migration policy (visa programme) | New Zealand, Tonga | country | 779 | 22% | cohort analytic | Instrumental variable | Theoretical: negative effect of unhealthy diets and decrease in physical activity, change in antenatal practices and health knowledge<br>Empirical: Change in consumption of meats, fats and milk, change in household composition; changes in income of limited importance | migrant-specific |
| Swartz et al. 2017 | Expanding Prenatal Care to Unauthorized Immigrant Women and the Effects on Infant Health | social policy (rollout of health insurance policy) | USA | regional (Oregon) | 213746 | 22% | cohort analytic | Difference in differences Intention-to-treat | Theoretical: direct effect, child health outcomes affected through healthcare utilisation of mother<br>Empirical: none | migrant-specific research question, but results may be extrapolated (-> effect of restrictions) |
| Toomey et al. 2014 | Impact of Arizona's SB 1070 Immigration Law on Utilization of Health Care and Public Assistance Among Mexican-Origin Adolescent Mothers and Their Mother Figures | migration policy (migration enforcement) | USA | city | 279 | 54% | cohort study | Regression adjustment | Theoretical: social stress theory (disadvantage exposes individuals to more stressful events)<br>Empirical: none | migrant-specific |
| Torres et al. 2022 | The Deferred Action for Childhood Arrivals program and birth outcomes in California: a quasi-experimental study | migration policy (visa programme) | USA | regional (California) | 57476 | 100% | cohort analytic | Difference in differences | Theoretical: improved employment, psychological wellbeing including reduced stress related to deportation, expanded access to healthcare<br>Empirical: none | migrant-specific |
| Venkataramani et al. 2017 | Health consequences of the US Deferred Action for Childhood Arrivals (DACA) immigration programme: a quasi-experimental study | migration policy (visa programme) | United States of America | country | 14973 | 100% | cohort analytic | Difference in differences Intention-to-treat | Theoretical: increased employment opportunities and income, raised future aspirations, eliminating risk of deportation<br>Empirical: none | migrant-specific |
| Wenner et al. 2020 | Differences in realized access to healthcare among newly arrived refugees in Germany: results from a natural quasi-experiment | healthcare access policy | Germany | regional (North-Rhine Westphalia) | 55452 person-quarters | 100% | ecological cohort analytic | Comparison of standardised incidence ratios Intention-to-treat | Theoretical: administrative effort, delays in referrals<br>Empirical: none | migrant-specific research question, but results may be extrapolated (-> effect of restrictions) |
| Wenner et al. 2022 | Inequalities in access to healthcare by local policy model among newly arrived refugees: evidence from population-based studies in two German states | healthcare access policy | Germany | country (2 regions: Berlin and Baden-Württemberg) | 863 | 100% | cross-sectional study | Regression adjustment | Theoretical: bureaucratic barriers<br>Empirical: none | migrant-specific research question, but results may be extrapolated (-> effect of restrictions) |

| | | | | | | | | | | |
|---|---|---|---|---|---|---|---|---|---|---|
| White et al. 2016 | Long-term Effects of Neighbourhood Deprivation on Diabetes Risk: Quasi-Experimental Evidence from a Refugee Dispersal Policy in Sweden | migration policy (residential assignment) | Sweden | country | 61386 | 100% | cohort analytic | Regression adjustment | Theoretical: lower purchasing power, reduced psychosocial resources, poorer food availability and walkability<br>Empirical: accumulation of effect consistent with abovementioned pathways | migrants as example |
| Worth 1963 | Urbanization and squatter resettlement as related to child health in Hong Kong | migration policy (residential assignment) | China | regional | 405 | 100% | cross-sectional study | Descriptive statistics | Theoretical: overcrowding, sanitation<br>Empirical: none | migrant-specific (results may be extrapolated?) |
| Ye & Rodriguez 2021 | Highly vulnerable communities and the Affordable Care Act: Health insurance coverage effects, 2010–2018 | healthcare access policy | USA | country | 2927402 | 28% | cohort analytic | Difference in differences; Propensity score matching | Theoretical: protection against chronic conditions<br>Empirical: none | migrants as relevant subgroup (-> axis of marginalisation) |

***** According to classification by Craig and colleagues (2)
